# Deep Learning of Fluorescence Lifetime Imaging Ophthalmoscopy for Type 2 Diabetes Classification

**DOI:** 10.64898/2026.08.04.26359728

**Authors:** SeongJun Kwon, Cecilia S. Lee, Aaron Y. Lee, Linying Zhang

## Abstract

**Purpose:** To evaluate whether fluorescence lifetime imaging ophthalmoscopy (FLIO) combined with deep learning can detect metabolic signatures for classification of type 2 diabetes mellitus (T2DM).

**Design:** Cross-sectional analysis of participants included in the AI-READI dataset (version 3) with FLIO imaging.

**Subjects:** 1,783 participants from the AI-READI dataset (version 3) with HbA1c measurements and FLIO imaging scans: 425 normoglycemic, 569 prediabetic, and 789 diabetic.

**Methods:** Glycemic groups were defined from self-reported diagnosis, glucose-lowering medication, and HbA1c. Center-of-mass mean lifetime (τCMM) maps, corrected for background and system timing offset, were generated from the raw time-resolved FLIO scans. Group differences in central-window τCMM were tested by one-way ANOVA and by age-adjusted analysis of covariance. Convolutional neural networks (CNNs), ResNet-18, and XGBoost were trained on τCMM maps for three-class (normal, prediabetic, diabetic) and two binary (normal vs. impaired; normal vs. diabetic) classification of T2DM and compared them with an age-only classifier and with models combining FLIO and age.

**Main Outcome Measures:** Macro-averaged area under the receiver operating characteristic curve (AUROC), sensitivity, specificity, accuracy, F1 score, and positive predictive value (PPV).

**Results:** Group-averaged τCMM was longer in prediabetic and diabetic than in normoglycemic participants in both the SSC (p < 0.001) and the LSC (p < 0.001), but only the SSC difference remained significant after age adjustment (p = 0.017). For 3-class classification, the CNN using FLIO achieved the highest AUROC (0.60 ± 0.04), compared with 0.51 ± 0.03 for an age-only classifier. ResNet-18 and XGBoost showed similar performance. Binary classification of normal vs. diabetic participants yielded AUROC 0.67 ± 0.04 (CNN), 0.64 ± 0.04 (ResNet-18), and 0.62 ± 0.04 (XGBoost). All three FLIO models exceeded the age-only classifier in all T2DM classification tasks, whereas adding age to the FLIO models did not improve model performance.

**Conclusions:** FLIO-derived τCMM maps capture retinal signals associated with diabetes status that exceed what age alone provides, but yield modest classification performance with current artificial intelligence models. These findings highlight both the potential and the challenges of using FLIO for early metabolic screening and monitoring, informing future development of clinically applicable imaging biomarkers.

## 1. Introduction

Type 2 diabetes mellitus (T2DM) is a major global health burden and a leading cause of vision loss worldwide. Early detection and monitoring of metabolic dysregulation are essential for preventing complications, including diabetic retinopathy (DR). Hemoglobin A1c (HbA1c), which reflects average blood glucose levels over the preceding two to three months, is widely used to assess long-term glycemic control in routine clinical care. However, HbA1c measurement requires invasive blood testing and may not capture tissue-specific metabolic changes.

The retina is uniquely positioned to serve as a window into assessing metabolic health through non-invasive retinal imaging techniques. Prior studies have explored the use of fundus photography and Optical Coherence Tomography (OCT) for diabetes and DR classification, demonstrating that retinal images contain predictive signals for systemic disease states, but these modalities provide limited insight into time-resolved biochemical processes. However, OCT and fundus autofluorescence (FAF) primarily capture static structural or intensity-based information and do not directly measure dynamic fluorescence properties.

Fluorescence lifetime imaging ophthalmoscopy (FLIO) is a non-invasive imaging technique that measures the decay time of endogenous retinal fluorophores following laser excitation.^1,2^ It utilizes a 473 nm laser for the excitation of fluorophores, and fluorescence emission is detected by a time-correlated single photon counting system in two distinct spectral channels: the short spectral channel (SSC: 498–560 nm) and the long spectral channel (LSC: 560–720 nm).^3^ The photons from the fluorophores, including flavin adenine dinucleotide (FAD), macular pigment, collagen, elastin, and advanced glycation end products (AGE), are considered to be predominantly detected in the SSC; and the fluorescence from A2E and lipofuscin, in the LSC.^4^

In contrast to FAF, FLIO quantifies both fluorescence intensity and the duration of the excited state (fluorescence lifetime), providing information about the biochemical metabolic processes within retinal tissue. Schweitzer *et al.* pioneered the application of time-resolved autofluorescence to the human fundus and demonstrated that fluorescence lifetime parameters reflect the biochemical and metabolic processes of retinal tissue.^1^ Subsequently, they showed that patients with T2DM, even in the absence of clinically apparent DR, exhibit prolonged fundus autofluorescence lifetimes, with specific lifetime components correlating with HbA1c levels.^5^ Schmidt *et al.* extended these findings to patients with non-proliferative diabetic retinopathy (NPDR), showing increased lifetimes across all Early Treatment Diabetic Retinopathy Study (ETDRS) grid regions relative to healthy controls.^6^ Collectively, these studies suggest that FLIO captures a metabolic signature of patients with T2DM that precedes structural retinal damage, motivating the investigation of FLIO as a pre-symptomatic marker of systemic glycemic state.

Despite the promising prior studies on FLIO and metabolic monitoring, most prior FLIO analyses apply multi-exponential curve fitting to model fluorescence decay signals, typically decomposing the signal into short and long lifetime components and deriving summary parameters such as amplitude-weighted mean lifetimes. While biologically interpretable, these approaches depend on predefined model assumptions, are sensitive to initialization and noise, and may not fully capture the complex, high-dimensional structure of fluorescence decay data. In parallel, artificial intelligence (AI), especially deep learning models, applied to retinal imaging modalities such as fundus photography and OCT have demonstrated strong performance in predicting systemic biomarkers and disease states directly from images, often without explicit feature engineering.^7–11^ Deep learning has been shown to estimate mean lifetime parameters comparable to those obtained with multiexponential fitting in fluorescence lifetime imaging microscopy (FLIM), the broader imaging modality on which FLIO is based.^12^ Despite these advances, deep learning approaches have not yet been explored for FLIO data, except one study where several state-of-the-art AI models were developed to classify smokers vs non-smokers based on FLIO signals, a goal substantially different from ours on T2DM classification.^13^

In this study, we investigate whether AI can detect metabolic signatures from FLIO to classify diabetes status. We evaluate multiple models, including convolutional neural networks (CNNs), ResNet, and XGBoost, to compare classification performance across model types. Our results provide insight into the potential of AI-based approaches for T2DM classification from FLIO.

## 2. Materials and Methods

### 2.1. Data Source

Data was obtained from the AI-READI (Artificial Intelligence Ready and Equitable Atlas for Diabetes Insights) (version 3), a publicly available, de-identified, multimodal dataset designed and optimized for artificial intelligence research in T2DM.^14–16^ This study did not constitute human subjects research and therefore did not require institutional review board approval or additional informed consent. The original AI-READI study was approved by the University of Washington Institutional Review Board (STUDY00016228), and all participants provided informed consent, including consent for future research use and sharing of their de-identified data. The study was conducted in accordance with the tenets of the Declaration of Helsinki.

### 2.2 Study Population and Outcome

We included participants with available HbA1c measurements and both short- and long-spectral-channel FLIO imaging. For participants with multiple FLIO imaging sessions, only the most recent imaging session was included in the study.

Participants were categorized into three glycemic groups using a diagnosis- and treatment-aware definition, which served as the ground-truth outcome labels. Participants were labeled diabetic if they self-reported a type 2 diabetes diagnosis, taking glucose-lowering medication (oral or injectable medications, including insulin), or had HbA1c > 6.4%; prediabetic if they did not report a diabetes diagnosis but reported a prediabetes diagnosis, or had HbA1c 5.7–6.4%; and normal otherwise. This definition classifies medically well-controlled, treated diabetic participants as diabetic regardless of their current HbA1c. An alternative definition using HbA1c alone (normal < 5.7%, prediabetic 5.7–6.4%, diabetic > 6.4%) was included as a sensitivity analysis (Supplementary Materials Section F.2 & F.3).

### 2.3 Spatial τCMM Mean Lifetime Map Generation

Each FLIO acquisition consists of a temporal stack of frames corresponding to discrete time bins that capture photon arrival times following laser excitation (Figure 1). For each pixel location (*x*, *y*), the sequence of photon counts across frames represents the temporal decay of fluorescence.

**Figure 1.**
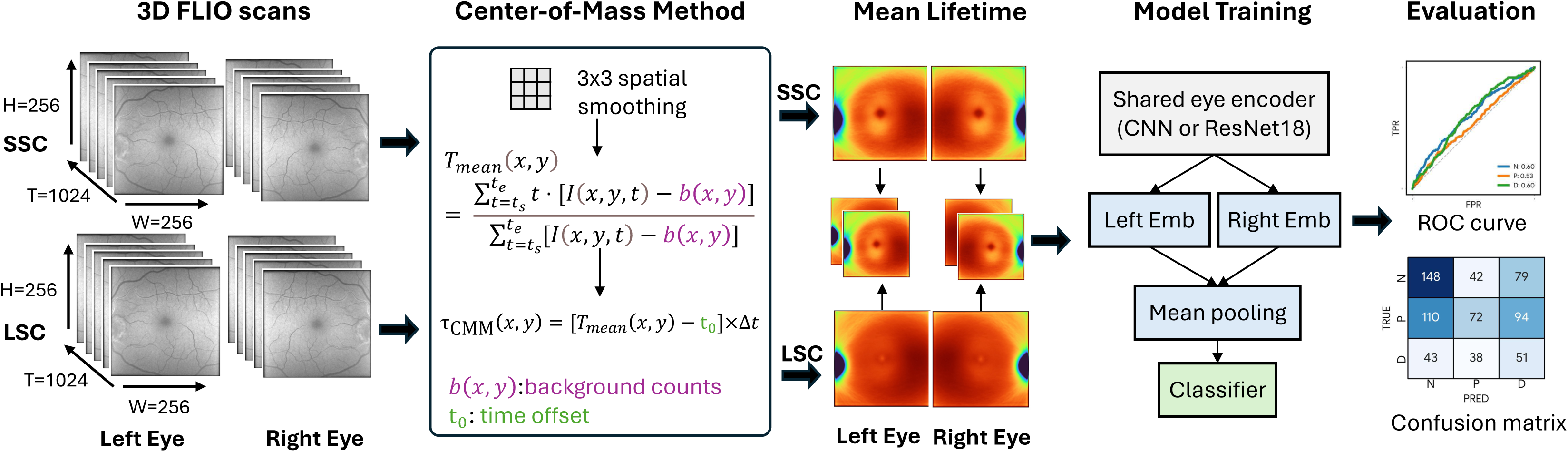
Schematic overview of the study workflow. Three-dimensional fluorescence lifetime imaging ophthalmoscopy (FLIO) scans underwent 3 × 3 average pooling, per-pixel background subtraction, and per-volume timing-offset correction to generate two-dimensional center-of-mass mean lifetime (τCMM) maps for the short- and long-spectral channels (SSC and LSC). These two-channel maps were stacked and used as inputs to deep learning models to classify participants’ glycemic status as normal, prediabetes, or diabetes.

#### Spatial Smoothing, Background Correction, and Photon Count Filtering

To reduce noise in photon measurements, spatial binning was applied prior to lifetime estimation. Specifically, for each temporal frame, pixel intensities were averaged within a 3 × 3 neighborhood. This smoothing operation reduces stochastic photon noise while preserving the overall spatial structure of the image.

Then, we removed ambient background counts. For each pixel, we estimated the background rate *b*(*x*, *y*) (counts per time bin) as the mean count over the time bins preceding the fluorescence rise, averaged over a 15 × 15-pixel neighborhood to suppress photon noise. The background was subtracted from every time bin of the active acquisition window, *t_s_* ≤ *t* ≤ *t_e_*, giving the net photon count for each pixel location (*x*, *y*):

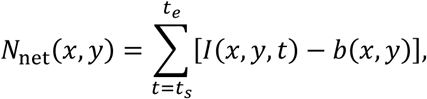

where *I*(*x*, *y*, *t*) represents the photon count at pixel (*x*, *y*) in time bin *t*. Pixels with insufficient photon counts produce unstable lifetime estimates. Therefore, pixels with net photon counts lower than 50 were excluded from analysis.

#### τCMM Estimation via the Center-of-Mass Method

Spatial maps were generated using the center-of-mass method (CMM). This method has been widely used in time-resolved fluorescence lifetime imaging as an alternative to parametric multi-exponential fitting.^17,18^ We denote the center-of-mass mean lifetime τCMM to distinguish it from the amplitude-weighted mean lifetime *τ_m_* conventionally reported in the FLIO literature.

The mean lifetime at each pixel was estimated as:

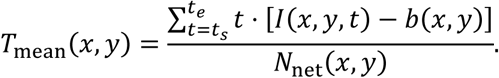

Second, recorded arrival times include a per-image system timing offset *t*_0_. To estimate it, the background-subtracted decays of all pixels of each image were summed, smoothed with a Gaussian kernel, and *t*_0_ was taken as the first time bin at which the smoothed decay reached 5% of its peak. The corrected τCMM was then obtained by removing the offset and converting time bins to physical time:

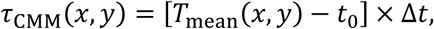

where Δ*t* denotes the frame time per bin. Details about the corrections are given in Supplementary Materials Section B. τCMM map computed without these corrections were evaluated as a sensitivity analysis (Supplementary Materials Section F.1 & F.3). This process yielded a two-dimensional spatial τCMM map for each eye and spectral channel.

### 2.4 Statistical Analysis of τCMM by Glycemic Group

To characterize spatial differences in mean τCMM across glycemic groups, pixel-wise lifetime maps were averaged within each group (normal, prediabetes, and diabetes) for each combination of eye laterality (left or right) and spectral channel (short or long). The resulting group-level maps were visualized to qualitatively compare spatial fluorescence lifetime patterns across glycemic groups (Figures 2 and 3).

**Figure 2.**
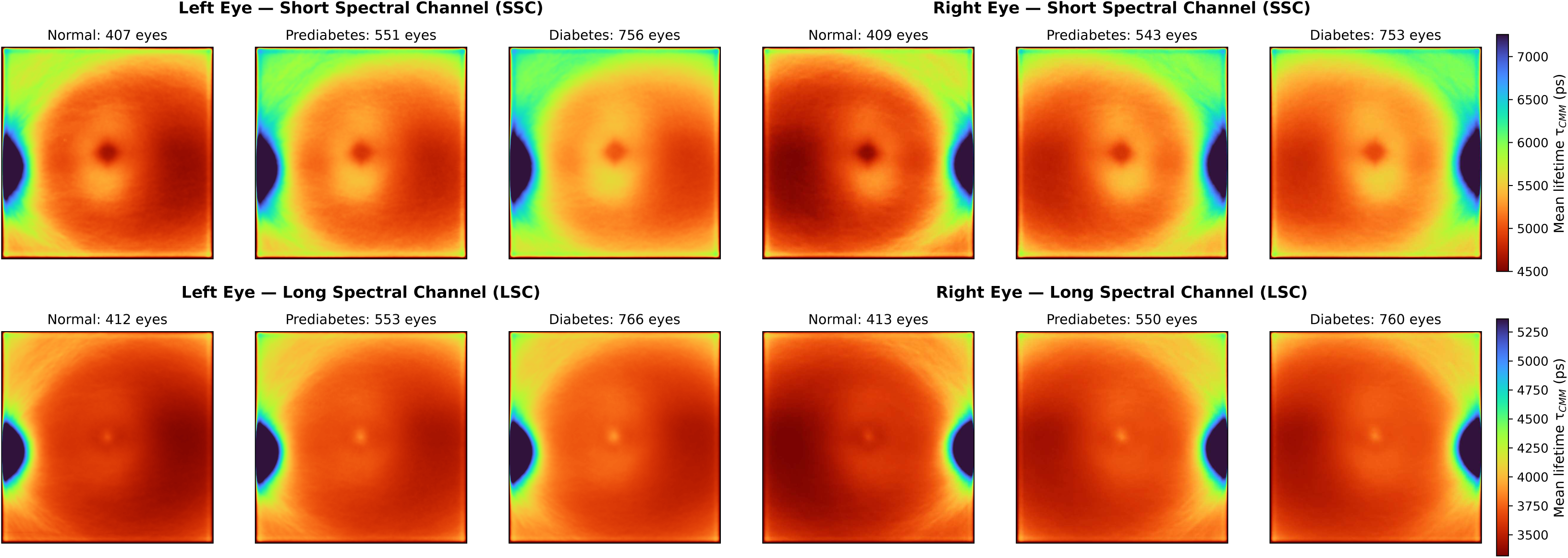
Group-averaged τCMM maps by glycemic group and spectral channel. Group-averaged spatial maps are shown for each combination of spectral channel (SSC, top; LSC, bottom) and eye laterality (Left, Right), separately for Normal, Prediabetes, and Diabetes participants. Following FLIO convention, the color scale runs from red (short) to blue (long); color bars are in picoseconds (ps). SSC = short spectral channel; LSC = long spectral channel.

**Figure 3.**
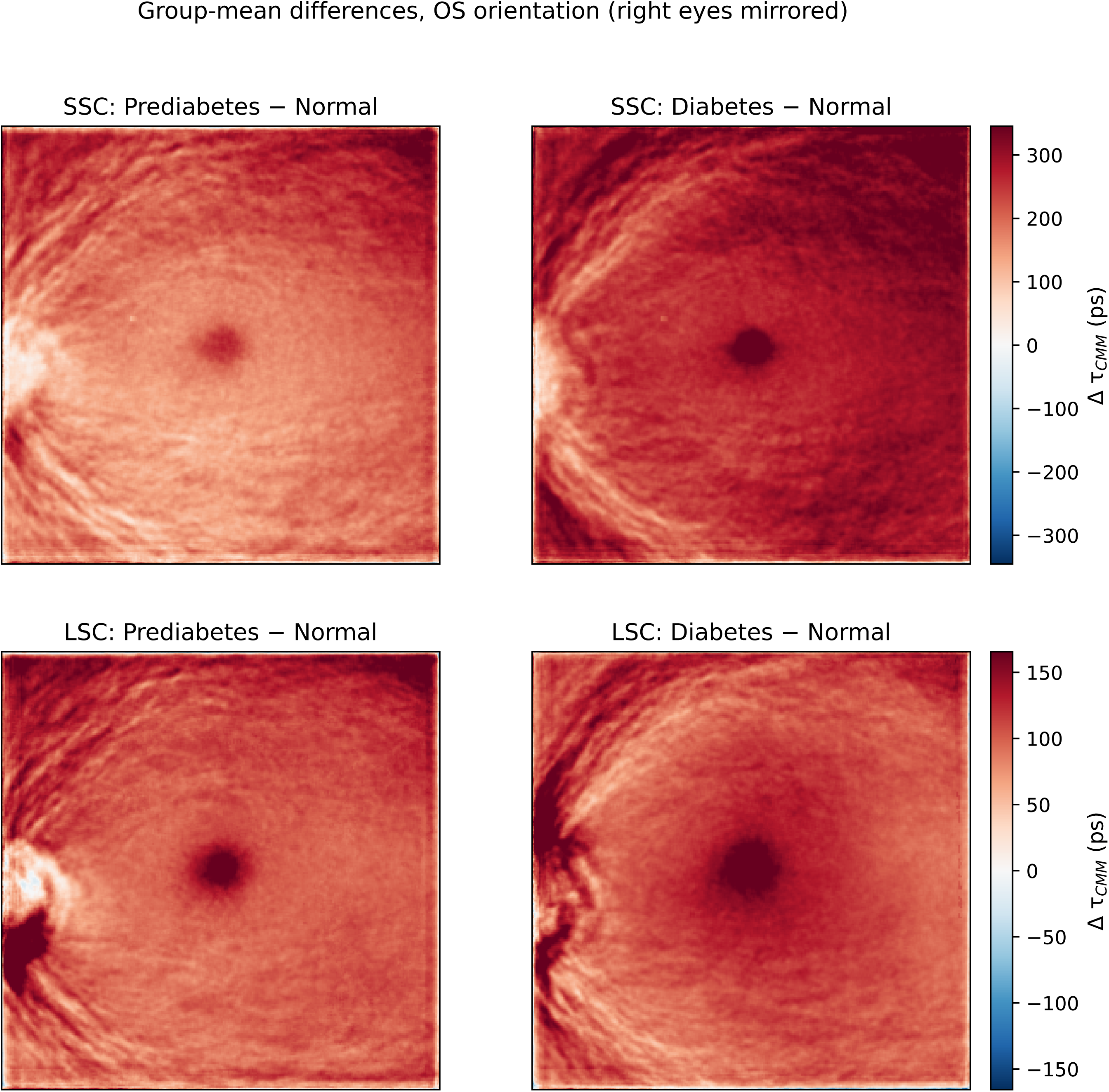
Group-difference maps of τCMM (diagnosis- and treatment-aware groups; background- and timing-offset-corrected maps). Pixel-wise differences of the group-averaged maps in OS orientation (right eyes mirrored horizontally before averaging; Prediabetes − Normal, left column; Diabetes − Normal, right column) for the short spectral channel (SSC, top row) and long spectral channel (LSC, bottom row). Panels show unadjusted group-mean differences; warm colors indicate longer τCMM in the impaired-glycemia group. Color bars are in picoseconds (ps). Corresponding maps for the alternative label and map definitions are shown in Supplementary Materials Figures A9, A13, and A17. τCMM = center-of-mass mean lifetime.

For quantitative comparisons, the mean τCMM was calculated within a square window centered on the image (not on the fovea); window position therefore does not depend on foveal detection, but it can differ from the anatomical foveal center depending on fixation. The primary comparison used a fixed 20 × 20-pixel central window (Figure 4), corresponding to approximately 2.3° of the 30° imaging field; the window location and size relative to the 256 × 256-pixel image and the ETDRS grid are shown in Supplementary Materials Figure A1. Pixels within the window were averaged to obtain an eye-level estimate; for participants with measurements from both eyes, the eye-level estimates were averaged to obtain a single participant-level value. Overall differences among the three glycemic groups were assessed using one-way analysis of variance (ANOVA), with *p* < 0.05 indicating that at least one group had a different mean τCMM. Pairwise differences were evaluated using Welch’s *t*-tests for three prespecified comparisons: normal versus prediabetes, normal versus diabetes, and prediabetes versus diabetes. Pairwise *p* values were adjusted using the Bonferroni correction. Because age differed across glycemic groups, the central-window comparison was additionally repeated as an analysis of covariance (ANCOVA) adjusting for age; sex, race, and ethnicity were not available for adjustment in the dataset.

**Figure 4.**
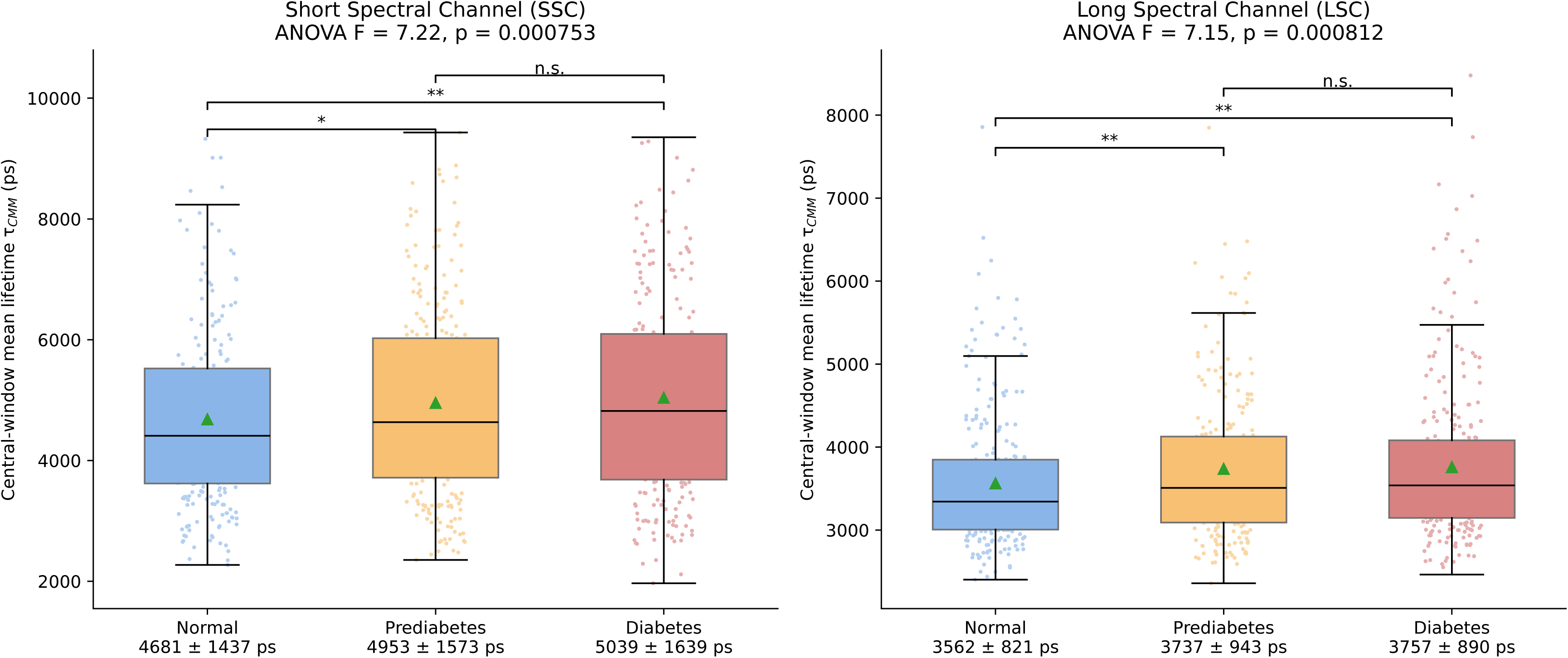
Participant-level distributions of τCMM within a 20 × 20-pixel window centered on the image, by glycemic group and spectral channel (unadjusted for age). Left: short spectral channel (SSC). Right: long spectral channel (LSC). Each participant’s value is the window mean averaged over the available eyes. Box plots show the interquartile range with median (horizontal line) and mean (triangle); overlaid points represent individual participants (subsampled for visual density). One-way ANOVA F and p values are shown in each panel title; after age adjustment, the SSC difference remained significant (ANCOVA p = 0.017), whereas the LSC difference did not (p = 0.16). Brackets denote pairwise Welch’s t-tests with Bonferroni correction across three contrasts. n.s. = not significant (p ≥ 0.05); *p < 0.05; **p < 0.01. Group means ± standard deviation (ps) are shown below each x-axis label. ANOVA = analysis of variance; ANCOVA = analysis of covariance; τCMM = center-of-mass mean lifetime.

As a sensitivity analysis, these comparisons were repeated across central-window sizes ranging from 10 × 10 to 190 × 190 pixels in increments of 10 pixels, separately for the short and long spectral channels, to evaluate the spatial extent and robustness of the observed group differences. The complete window-size sweep and corresponding results are presented in Supplementary Materials Section C. All tests were two-sided, and Bonferroni-adjusted *p* < 0.05 was considered statistically significant.

### 2.5 Model Development and Evaluation

We developed and evaluated convolutional neural network (CNN), ResNet-18, and extreme gradient boosting (XGBoost) models to classify glycemic status from spatial τCMM maps. All models made one prediction per participant. The primary task was three-class classification of normal, prediabetic, and diabetic status; we also evaluated two binary classification tasks: normal versus impaired glycemia and normal versus diabetes.

Because retinal anatomy is approximately mirror-symmetric between fellow eyes, right-eye maps were horizontally flipped to the left-eye (OS) orientation before encoding (laterality canonicalization), so that both eyes share a common anatomical frame; canonicalization was applied during training and inference for all models.

#### CNN inputs and architecture

For each eye, the SSC and LSC τCMM maps (256 × 256 pixels), each standardized to zero mean and unit variance over its valid pixels, were stacked as a two-channel input image and processed by a shared CNN encoder. The encoder consisted of four convolutional blocks with 3 × 3 kernels; each block included batch normalization and rectified linear unit activation, the first three were followed by max pooling, and the number of feature channels increased from 32 to 256 across successive blocks. The final convolutional features were aggregated by adaptive average pooling into a 256-dimensional eye embedding.

The embeddings of a participant’s available eyes (one or two) were mean-pooled into a participant-level embedding, which was then passed to a fully connected classification head with dropout regularization. The output layer generated class logits, which were converted to class probabilities using a softmax function. Variations in CNN depth, channel width, and regularization were evaluated during model development; detailed configurations are provided in the Supplementary Materials Section A.

#### ResNet-18 inputs and architecture

ResNet-18 was evaluated as a second image-based model using the same two-channel τCMM input, laterality canonicalization, and participant-level mean pooling as the CNN, with the first convolutional layer adapted to two input channels. ResNet-18 uses residual blocks with identity skip connections to facilitate optimization of deeper networks. ResNet-18 was trained from random initialization and class-weighted cross-entropy loss. Additional architectural and implementation details are provided in the Supplementary Materials Section A.

#### XGBoost inputs and architecture

Unlike the image-based models, XGBoost was trained using summary features extracted from the laterality-canonicalized τCMM maps: the mean τCMM within the superior, nasal, inferior, and temporal sectors of the inner ring of the Early Treatment Diabetic Retinopathy Study (ETDRS) grid for the SSC and of the outer ring for the LSC, following prior work.^13^ Sector means were averaged over a participant’s eyes and supplemented with derived features (per-sector SSC − LSC differences, SSC/LSC ratios, and SSC–LSC means; the mean, standard deviation, minimum, maximum, and range across the eight sector means; and four within-channel products of sector means), yielding 29 features. Hyperparameters—including tree depth, learning rate, subsampling rate, and regularization parameters—were selected within the inner cross-validation procedure (Supplementary Materials Section A).

#### FLIO + age models

To assess whether FLIO adds information beyond age, each model was also trained with age as an additional input. For the CNN and ResNet-18, age, standardized to zero mean and unit variance, was concatenated to the pooled participant embedding before the classification head; for XGBoost, age was added to the feature set. An XGBoost classifier trained on age alone served as the age-only baseline.

#### Classification tasks

The three-class task classified participants as having normal glycemia, prediabetes, or diabetes. Because prediabetes represents an intermediate glycemic state that may overlap with both neighboring groups, we additionally evaluated two binary tasks. For the normal-versus-impaired task, participants with prediabetes or diabetes were combined into an impaired-glycemia group. For the normal-versus-diabetes task, participants with prediabetes were excluded. The binary tasks used the same model architectures and nested cross-validation framework as the three-class task, with the output layers modified for two-class prediction and class-weighted loss calculated using two classes. Results for the binary tasks are reported at the participant level in Table 3. A secondary analysis predicting continuous HbA1c levels is described in Supplementary Materials Section D.

#### Model training and cross-validation

All models were trained and evaluated using nested five-fold cross-validation. Participants were assigned to five stratified outer folds, with both eyes from a given participant retained in the same fold to prevent information leakage. In each iteration, four outer folds were used for model development and the remaining fold was held out for evaluation. Hyperparameter selection was conducted exclusively within the outer training set using an inner stratified five-fold cross-validation procedure. The selected model was then evaluated on the held-out outer test fold. This process was repeated until each outer fold had served once as the test set. Performance metrics were calculated separately for each outer test fold and summarized as the mean ± standard deviation across the five folds.

#### Performance evaluation

For the three-class task, accuracy, F1 score, sensitivity, specificity, area under the receiver operating characteristic curve (AUROC), and positive predictive value (PPV) were calculated for each class and then macro-averaged across the three classes. For binary tasks, the corresponding binary classification metrics were reported. Confusion matrices were constructed by pooling predictions from the five held-out outer test folds.

### 2.6 Model Interpretation

To interpret model predictions, we applied Gradient-weighted Class Activation Mapping (Grad-CAM)^19^ to the CNN model as it was the best performing model. Grad-CAM generates saliency maps by highlighting regions in the input image that contribute most strongly to the model’s predictions.

For each participant, Grad-CAM was computed from the final convolutional layer of the CNN on participants in the outer fold held out for evaluation. To obtain a robust representation of model attention rather than individual sample variability, we averaged the per-eye Grad-CAMs across all test eyes, stratified by confusion-matrix cell (true × predicted class), to visualize attention patterns in both correctly classified and misclassified cases (Figure 6).

### 2.7 Sensitivity Analyses

We conduct sensitivity analyses to assess the robustness of the group-averaged difference and model performance on classifying glycemic states under three alternative conditions: (1) with and without adjustment for comorbidities, (2) using alternative definitions of HbA1c-based glycemic status, and (3) using uncorrected τCMM mean lifetime maps.

#### Comorbidities

We assess the sensitivity of group-averaged τCMM difference and model performance with respect to systemic and ocular comorbidities. Group comparisons and model classification were repeated after excluding participants with self-reported diabetic retinopathy, cataract, glaucoma, age-related macular degeneration (AMD), or any of the ocular conditions. Group comparisons were additionally repeated after excluding participants with each self-reported systemic condition in Table 1 (hypertension, high cholesterol, obesity, kidney disease, and cardiovascular disease), with any of these systemic conditions, and with any ocular or systemic condition.

**Table 1.** Cohort characteristics. Values are mean ± standard deviation (SD) unless otherwise indicated. Glycemic history and treatment, systemic, and ocular comorbidities are self-reported conditions and rows are n (%). Cardiovascular disease includes heart attack, stroke, other heart issues, or circulation problems. Systolic blood pressure and body mass index were measured at the study visit. Glycemic groups defined by the diagnosis- and treatment-aware label. τCMM values are background- and timing-offset-corrected mean lifetimes. HbA1c = hemoglobin A1c; BP = blood pressure; ps = picoseconds; SSC = short spectral channel; LSC = long spectral channel; τCMM = center-of-mass mean lifetime; RVO = retinal vascular occlusion; AMD = age-related macular degeneration.

|  | **Overall (n = 1,783)** | **Normal (n = 425)** | **Prediabetic (n = 569)** | **Diabetic (n = 789)** |
|---|---|---|---|---|
| **Demographics** |  |  |  |  |
| **Age (years)** | 60.5 ± 11.2 | 58.4 ± 11.4 | 60.7 ± 11.1 | 61.6 ± 10.9 |
| **Measurements** |  |  |  |  |
| **HbA1c (%)** | 6.10 ± 1.10 | 5.35 ± 0.22 | 5.77 ± 0.26 | 6.74 ± 1.36 |
| **Systolic BP (mmHg)** | 127.7 ± 16.0 | 126.1 ± 15.2 | 127.5 ± 15.6 | 128.7 ± 16.7 |
| **Body mass index (kg/m²)** | 30.0 ± 7.2 | 27.6 ± 6.2 | 29.6 ± 7.0 | 31.6 ± 7.5 |
| **Ocular Characteristics** |  |  |  |  |
| **SSC τCMM (ps)** | 5384.0 ± 1187.8 | 5190.7 ± 1019.4 | 5419.0 ± 1192.6 | 5462.4 ± 1256.2 |
| **LSC τCMM (ps)** | 3778.7 ± 586.7 | 3688.5 ± 500.5 | 3801.3 ± 577.5 | 3811.0 ± 630.4 |
| **Glycemic history and treatment, n (%)** |  |  |  |  |
| **Type 2 diabetes** | 677 (38.0%) | 0 (0.0%) | 0 (0.0%) | 677 (85.8%) |
| **Prediabetes** | 434 (24.3%) | 0 (0.0%) | 339 (59.6%) | 95 (12.0%) |
| **Oral or non-insulin-injectable medication** | 520 (29.2%) | 0 (0.0%) | 0 (0.0%) | 520 (65.9%) |
| **Insulin use** | 192 (10.8%) | 0 (0.0%) | 0 (0.0%) | 192 (24.3%) |
| **Systemic comorbidities, n (%)** |  |  |  |  |
| **Hypertension** | 891 (50.0%) | 122 (28.7%) | 234 (41.1%) | 535 (67.8%) |
| **High cholesterol** | 893 (50.1%) | 137 (32.2%) | 265 (46.6%) | 491 (62.2%) |
| **Obesity** | 647 (36.3%) | 94 (22.1%) | 185 (32.5%) | 368 (46.6%) |
| **Kidney disease** | 188 (10.5%) | 21 (4.9%) | 43 (7.6%) | 124 (15.7%) |
| **Cardiovascular disease** | 448 (25.1%) | 69 (16.2%) | 114 (20.0%) | 265 (33.6%) |
| **Ocular comorbidities, n (%)** |  |  |  |  |
| **Diabetic retinopathy** | 72 (4.0%) | 1 (0.2%) | 1 (0.2%) | 70 (8.9%) |
| **RVO** | 19 (1.1%) | 6 (1.4%) | 3 (0.5%) | 10 (1.3%) |
| **Cataract** | 528 (29.6%) | 97 (22.8%) | 154 (27.1%) | 277 (35.1%) |
| **Glaucoma** | 138 (7.7%) | 18 (4.2%) | 40 (7.0%) | 80 (10.1%) |
| **AMD** | 90 (5.0%) | 11 (2.6%) | 38 (6.7%) | 41 (5.2%) |

#### Alternative definition of HbA1c-based glycemic status definition

An alternative definition using HbA1c alone (normal < 5.7%, prediabetic 5.7–6.4%, diabetic > 6.4%) was included as a sensitivity analysis (Supplementary Materials Section F.2 & F.3).

#### Uncorrected τCMM mean lifetime map

We computed τCMM mean lifetime map without background- and time-offset-corrections as part of the sensitivity analysis (Supplementary Materials Section F.1 & F.3).

## 3. Results

### 3.1 Cohort Characteristics

The final dataset included 1,783 participants with available HbA1c measurements and a total of 6,912 FLIO scans (Table 1). Under the diagnosis- and treatment-aware glycemic group definition, 425 participants were normal, 569 prediabetic, and 789 diabetic (Table 1). The cohort had a mean age of 60.5 ± 11.2 years, with diabetic participants older on average than normal participants (61.6 ± 10.9 vs. 58.4 ± 11.4 years). Mean HbA1c is higher in the diabetc than the normal group (6.74 ± 1.36 vs. 5.35 ± 0.22). Mean corrected τCMM was longer in the diabetic than the normal group in both the SSC (5,462.4 ± 1,256.2 vs. 5,190.7 ± 1,019.4 ps) and the LSC (3,811.0 ± 630.4 vs. 3,688.5 ± 500.5 ps). Ocular and systemic comorbidities are more prevalent in the diabetic group than in the normal group.

### 3.2 Spatial τCMM Mean Lifetime Maps

Group-averaged mean lifetime maps revealed distinct spatial patterns across glycemic groups (Figure 2). Quantitative comparison of the mean τCMM within the 20 × 20-pixel central window is shown in Figure 4. In the SSC, mean participant-level τCMM increased from 4,681 ± 1,437 ps in the normal group to 4,953 ± 1,573 ps in prediabetes and 5,039 ± 1,639 ps in diabetes, and the three groups differed overall (one-way ANOVA F = 7.22, p < 0.001). Pairwise comparisons localized this difference to the contrast between normal glycemia and the two impaired groups: normal versus prediabetes (Bonferroni-adjusted p = 0.014) and normal versus diabetes (p < 0.001) were significant, whereas prediabetes versus diabetes was not. In the LSC, group means were 3,562 ± 821 ps, 3,737 ± 943 ps, and 3,757 ± 890 ps for normal, prediabetes, and diabetes, respectively (F = 7.15, p < 0.001; normal versus prediabetes p = 0.006 and normal versus diabetes p < 0.001). After adjustment for age, the SSC difference remained significant (ANCOVA p = 0.017), whereas the LSC difference did not (p = 0.16), indicating that the LSC contrast is largely explained by the older age of the impaired-glycemia groups.

To assess whether these findings depended on the choice of central-window size, we repeated the comparisons across windows from 10 × 10 to 190 × 190 pixels (Supplementary Materials Figures A3 and A4). Both the SSC and LSC differences were significant at every evaluated window size (ANOVA p < 0.002 throughout), with normal versus diabetes significant at every size (Bonferroni-adjusted p < 0.002) and prediabetes versus diabetes at none. The consistent separation of group-averaged maps across both spectral channels and all window sizes, with the greatest contrast between normal and diabetes, suggests that τCMM-derived FLIO representations capture retinal patterns associated with diabetes status, although age explains part of the group-mean difference in the LSC.

### 3.3. Model Performance on T2DM Classification

Model performance for 3-class classification is summarized in Table 2. Across all models, 3-class classification performance was modest. Among the FLIO-only models, the CNN achieved the highest AUROC (0.60 ± 0.04), followed by ResNet-18 (0.57 ± 0.04) and XGBoost (0.57 ± 0.03). The age-only classifier performed close to chance (AUROC 0.51 ± 0.03). Adding age as an input did not improve discrimination (AUROC 0.59 ± 0.02 for CNN + age, 0.55 ± 0.04 for ResNet-18 + age, and 0.57 ± 0.01 for XGBoost + age). The CNN also achieved the best or near-best performance across the remaining evaluation metrics.

**Table 2.**
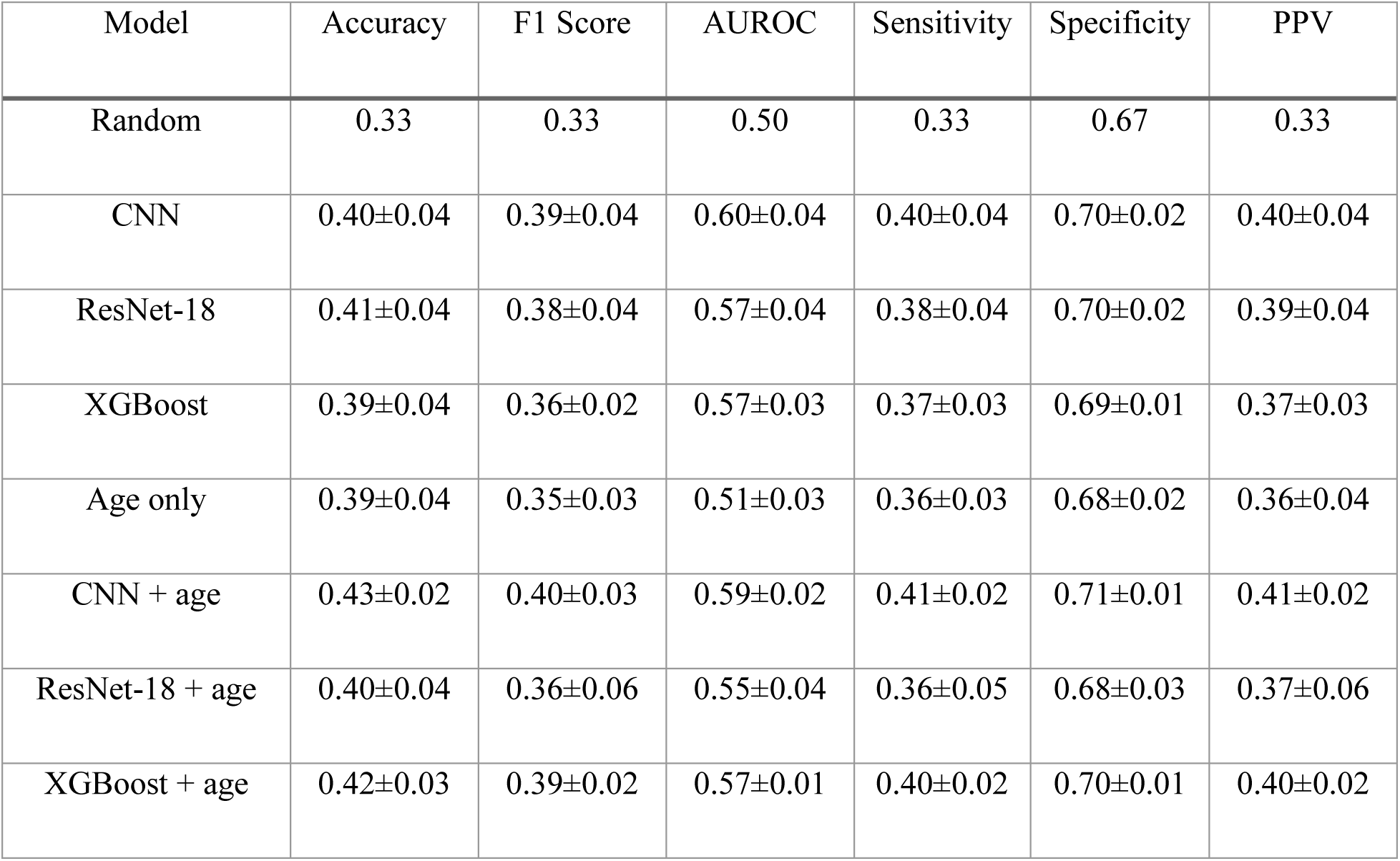
Model performance for 3-class classification. All models are participant-level with laterality canonicalization (right-eye maps mirrored to left-eye orientation). Age only = XGBoost trained on age alone; + age = the same model with age as an additional input (CNN and ResNet-18: age concatenated to the pooled image embedding; XGBoost: age added as a feature). Values are mean ± standard deviation across 5 outer test folds of nested cross-validation. AUROC = area under the receiver operating characteristic curve; CNN = convolutional neural network; PPV = positive predictive value; Random = uninformed random prediction.

Confusion matrices and ROC curves for FLIO-only and FLIO + age models are shown in Figure 5. All models showed substantial overlap between classes. Prediabetes was the least separable class (per-class AUROC 0.52–0.55 across the FLIO-only models), whereas normal and diabetic participants were discriminated somewhat better (AUROC 0.57–0.62). The CNN correctly classified the largest share of normal participants (50%), whereas ResNet-18 and XGBoost predicted the diabetes class more often, identifying more diabetic participants (55% and 48%, vs. 45% for the CNN).

**Figure 5.**
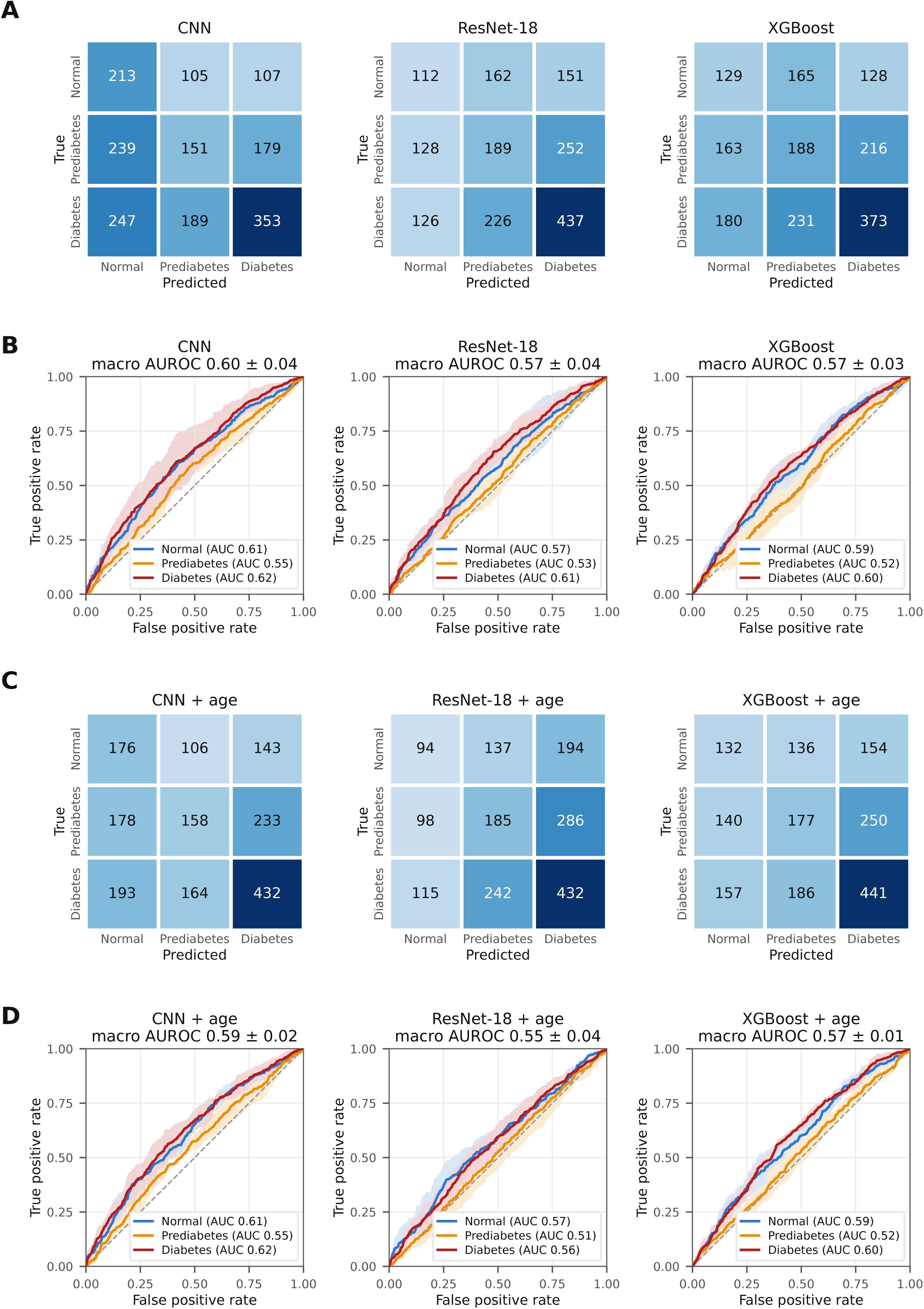
Three-class classification performance using FLIO alone and FLIO combined with age. (A) Confusion matrices for the CNN, ResNet-18, and XGBoost models using FLIO alone, pooled across the five outer test folds of nested cross-validation. Cell values are participant counts. (B) Per-class receiver operating characteristic (ROC) curves for each model, averaged across the outer test folds, with shaded regions denoting fold-to-fold standard deviation; per-class AUROCs are given in the legends and the macro-averaged AUROC (mean ± standard deviation across folds) in the panel titles. (C, D) Corresponding confusion matrices and ROC curves for the same models with age as an additional input. AUROC = area under the receiver operating characteristic curve; CNN = convolutional neural network.

Aggregated Grad-CAM saliency maps are shown in Figure 6. Attention magnitude varied primarily by true class rather than by predicted class. In participants with diabetes, Grad-CAM for the diabetes class was high across the macula, including the fovea, perifoveal ring, and region adjacent to the optic disc. In contrast, among normal and prediabetic participants. Within each true class, saliency patterns were broadly similar across the three predicted classes, indicating that the aggregated maps track characteristics of the input class more than the basis of individual classification decisions. Because Grad-CAM displays only positive contributions, this pattern suggests that the CNN responds to the presence of diffuse macular τCMM changes in diabetes, whereas normal and prediabetic predictions reflect their absence. High values along the image border reflect convolution padding at the edge of the field.

**Figure 6.**
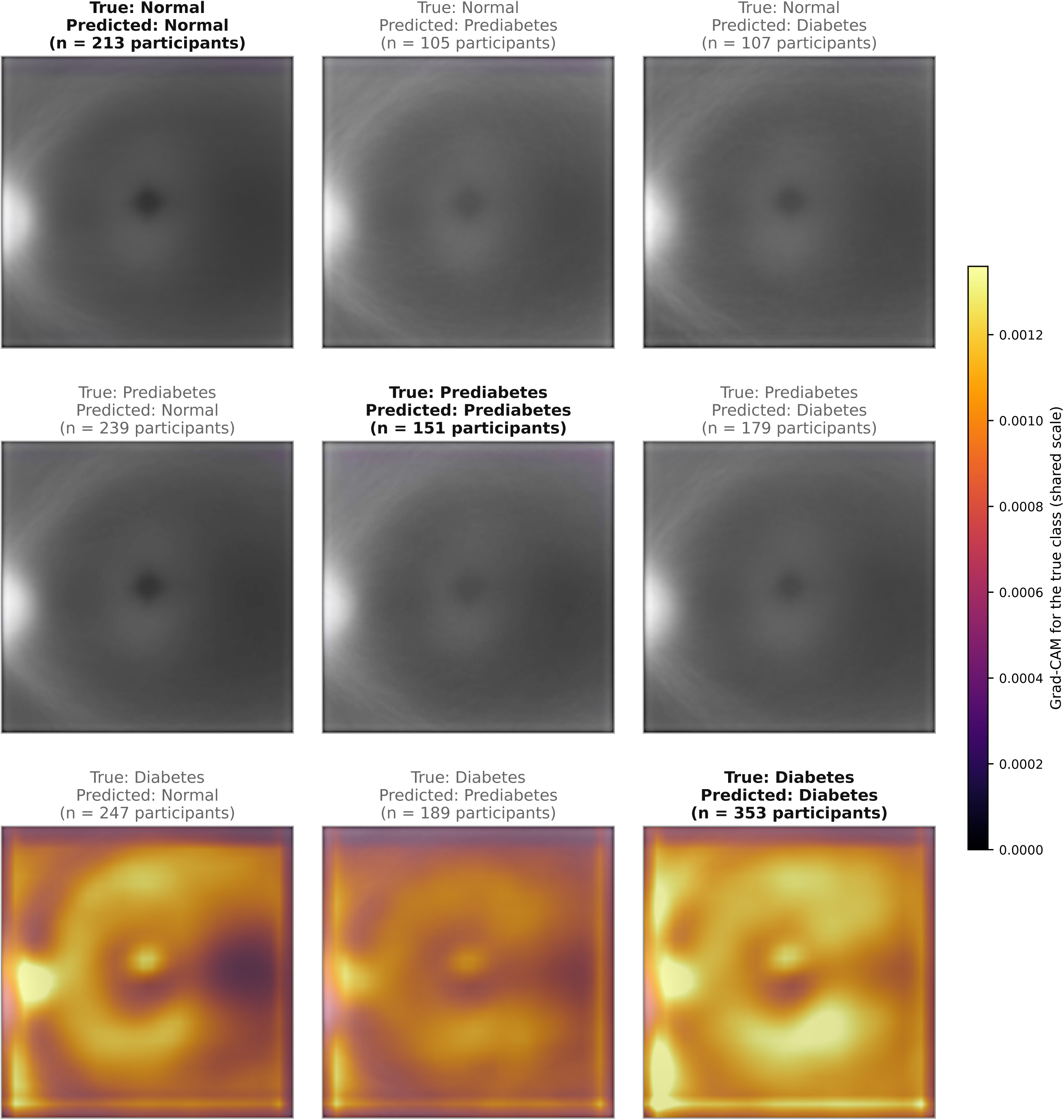
Aggregated Grad-CAM saliency maps of the CNN model using FLIO alone by confusion-matrix cell. Rows correspond to the true class and columns to the predicted class; the number of participants in each cell is shown above it, and correctly classified cells have bold titles. For each participant, Grad-CAM for the true class was computed at the final convolutional layer of the CNN from the outer fold in which the participant was held out; per-eye maps (left-eye orientation) were averaged within each cell and overlaid on the mean SSC input. All cells share one color scale; warmer, more opaque colors indicate a greater positive contribution to the true-class score. SSC = short spectral channel. CNN = convolutional neural network; Grad-CAM = Gradient-weighted Class Activation Mapping.

### 3.4 Binary Classification of Glycemic Status

As the prediabetes class is hard to distinguish from neighboring classes, we additionally evaluated two binary scenarios Normal vs. Impaired (ties the prediabetes and diabetes class as one impaired class, n = 1,783) and Normal vs. Diabetes (dropped prediabetes class, n = 1,214). Table 3 reports the participant-level binary results.

**Table 3.**
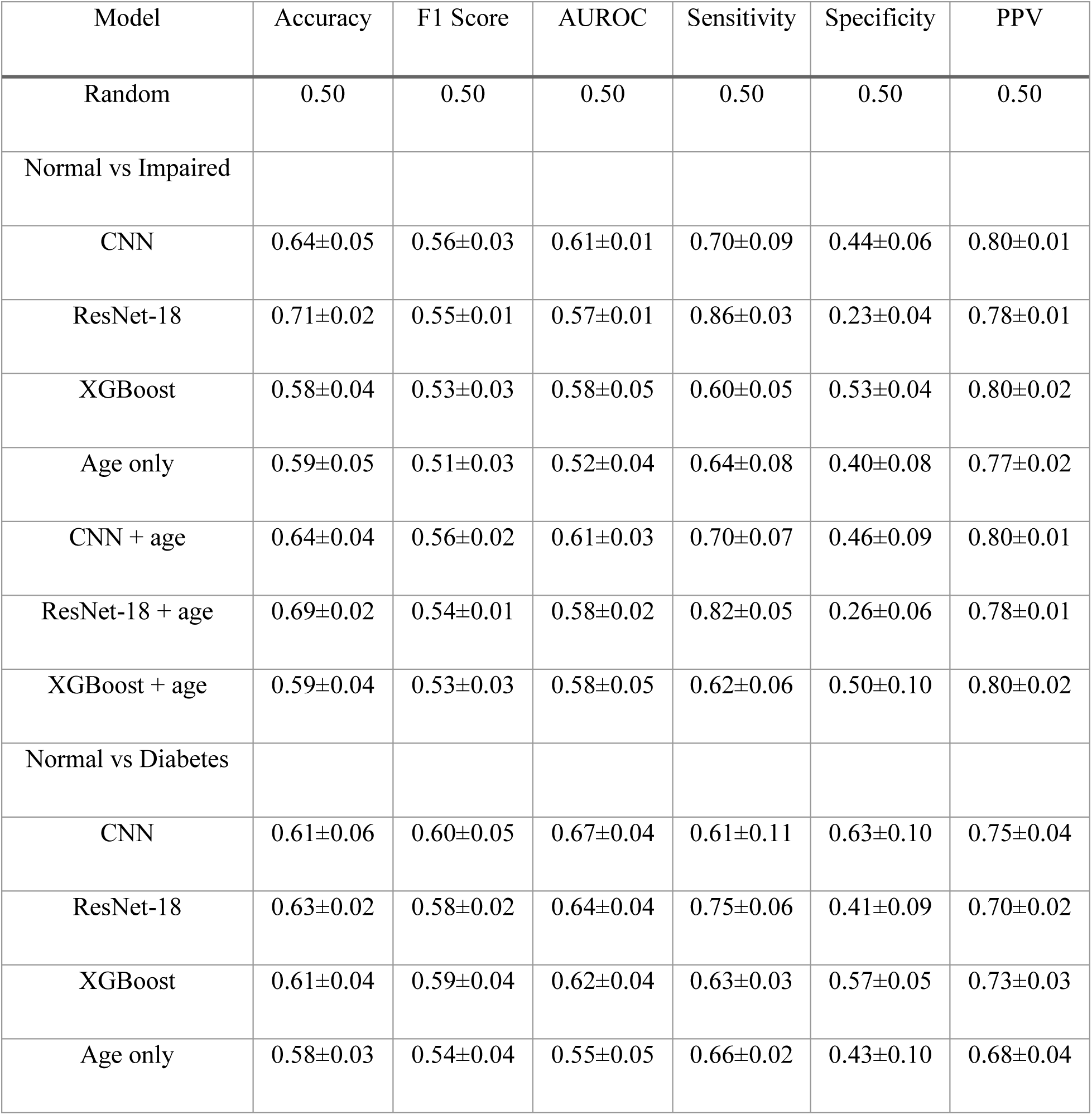
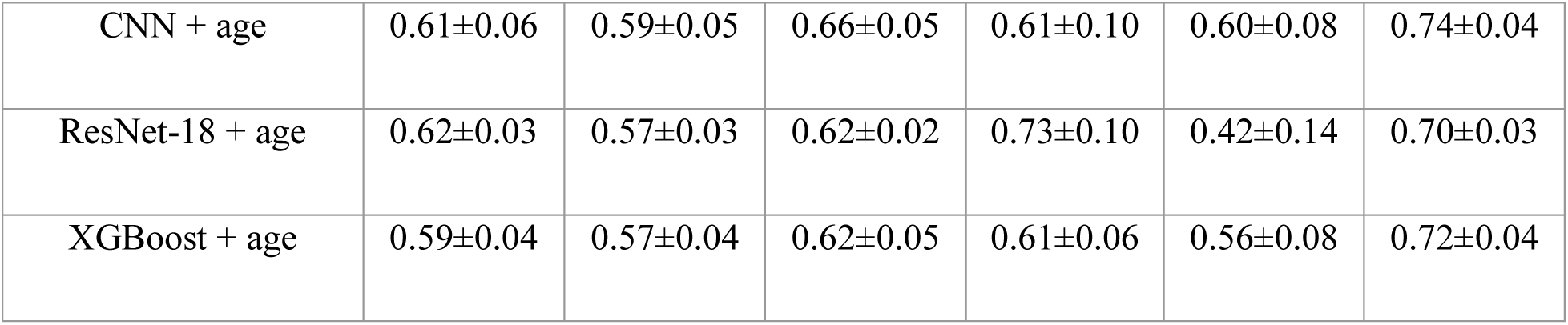
Model performance for binary classification tasks. For Normal vs. Impaired, prediabetic and diabetic participants were merged into a single Impaired class. For Normal vs. Diabetes, prediabetic participants were excluded. Age only = XGBoost trained on age alone; + age = the same model with age as an additional input. Values are mean ± standard deviation across 5 outer test folds of nested cross-validation. AUROC = area under the receiver operating characteristic curve; CNN = convolutional neural network; PPV = positive predictive value; Random = uninformed random prediction.

| Model | Accuracy | F1 Score | AUROC | Sensitivity | Specificity | PPV |
|---|---|---|---|---|---|---|
| Random | 0.50 | 0.50 | 0.50 | 0.50 | 0.50 | 0.50 |
| Normal vs Impaired |  |  |  |  |  |  |
| CNN | 0.64±0.05 | 0.56±0.03 | 0.61±0.01 | 0.70±0.09 | 0.44±0.06 | 0.80±0.01 |
| ResNet-18 | 0.71±0.02 | 0.55±0.01 | 0.57±0.01 | 0.86±0.03 | 0.23±0.04 | 0.78±0.01 |
| XGBoost | 0.58±0.04 | 0.53±0.03 | 0.58±0.05 | 0.60±0.05 | 0.53±0.04 | 0.80±0.02 |
| Age only | 0.59±0.05 | 0.51±0.03 | 0.52±0.04 | 0.64±0.08 | 0.40±0.08 | 0.77±0.02 |
| CNN + age | 0.64±0.04 | 0.56±0.02 | 0.61±0.03 | 0.70±0.07 | 0.46±0.09 | 0.80±0.01 |
| ResNet-18 + age | 0.69±0.02 | 0.54±0.01 | 0.58±0.02 | 0.82±0.05 | 0.26±0.06 | 0.78±0.01 |
| XGBoost + age | 0.59±0.04 | 0.53±0.03 | 0.58±0.05 | 0.62±0.06 | 0.50±0.10 | 0.80±0.02 |
| Normal vs Diabetes |  |  |  |  |  |  |
| CNN | 0.61±0.06 | 0.60±0.05 | 0.67±0.04 | 0.61±0.11 | 0.63±0.10 | 0.75±0.04 |
| ResNet-18 | 0.63±0.02 | 0.58±0.02 | 0.64±0.04 | 0.75±0.06 | 0.41±0.09 | 0.70±0.02 |
| XGBoost | 0.61±0.04 | 0.59±0.04 | 0.62±0.04 | 0.63±0.03 | 0.57±0.05 | 0.73±0.03 |
| Age only | 0.58±0.03 | 0.54±0.04 | 0.55±0.05 | 0.66±0.02 | 0.43±0.10 | 0.68±0.04 |
| CNN + age | 0.61±0.06 | 0.59±0.05 | 0.66±0.05 | 0.61±0.10 | 0.60±0.08 | 0.74±0.04 |
| ResNet-18 + age | 0.62±0.03 | 0.57±0.03 | 0.62±0.02 | 0.73±0.10 | 0.42±0.14 | 0.70±0.03 |
| XGBoost + age | 0.59±0.04 | 0.57±0.04 | 0.62±0.05 | 0.61±0.06 | 0.56±0.08 | 0.72±0.04 |

The Normal vs. Diabetes task yielded the best discrimination for every model. For Normal vs. Impaired, the CNN achieved AUROC 0.61 ± 0.01, ResNet-18 0.57 ± 0.01, and XGBoost 0.58 ± 0.05. For Normal vs. Diabetes, the CNN achieved the highest performance of any configuration in the primary analysis (AUROC 0.67 ± 0.04; F1 0.60 ± 0.05), followed by ResNet-18 (0.64 ± 0.04) and XGBoost (0.62 ± 0.04). The age-only classifier reached AUROC 0.52 ± 0.04 (Normal vs. Impaired) and 0.55 ± 0.05 (Normal vs. Diabetes), and adding age did not improve the FLIO models in binary classifications, consistent the findings from three-class classification (Table 3).

### 3.5 Sensitivity Analyses

#### Comorbidity exclusions

Excluding participants with ocular comorbidities generally preserved the *τ*CMM differences across both spectral channels, with and without age adjustment, except cataract. Excluding participants with cataract increased the between-group contrast in both the SSC and LSC, consistent with lens opacity and lens fluorescence contributing noise to the retinal signal. In contrast, excluding participants with systemic conditions generally changed the significant SSC group differences to nonsignificant, except after exclusion of participants with kidney disease. The age-adjusted LSC difference was not significant in the full cohort and remained nonsignificant after exclusion of systemic conditions (Supplementary Table A2).

We further assessed the impact of ocular comorbidities on classifying glycemic status. Ocular comorbidities led to small changes on the classification of glycemic status, with AUROC ranging from −0.04 to +0.03. Model ranking is preserved with CNN having the highest 3-class AUROC. (Supplementary Table A3).

#### Alternative glycemic label and τCMM map definitions

Results were similar under HbA1c-only labels and with uncorrected τCMM maps (Supplementary Section F, Tables A4–A10 and Figures A6–A17).

The corrected τCMM slightly weakens the SSC contrast. With the primary glycemic labels, the age-adjusted SSC group difference was weaker on the corrected (ANCOVA p = 0.017) than the uncorrected maps (ANCOVA p=0.003). Similar weakening effect was observed for the HbA1c-only glycemic label. The age-adjusted LSC group difference remains unsignificant across all four label-map combinations (Supplementary Table A4).

Classification performance of glycemic status was essentially unchanged by the label and the τCMM corrections (Supplementary Table A5-A10). Across all four label-map combinations, three-class AUROCs ranged from 0.567 to 0.596 for the CNN, 0.528 to 0.568 for ResNet-18, and 0.537 to 0.576 for XGBoost. Between corrected and uncorrected maps, AUROC differences ranged from −0.026 to +0.036 across all models and tasks. The models therefore rely on relative spatial patterns rather than absolute scale of τCMM. Between diagnosis- and treatment-aware and HbA1c-only labels, AUROC differences ranged from −0.039 to +0.059 (none significant after Bonferroni correction).

## 4. Discussion

This study shows that CMM-based FLIO mean lifetime maps carry a measurable signal related to glycemic status. Group-averaged τCMM lifetime differed significantly across glycemic groups in the SSC, and the difference persisted after age adjustment. All three FLIO models outperformed an age-only classifier, suggesting that the FLIO signal is not simply a proxy for age, although individual-level discrimination of glycemic status remained modest. By computing τCMM maps, we reduced complex spatiotemporal photon-counting data to interpretable two-dimensional representations, enabling the application of standard image-based deep learning models for glycemic classification.

The τCMM values reported in this study should not be interpreted as equivalent to conventional FLIO *τ_m_* values derived from multi-exponential fitting. First, τCMM was computed without instrument response function (IRF) deconvolution: the timing-offset correction removes the electronic delay but not the effect of the finite duration of the excitation pulse,^20^ and even after background and timing-offset correction, central-window values (approximately 4.7–5.0 ns in the SSC and 3.6–3.8 ns in the LSC) remain much longer than fitted *τ_m_* values reported in the FLIO literature. Second, uniform background counts inflate τCMM in dim pixels, which produced apparently long foveal values in uncorrected LSC maps (Supplementary Materials Figure A6); background correction removed most of this artifact while leaving classification performance essentially unchanged (Table 2&3 vs Table A5&A6), indicating that the models rely on relative spatial patterns rather than absolute scale. Interpretation should therefore focus on relative spatial and group differences rather than absolute magnitude.

Visualization of aggregated τCMM maps revealed longer τCMM in prediabetic and, more markedly, diabetic participants across most of the macula, with the largest differences at the fovea (Figures 2 and 3). These differences may reflect metabolic alterations in retinal fluorophores associated with chronic hyperglycemia; they persisted after age adjustment and after excluding participants with self-reported diabetic retinopathy. That the models outperformed an age-only classifier suggests that FLIO captures diabetes-related retinal signals beyond those attributable to age.

Discrimination in our study (best AUROC 0.67, CNN, normal versus diabetes) was also lower than that reported by Schmidt *et al.* for classifying nonproliferative diabetic retinopathy versus healthy controls (AUROC 0.865),^6^ likely because only 8.9% of our diabetic participants reported DR, so that our diabetes group largely represents an earlier disease stage, and because our estimates were obtained from patients in the evaluation folds rather than from a logistic-regression model fitted to a small case–control sample (33 cases and 28 controls). In contrast to the classification task, regression models predicting continuous HbA1c levels performed poorly and exhibited prediction collapse toward the population mean (Supplementary Materials Figure A5). This phenomenon suggests that while FLIO captures metabolic differences between glycemic states, the signal may be insufficient to recover fine-grained variation in HbA1c. HbA1c reflects systemic glycemic exposure rather than localized retinal metabolism, which may limit the predictive capacity of retinal imaging alone.

Overall, these results suggest that while τCMM maps contain information related to glycemic states, the separation between classes is subtle, leading to overlapping predictions across categories. The consistent performance across different model types further indicates that the limitation may stem from the intrinsic signal in the data rather than model capacity.

These findings have important implications for the use of FLIO in clinical applications. While FLIO may serve as a promising noninvasive screening tool for identifying metabolic abnormalities, it is unlikely to replace blood-based HbA1c measurements for precise glycemic quantification.

Several limitations should be noted. First, this study used a single dataset, and external validation is needed to assess generalizability. Second, only FLIO-derived inputs were included; integrating FLIO with other imaging modalities such as OCT or fundus photography may further improve predictive performance. Third, outcome labels combined self-reported diagnosis and treatment and a single HbA1c measurement; diabetes duration was not available, and self-reported information may be imprecise. Fourth, diabetic macular edema status, panretinal or focal laser history, and lens status (phakic vs. pseudophakic) are not recorded in the AI-READI dataset (version 3) and could not be assessed; comorbidity information was limited to self-report. Fifth, no measured instrument response function is available for these acquisitions, precluding reliable multi-exponential reconvolution fitting; comparison of τCMM against fitted *τ_m_* on datasets with measured IRFs remains future work.

In summary, FLIO-derived τCMM maps contain spatial information related to diabetes status beyond age alone, but the signal is too weak to support accurate classification of glycemic states at the individual level. These findings delineate both the potential and the challenges of FLIO-based metabolic assessment.

## Data Availability

All data produced in the present study are available upon reasonable request to the authors

## Declaration of Generative AI and AI-assisted technologies in the writing process

During the preparation of this work the authors used Claude Opus 4.8 in order to edit and improve the readability and language of the manuscript. GPT-5.3 was additionally used to assist in developing the analytical pipeline. After using these tools/services, the authors reviewed and edited the content as needed and take full responsibility for the content of the publication.

## Meeting Presentation

None

## Abbreviations and Acronyms

AGE: advanced glycation end products
AI: artificial intelligence
AI-READI: Artificial Intelligence Ready and Equitable Atlas for Diabetes Insights
AMD: age-related macular degeneration
ANCOVA: analysis of covariance
ANOVA: analysis of variance
AUROC: area under the receiver operating characteristic curve
CMM: center-of-mass method
CNN: convolutional neural network
DR: diabetic retinopathy
ETDRS: Early Treatment Diabetic Retinopathy Study
FAD: flavin adenine dinucleotide
FAF: fundus autofluorescence
FLIM: fluorescence lifetime imaging
FLIO: fluorescence lifetime imaging ophthalmoscopy
Grad-CAM: Gradient-weighted Class Activation Mapping
HbA1c: hemoglobin A1c
IRF: instrument response function
LSC: long spectral channel
NPDR: non-proliferative diabetic retinopathy
OCT: optical coherence tomography
PPV: positive predictive value
ROC: receiver operating characteristic
SSC: short spectral channel
T2DM: type 2 diabetes mellitus
τCMM: center-of-mass mean lifetime

## Supplementary Materials

### A. Model training details

#### A.1 CNN and ResNet-18

The CNN and ResNet-18 models were trained with the Adam optimizer (learning rate 1 × 10⁻⁴, weight decay 1 × 10⁻⁴) for up to 50 epochs per fold with a batch size of 16. A ReduceLROnPlateau scheduler (factor 0.5, patience 3 epochs) reduced the learning rate when validation macro F1 plateaued, and the model state from the epoch with the highest validation macro F1 was retained as the final fold checkpoint. No gradient clipping or weight averaging was used.

#### A.2 XGBoost

The XGBoost baseline used the hist tree method throughout, with the inner cross-validation grid summarized in Table A1.

**Table A1.**
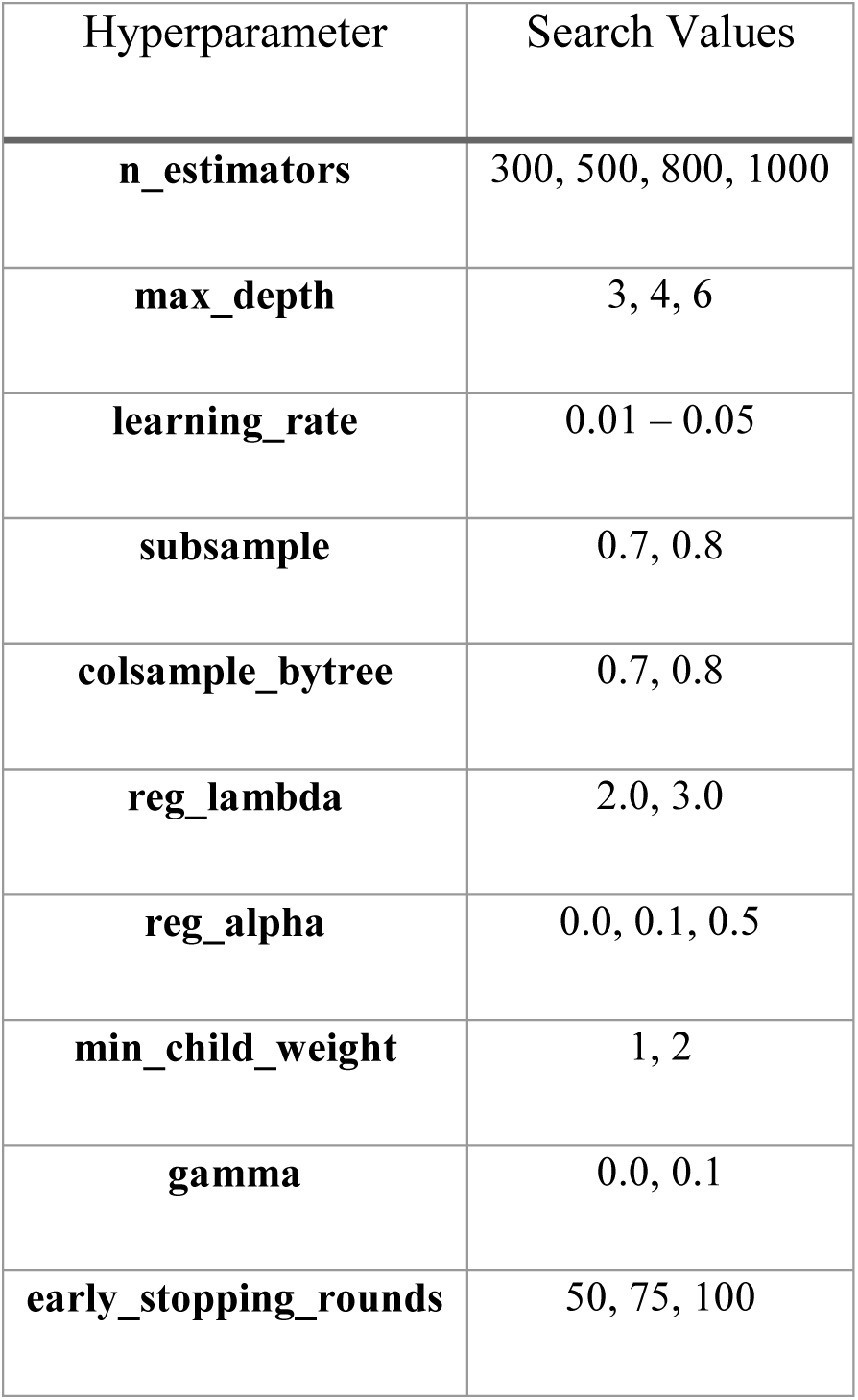
XGBoost hyperparameter grid searched via inner cross-validation.

The XGBoost hist tree method was used throughout. Hyperparameter names follow the XGBoost Python API conventions.

#### A.3 Class-balanced loss

The deep models were trained with class-weighted cross-entropy. Weights followed the standard scikit-learn formulation,

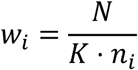

where *N* is the total number of training participants in the fold, *K* is the number of classes, and *nᵢ* is the count of class *i* in that fold. Weights were renormalized to unit mean before use. The same formulation was applied to the binary tasks with *K* = 2.

#### A.4 Random seeds and reproducibility

A global seed of 42 was used for the outer cross-validation. The inner cross-validation seeds were offset by the outer-fold index (seed = 42 + *k* for outer fold *k*) so that inner splits did not overlap across folds.

#### A.5 Participant-level aggregation

Per-eye encoder outputs (256-dimensional for CNN, 512-dimensional for ResNet-18) were aggregated to per-participant representations by mean pooling over each participant’s available eyes (1 or 2). For the participants with a single eye, the singleton embedding was used directly without zero-padding. Participant predictions were obtained from the classifier head applied to the pooled embedding. Right-eye maps were mirrored horizontally to the left-eye orientation before encoding (laterality canonicalization). In the FLIO + age models, age scaled as (age − 60)/10 was concatenated to the pooled embedding before the classifier head.

### B. Details about τCMM mean lifetime estimation

#### B.1 Uncorrected τCMM mean lifetime estimation

The mean lifetime at each pixel was estimated as:

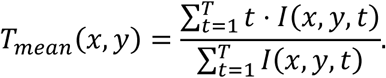

The mean lifetimes were converted from bin units to physical time using the frame time recorded in the FLIO metadata which specifies the temporal spacing between bins. The mean fluorescence lifetime at each pixel was computed as:

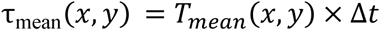

where Δ*t* denotes the frame time per bin.

This process yielded a two-dimensional spatial map of the mean fluorescence lifetime for each eye and spectral channel. The main difference between this uncorrected method and the corrected method presented in the main text is the lack of background- and time-offset-corrections.

#### B.2 Impact of Intensity Filtering on τCMM Mean-Lifetime Maps

We assessed the impact of the intensity filtering, which excludes pixels with fewer than 50 photons, in estimation of the τCMM mean lifetime maps. For the corrected τCMM mean lifetime maps, the median proportion of retained pixels was 99.84% for SSC (IQR, 99.44%–99.99%) and 100% for LSC (IQR, 99.87%–100%). Pixels were excluded in 142 SSC eyes (4.1%) and 84 LSC eyes (2.4%).

Excluded pixels were predominantly located at frame borders and in dim regions of a small number of low-quality scans. Therefore, the intensity threshold is unlikely to have systematically removed pixels from low-intensity anatomical structures or materially altered the spatial distribution of retinal lifetime signals.

### C. Sensitivity analysis of Center Region of Interest

To assess the spatial extent and robustness of group-level τCMM differences, we performed a sensitivity sweep across central-window sizes on the primary (corrected) maps with the diagnosis- and treatment-aware groups. For each window size from 10 to 190 pixels at 10-pixel intervals, we computed the per-eye mean τCMM within a square window centered on the image center, separately for the short and long spectral channels. The location of the primary 20 × 20-pixel window and of the ETDRS rings is shown in Figure A1.

For each window size, group-level differences were assessed using one-way ANOVA on per-participant means, with pairwise post-hoc comparisons performed using Welch’s t-test, Bonferroni-corrected for three pairwise contrasts (Normal vs. Prediabetes, Normal vs. Diabetes, Prediabetes vs. Diabetes). For visualization of the sweep, a group-aggregated short-spectral-channel τCMM map was constructed by averaging across all eyes (right eyes mirrored to left-eye orientation; Figure A2), with sweep-window outlines overlaid to indicate the spatial extent of each evaluated region. Both the SSC and LSC group differences were significant at every window size (ANOVA p < 0.002), normal versus diabetes was significant at every size (Bonferroni-adjusted p < 0.002), and prediabetes versus diabetes at none (Figures A3 and A4).

**Figure A1.**
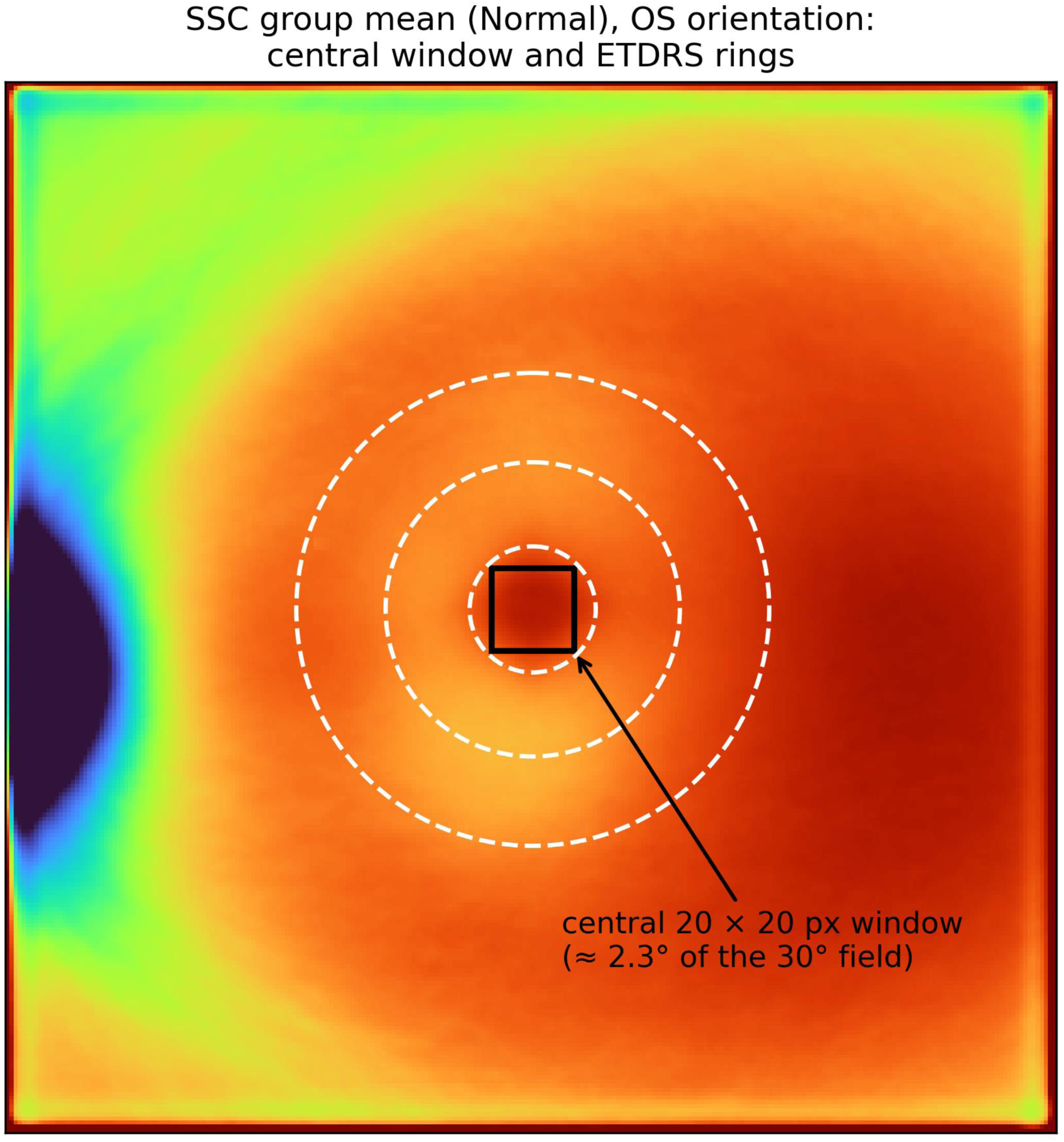
Location and scale of the analysis regions. The fixed 20 × 20-pixel central window used in the primary comparison and the ETDRS rings are overlaid on the group-mean SSC τCMM map of normal participants. The window is centered on the image, not on the fovea. ETDRS = Early Treatment Diabetic Retinopathy Study; SSC = short spectral channel; τCMM = center-of-mass mean lifetime.

**Figure A2.**
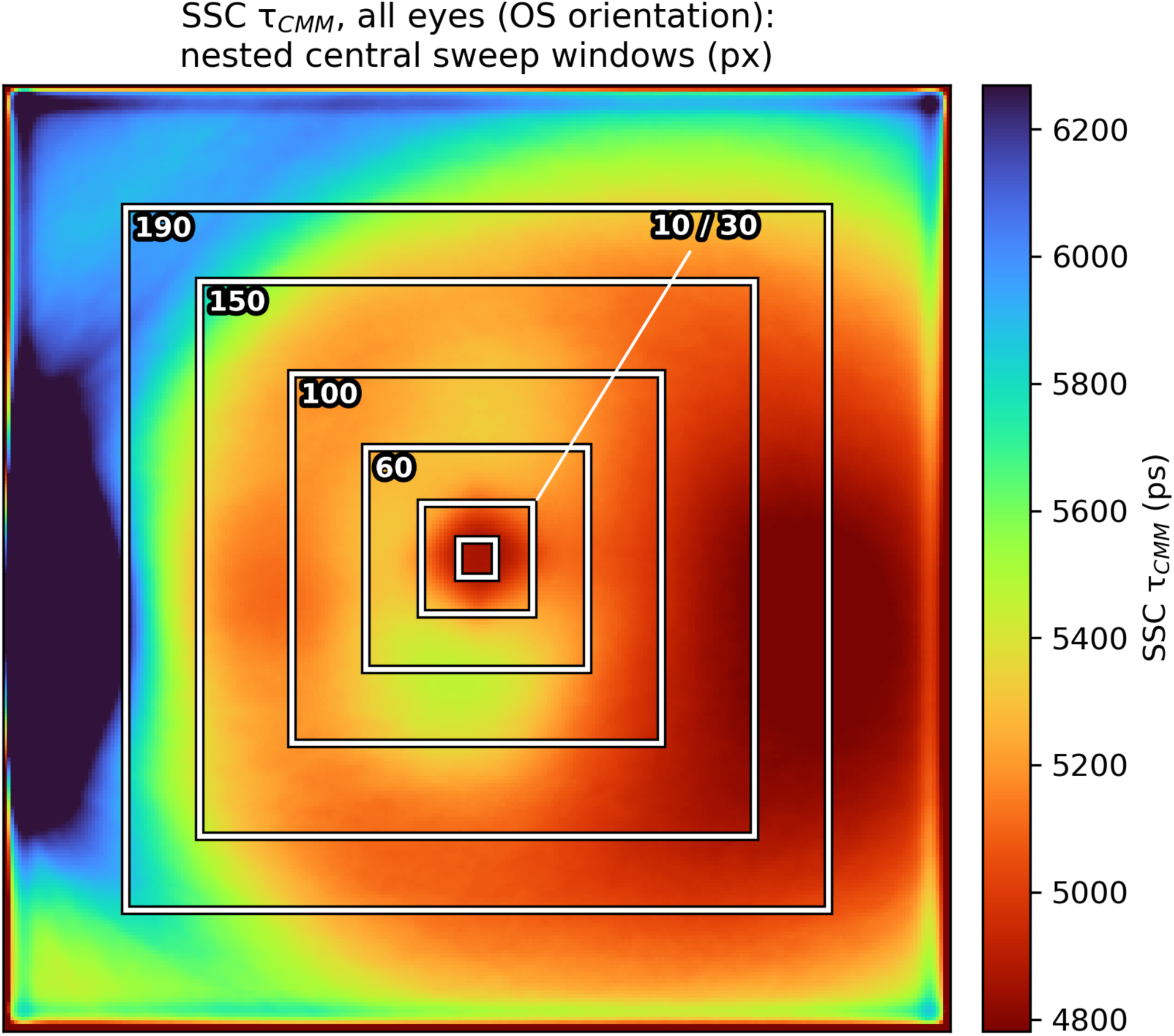
Aggregated short-spectral-channel (SSC) τCMM map (corrected maps, all eyes, left-eye orientation) with the sweep-window outlines overlaid. Nested square windows centered on the image indicate the sweep sizes (10 × 10, 30 × 30, 60 × 60, 100 × 100, 150 × 150, 190 × 190 pixels). Color bar shows SSC τCMM (ps).

**Figure A3.**
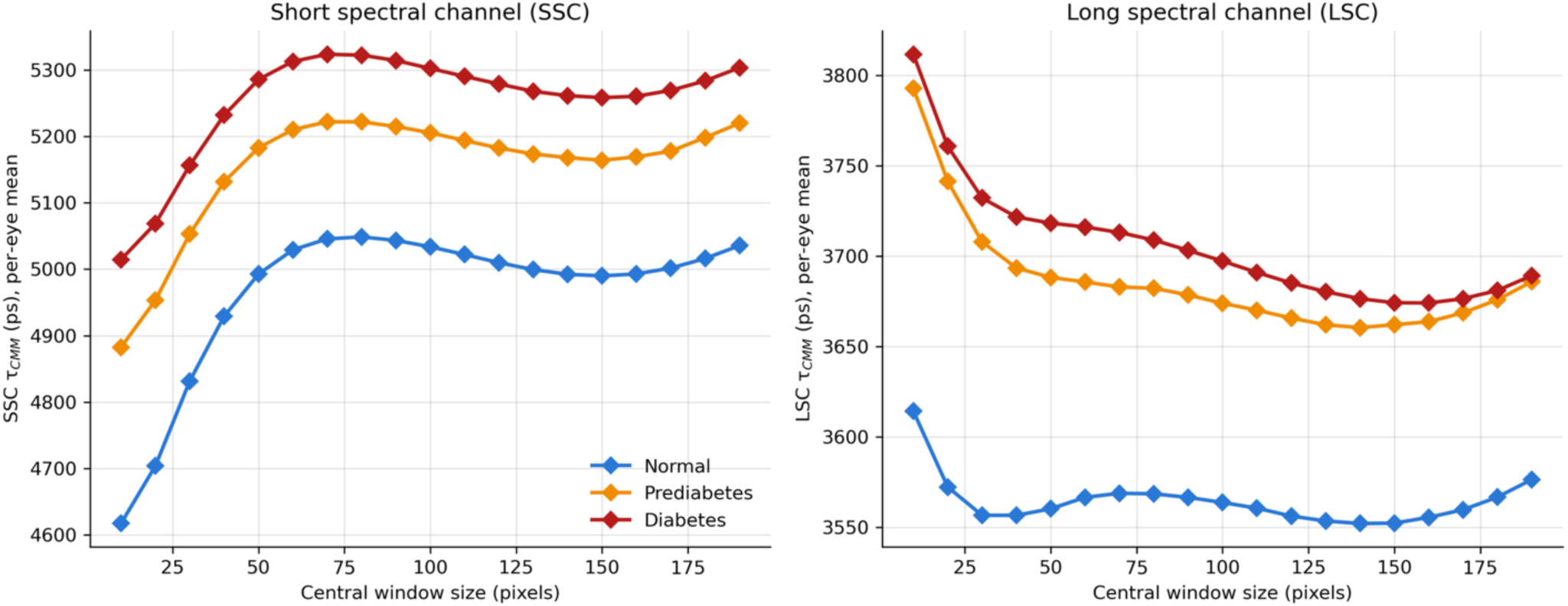
Per-group mean τCMM as a function of central-window size for the short spectral channel (SSC, left) and long spectral channel (LSC, right), on corrected maps with diagnosis- and treatment-aware groups. Each point is the per-eye mean τCMM averaged across the group at that window size; window sizes were swept from 10 to 190 pixels in 10-pixel steps. Color indicates glycemic group (Normal, blue; Prediabetes, orange; Diabetes, red). τCMM = center-of-mass mean lifetime; ps = picoseconds.

**Figure A4.**
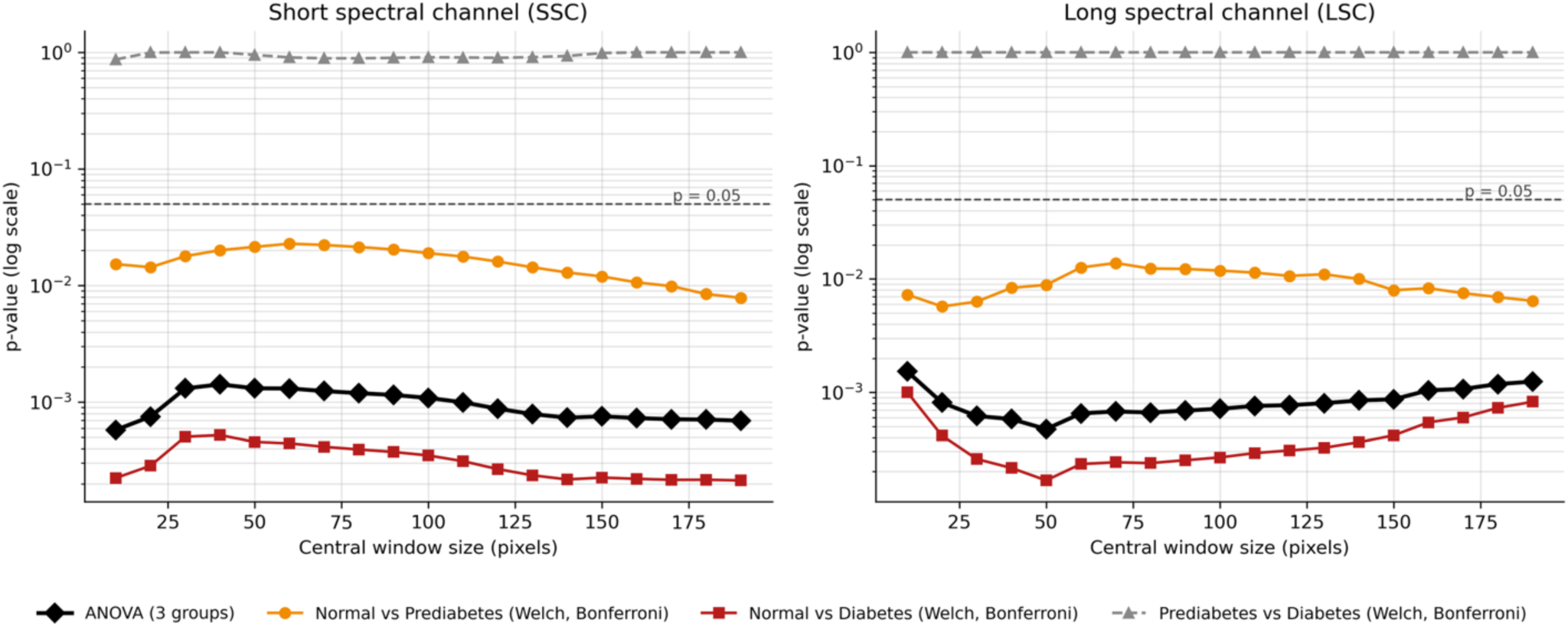
One-way analysis of variance (ANOVA) and pairwise Welch’s t-test p-values (log scale) as a function of central-window size for the short spectral channel (SSC, left) and long spectral channel (LSC, right), on corrected maps with diagnosis- and treatment-aware groups (participant-level means). Black diamonds: overall ANOVA p across the three groups. Orange circles, red squares, and gray triangles: pairwise Welch’s t-tests (normal vs. prediabetes, normal vs. diabetes, prediabetes vs. diabetes), Bonferroni-corrected across three contrasts. The dashed gray horizontal line indicates p = 0.05.

### D. HbA1c Prediction

#### D.1 Model training

To assess whether FLIO-derived features can capture continuous glycemic variation, we trained models to predict HbA1c values directly from τCMM maps. The same model architectures described for classification (CNN, ResNet18, and XGBoost) were used, with the primary modification being the replacement of the final classification layer with a regression output layer.

Models were trained using the same procedures as in the classification setting. For deep learning models, Smooth L1 loss was used for optimization, while XGBoost models were trained using regression objectives.

Model performance was assessed using mean absolute error (MAE). Agreement between predicted and observed HbA1c values was evaluated using Bland–Altman analysis.

#### D.2 Results

Regression models trained to predict continuous HbA1c values achieved a test mean absolute error of approximately 0.76. However, predicted values showed strong collapse toward the mean HbA1c level, indicating that the model failed to capture meaningful variation in HbA1c across patients. These regression analyses were performed with the eye-level models of the uncorrected maps.

**Figure A5.**
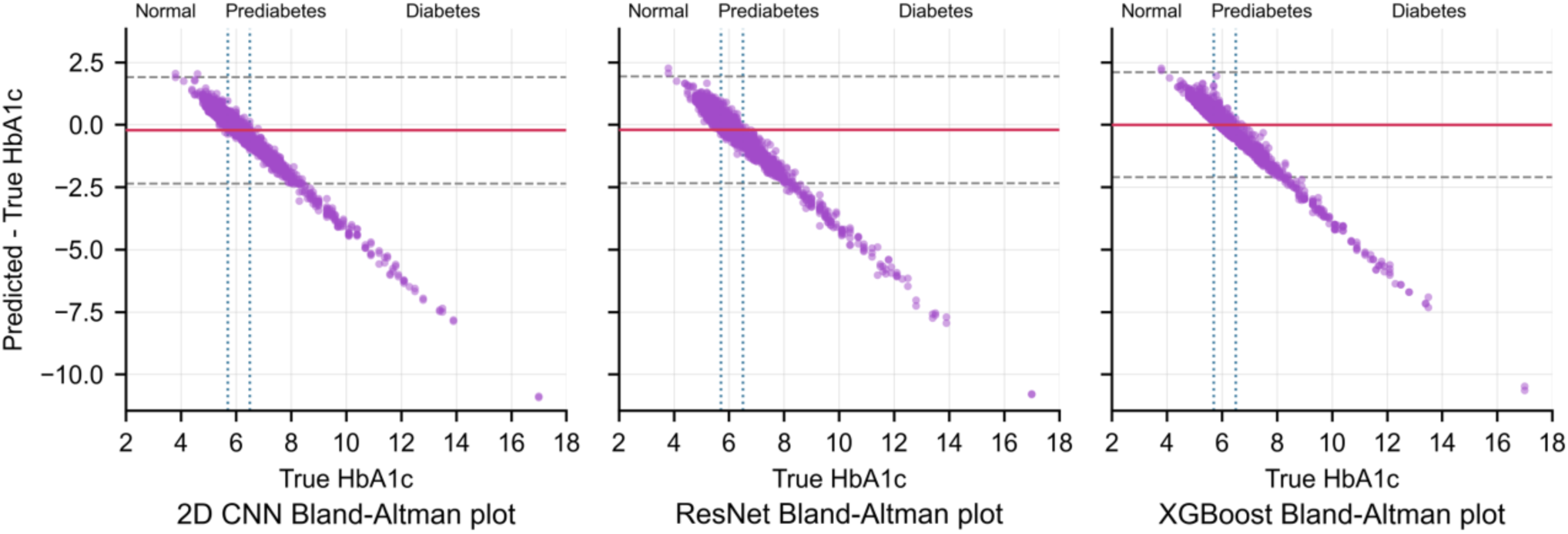
Bland-Altman plots comparing predicted versus true hemoglobin A1c (HbA1c) for the 2D CNN, ResNet-18, and XGBoost regression models. The x-axis shows true HbA1c (%); the y-axis shows the prediction error (predicted − true). The solid red line indicates the mean bias; dashed gray lines indicate the 95% limits of agreement (bias ± 1.96 × standard deviation). Vertical dotted lines mark the clinical HbA1c cutoffs (5.7% and 6.4%); Normal / Prediabetes / Diabetes labels above each panel indicate the corresponding glycemic ranges. CNN = convolutional neural network.

### E. Sensitivity analyses: comorbidities

On the primary labels and corrected maps, the central-window group comparison (Table A2) was repeated after excluding, in turn, participants with each self-reported ocular condition (diabetic retinopathy, cataract, glaucoma, or age-related macular degeneration) and each self-reported systemic condition (hypertension, high cholesterol, obesity, kidney disease, or cardiovascular disease), and after excluding participants with any ocular condition, any systemic condition, or any ocular or systemic condition. All three classifiers were retrained, under the same nested cross-validation protocol, after excluding each of the four ocular conditions and after excluding any of the four ocular conditions (Table A3).

**Table A2.**
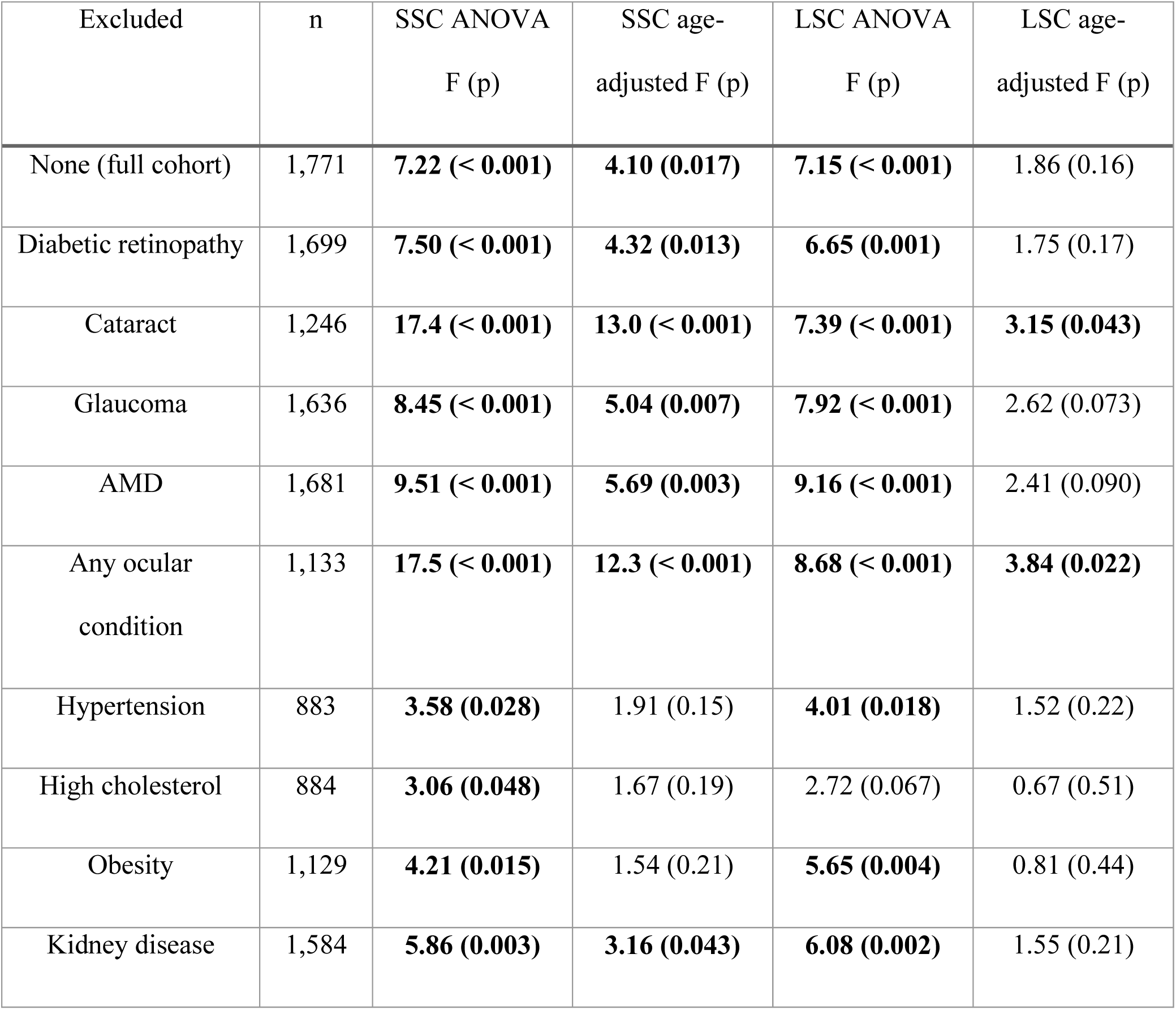
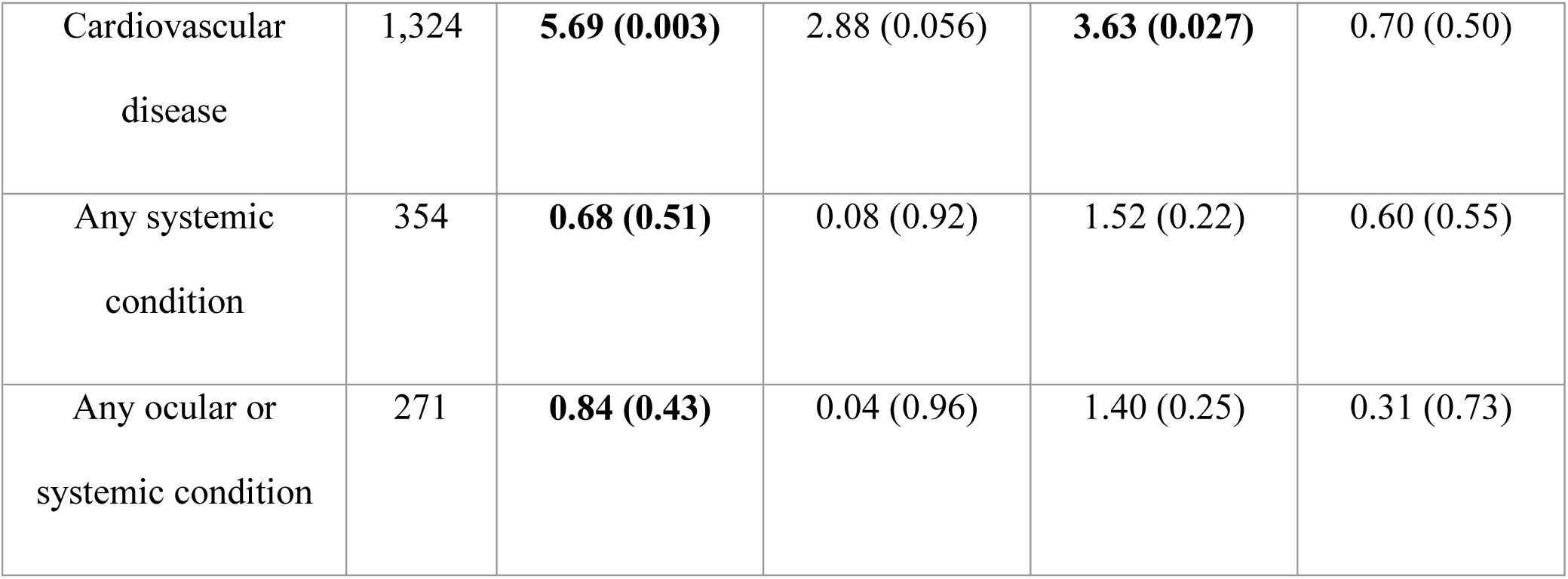
Central-window τCMM group comparison after excluding participants with self-reported ocular or systemic comorbidities (diagnosis- and treatment-aware labels; background- and timing-offset-corrected τCMM maps). n = participants with a valid SSC central-window value. Each single-condition row excludes participants reporting that condition. Any ocular condition = diabetic retinopathy, cataract, glaucoma, AMD, or retinal vascular occlusion (RVO); any systemic condition = hypertension, high cholesterol, obesity, kidney disease, or cardiovascular disease (heart attack, stroke, other heart issues, or circulation problems); any ocular or systemic condition = any of these ten conditions. All conditions are self-reported (Table 1). ANOVA F (p): one-way analysis of variance across the three glycemic groups; age-adjusted F (p): F-test of the group effect (2 df) in an analysis of covariance with age as covariate. *p* < 0.05 was considered statistically significant and bolded. AMD = age-related macular degeneration; LSC = long spectral channel; SSC = short spectral channel; τCMM = center-of-mass mean lifetime.

**Table A3.**
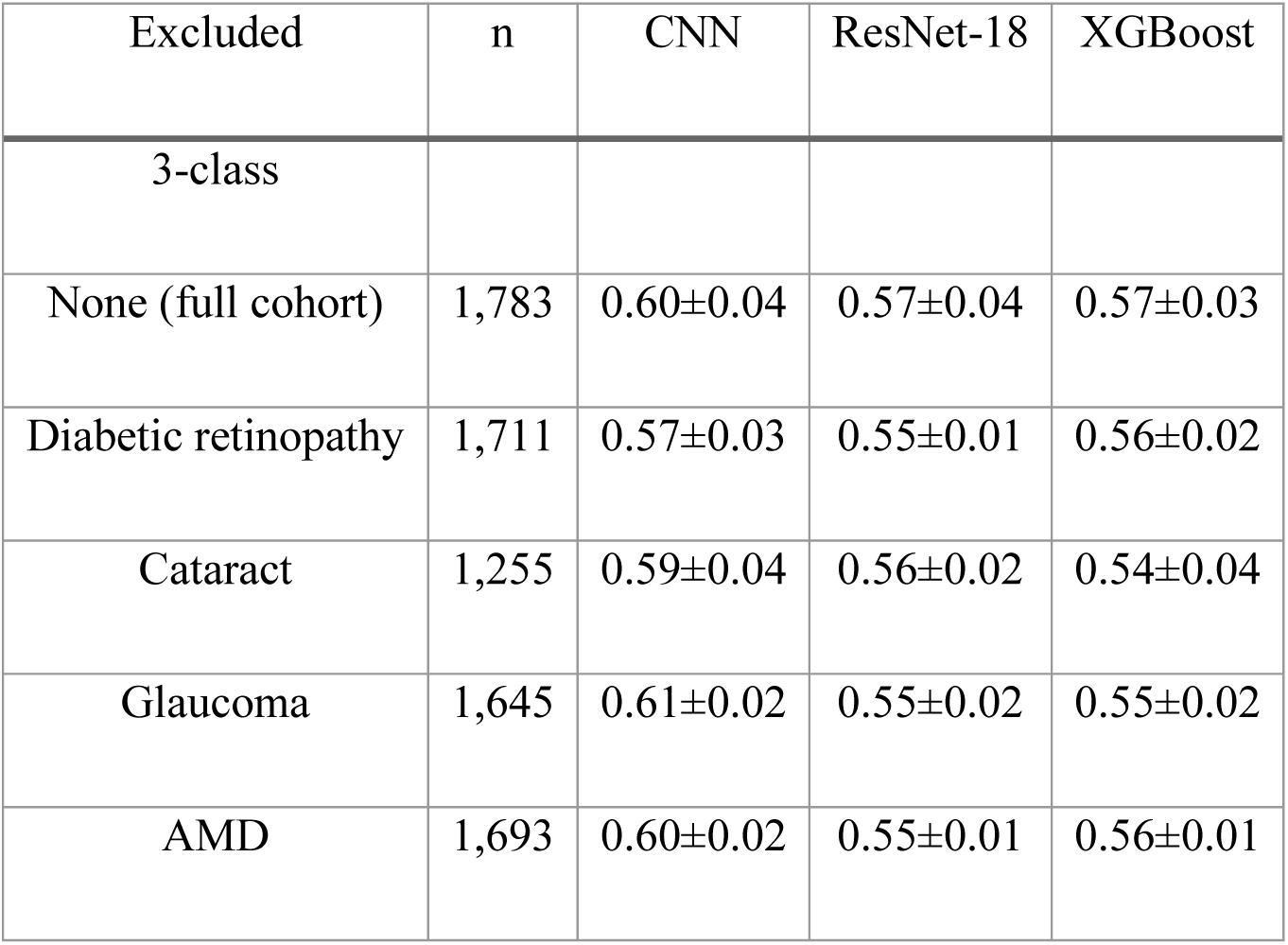
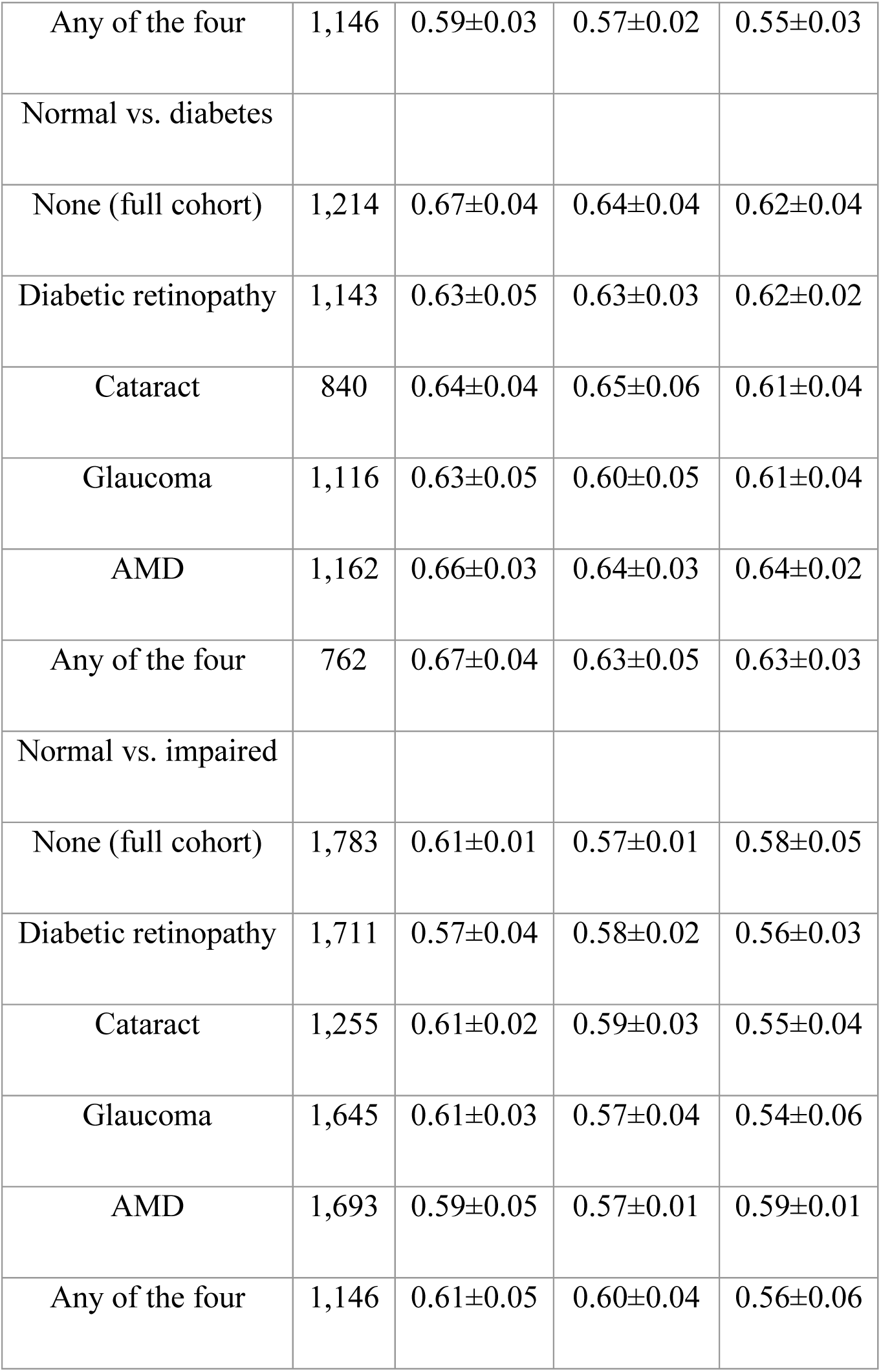
Classification performance (AUROC) after excluding participants with self-reported ocular comorbidities (diagnosis- and treatment-aware labels; background- and timing-offset-corrected τCMM maps). Each model was retrained on the exclusion cohort; values are mean ± standard deviation across the 5 outer test folds of nested cross-validation. n = participants evaluated by the CNN and ResNet-18; XGBoost was evaluated on the participants with complete ETDRS features (up to 10 fewer). Any of the four = diabetic retinopathy, cataract, glaucoma, or AMD; unlike the any-ocular-condition row of Table A2, participants reporting retinal vascular occlusion were not additionally excluded. AMD = age-related macular degeneration; AUROC = area under the receiver operating characteristic curve; CNN = convolutional neural network.

### F. Results under alternative label and map definitions

This section reports the complete results for the three alternative combinations of outcome label (diagnosis- and treatment-aware vs. HbA1c-only) and τCMM map (corrected vs. uncorrected); the primary combination (diagnosis- and treatment-aware labels, corrected maps) is reported in the main text. HbA1c-only labels assign normal (HbA1c < 5.7%), prediabetic (5.7–6.4%), and diabetic (> 6.4%) status from the measured HbA1c alone (671, 726, and 386 participants). Uncorrected maps are the center of mass of the raw photon-count histogram without background subtraction or timing-offset removal, as described in Supplementary Section B. All other procedures are identical to the primary analysis.

**Table A4.**
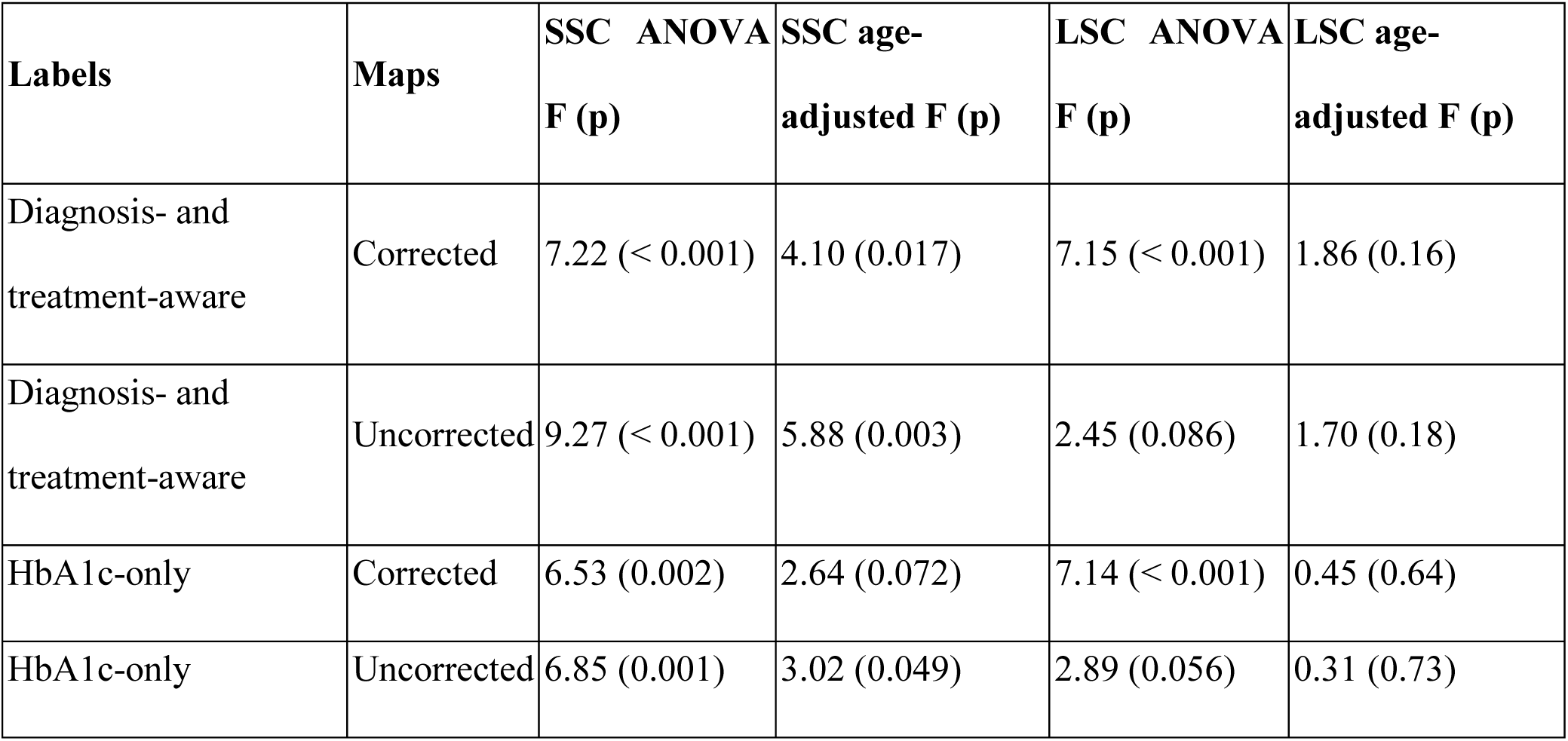
Central-window τCMM group comparison across the four label–map combinations.

#### F.1 Diagnosis- and treatment-aware labels, uncorrected maps

Central-window comparison: SSC ANOVA F = 9.27, p < 0.001 (age-adjusted p = 0.003); LSC ANOVA F = 2.45, p = 0.086 (age-adjusted p = 0.18) (Figures A6–A9).

**Table A5.**
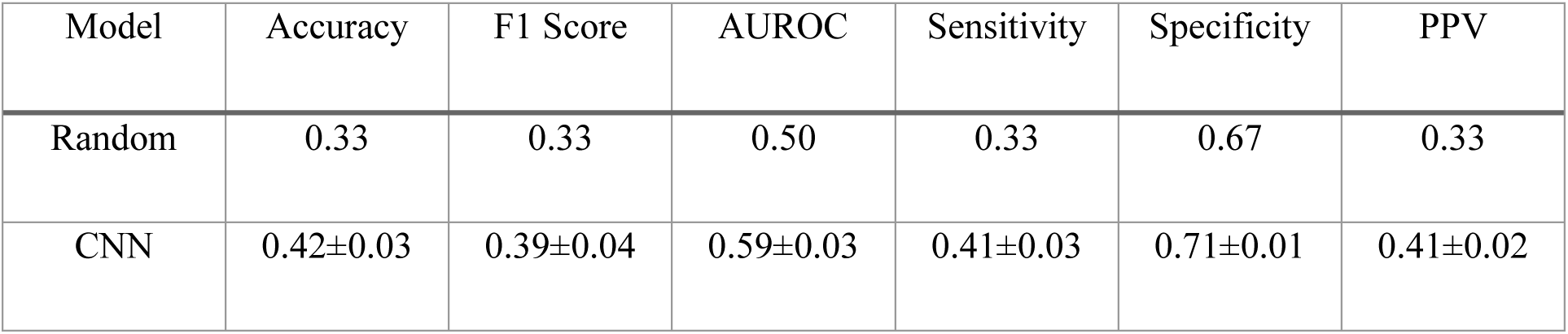
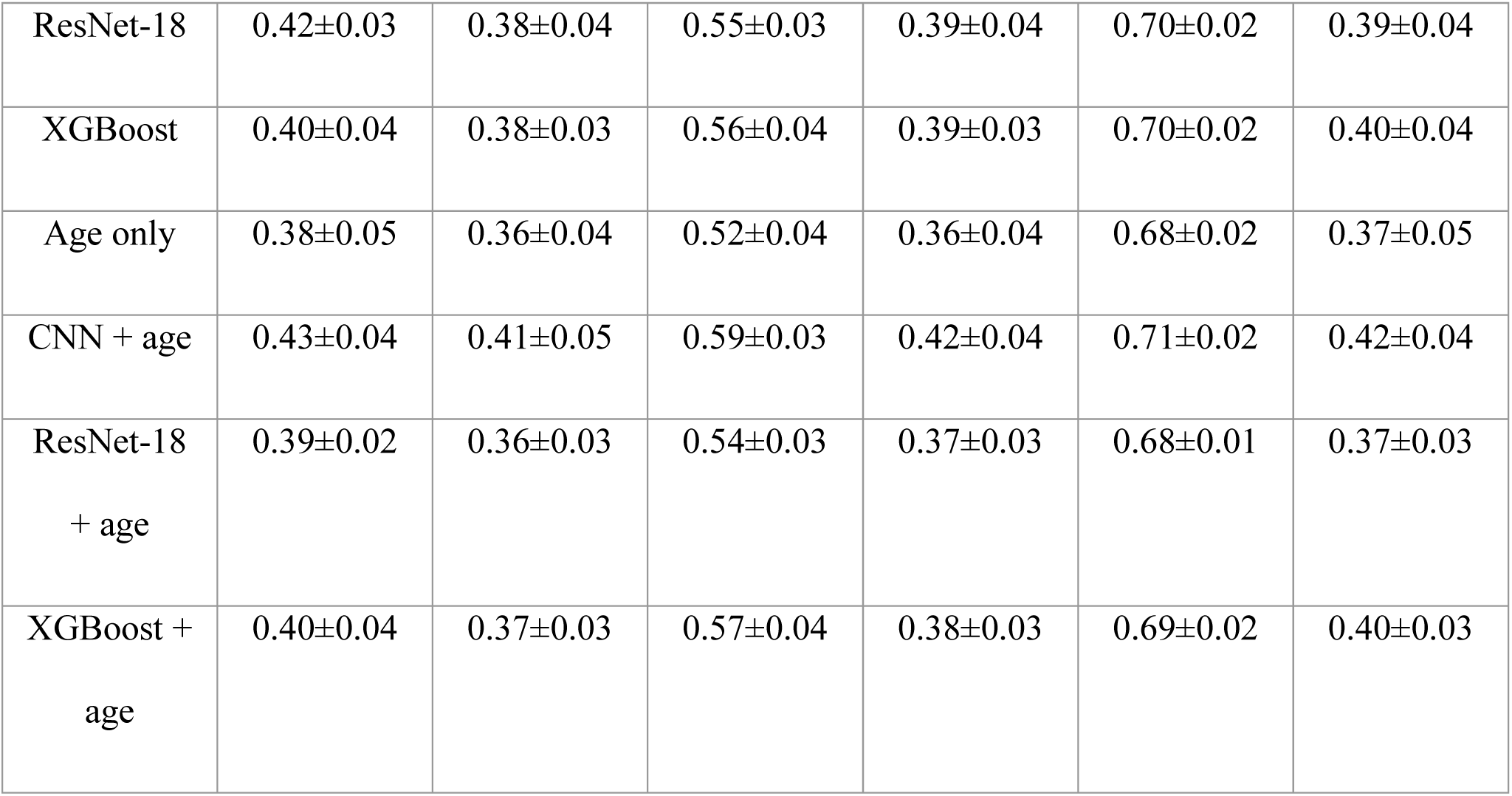
Model performance for 3-class classification, macro-averaged (diagnosis- and treatment-aware labels; uncorrected τCMM maps). Models and cohorts as in main-text Table 2; XGBoost and Age only were evaluated on the 1,774 participants with complete ETDRS features, CNN and ResNet-18 on all 1,783 participants. Values are mean ± standard deviation across 5 outer test folds of nested cross-validation. AUROC = area under the receiver operating characteristic curve; CNN = convolutional neural network; PPV = positive predictive value; Random = uninformed random prediction.

**Table A6.**
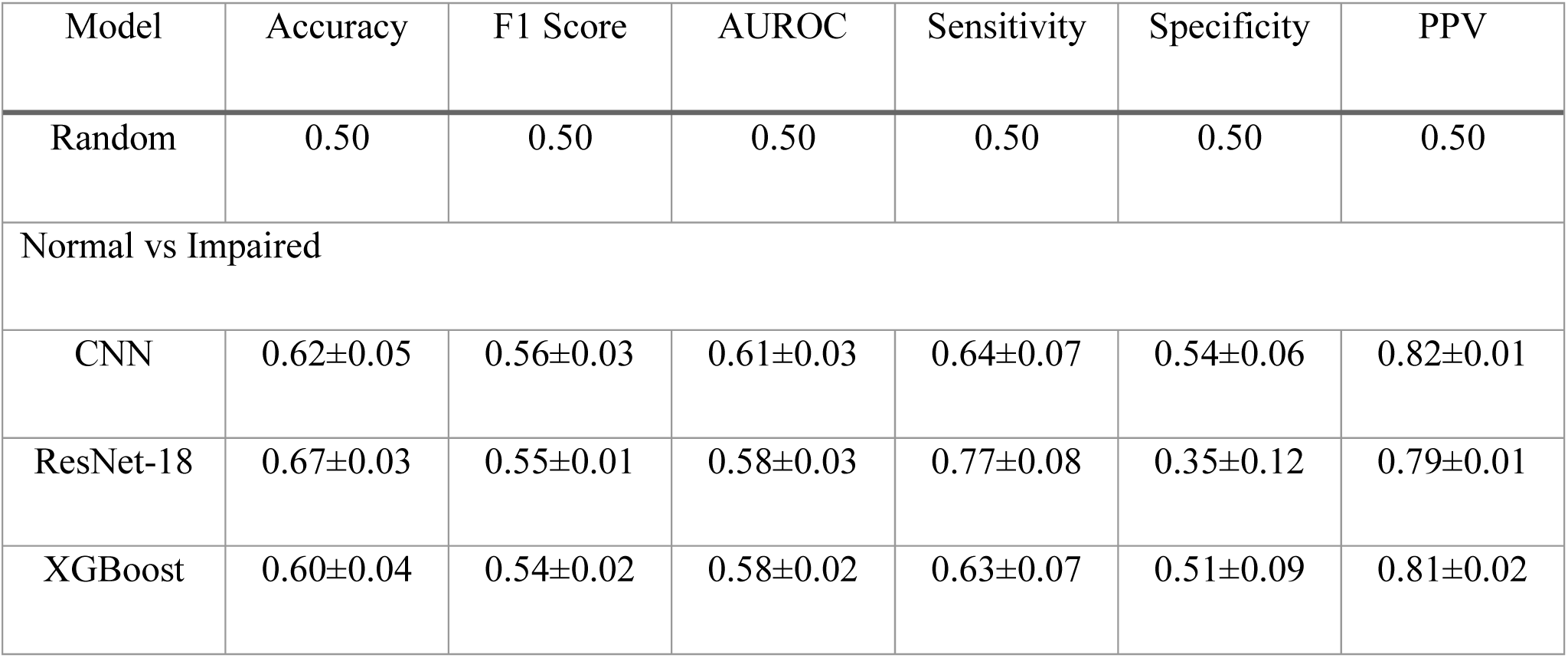
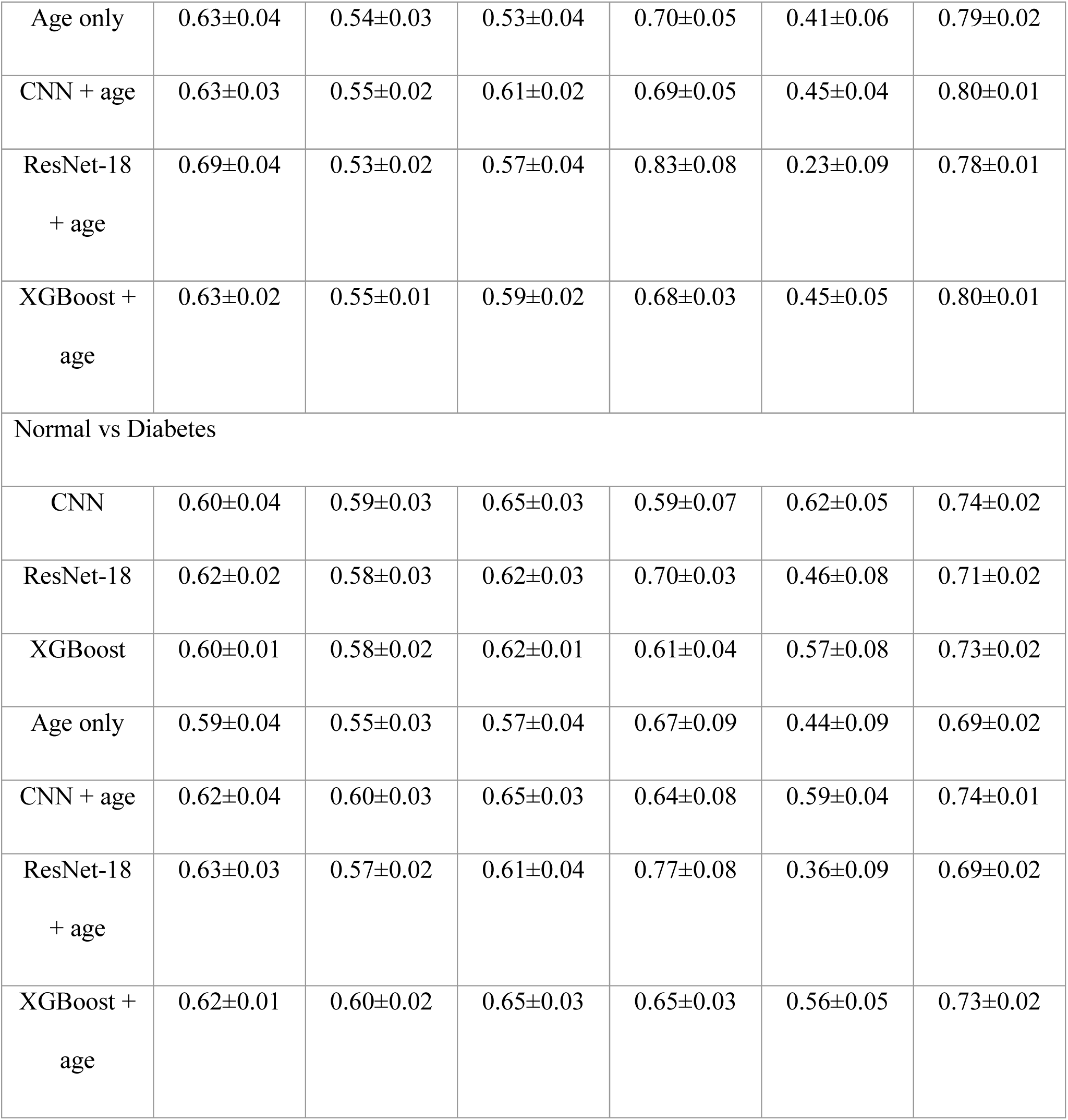
Model performance for binary classification tasks (diagnosis- and treatment-aware labels; uncorrected τCMM maps). For Normal vs. Impaired, prediabetic and diabetic participants were merged into a single Impaired class; for Normal vs. Diabetes, prediabetic participants were excluded. Models and cohorts as in main-text Table 2; XGBoost and Age only were evaluated on the 1,774 participants with complete ETDRS features, CNN and ResNet-18 on all 1,783 participants. Values are mean ± standard deviation across 5 outer test folds of nested cross-validation. AUROC = area under the receiver operating characteristic curve; CNN = convolutional neural network; PPV = positive predictive value; Random = uninformed random prediction.

**Figure A6.**
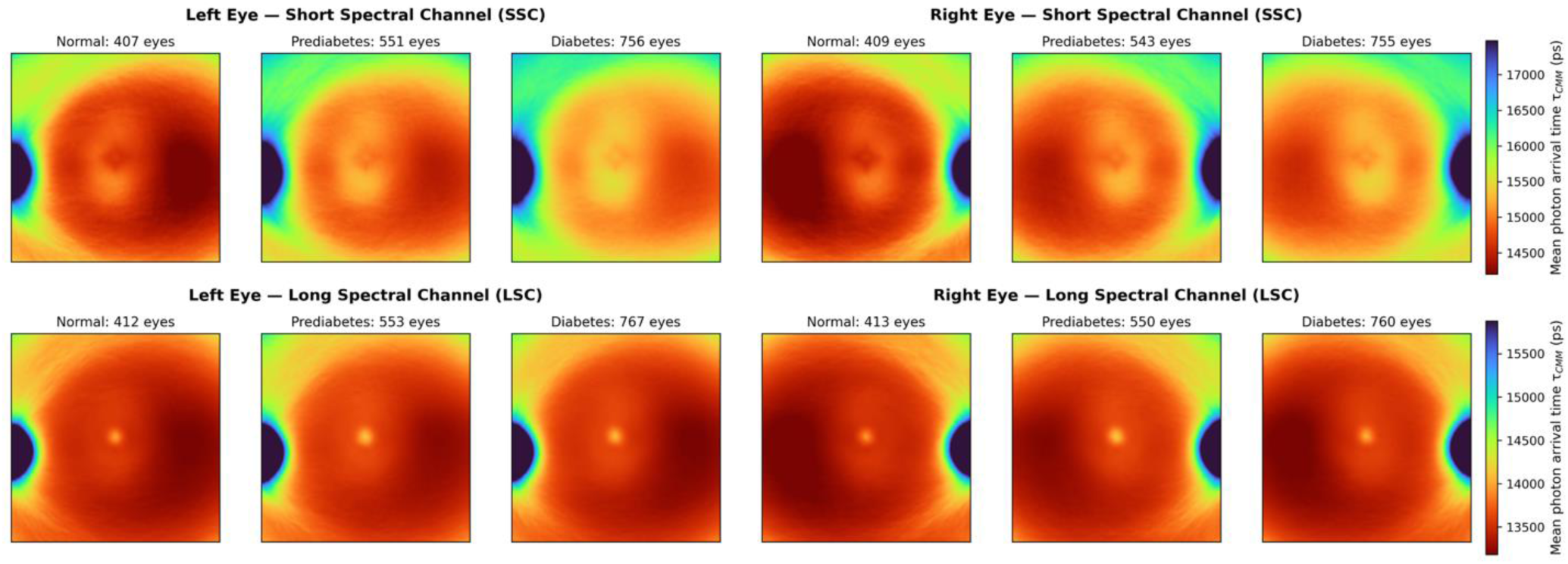
Counterpart of main-text Figure 2 (diagnosis- and treatment-aware labels; uncorrected τCMM maps). Aggregated τCMM maps by glycemic group, spectral channel (SSC, top; LSC, bottom), and eye laterality; sample counts (eyes) above each panel; color scale from red (short) to blue (long), in picoseconds. Uncorrected values include a system timing offset, and the apparently long foveal LSC values reflect background-related intensity coupling of the uncorrected estimator (Section B).

**Figure A7.**
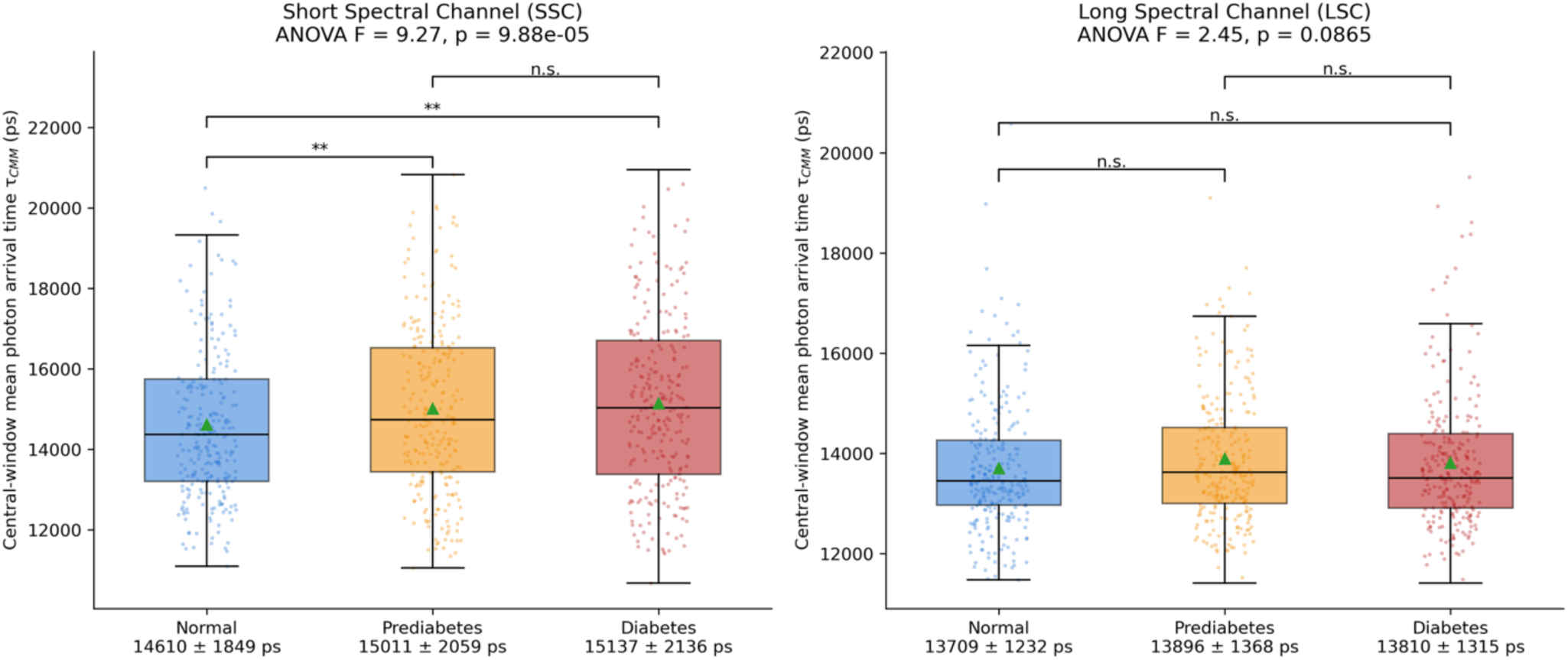
Counterpart of main-text Figure 4 (diagnosis- and treatment-aware labels; uncorrected τCMM maps). Participant-level distributions of τCMM within the central 20 × 20-pixel window by glycemic group and spectral channel; one-way ANOVA F and p in each panel title; brackets denote Bonferroni-corrected pairwise Welch’s t-tests (n.s. = not significant; *p < 0.05; **p < 0.01); group means ± standard deviation (ps) below each x-axis label.

**Figure A8.**
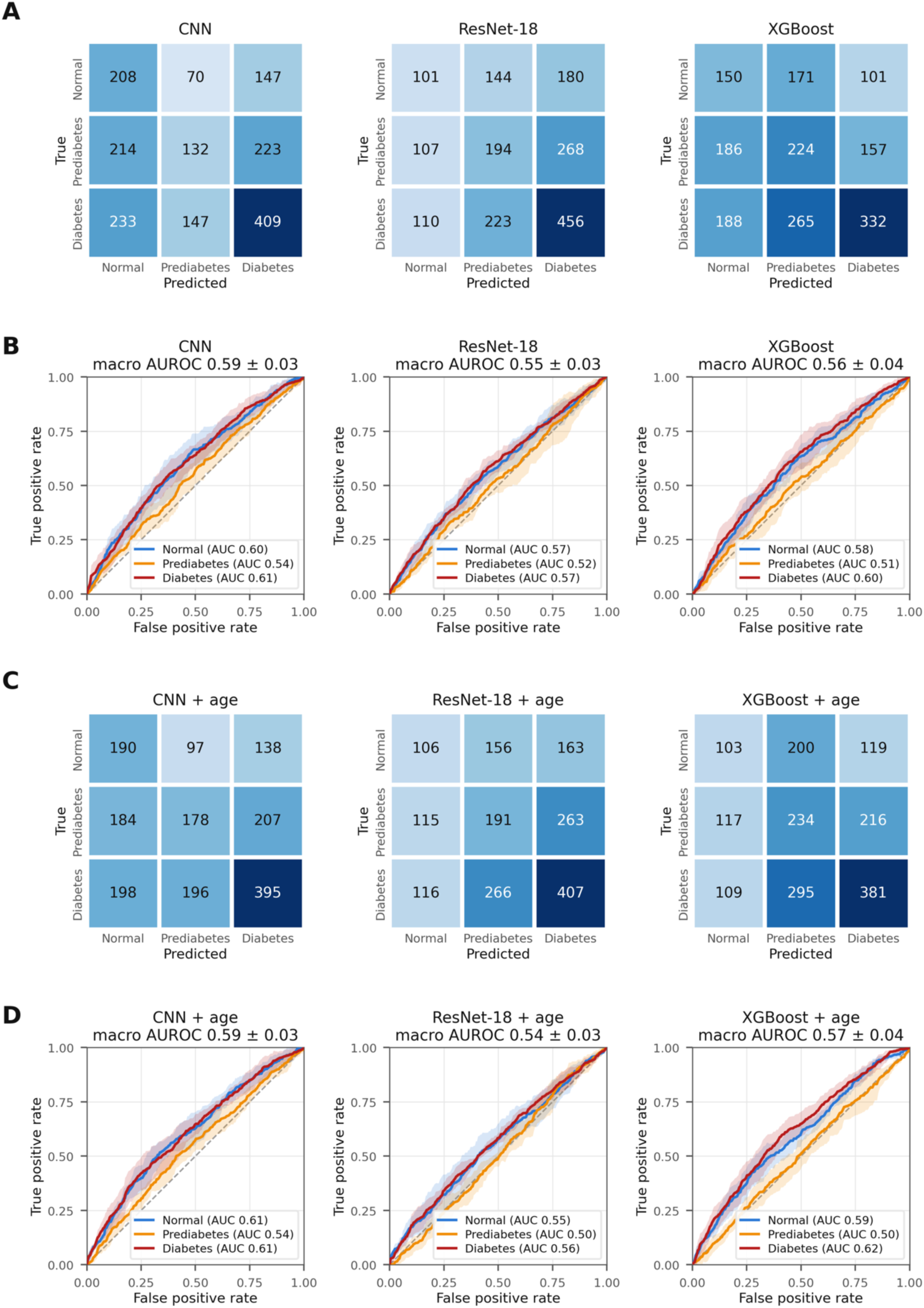
Counterpart of main-text Figure 5 (diagnosis- and treatment-aware labels; uncorrected τCMM maps). (A) Confusion matrices (participant counts, pooled across outer test folds) and (B) per-class ROC curves (mean ± fold-to-fold standard deviation; macro AUROC in panel titles) for the CNN, ResNet-18, and XGBoost FLIO-only models; (C, D) the same for the models with age as an additional input.

**Figure A9.**
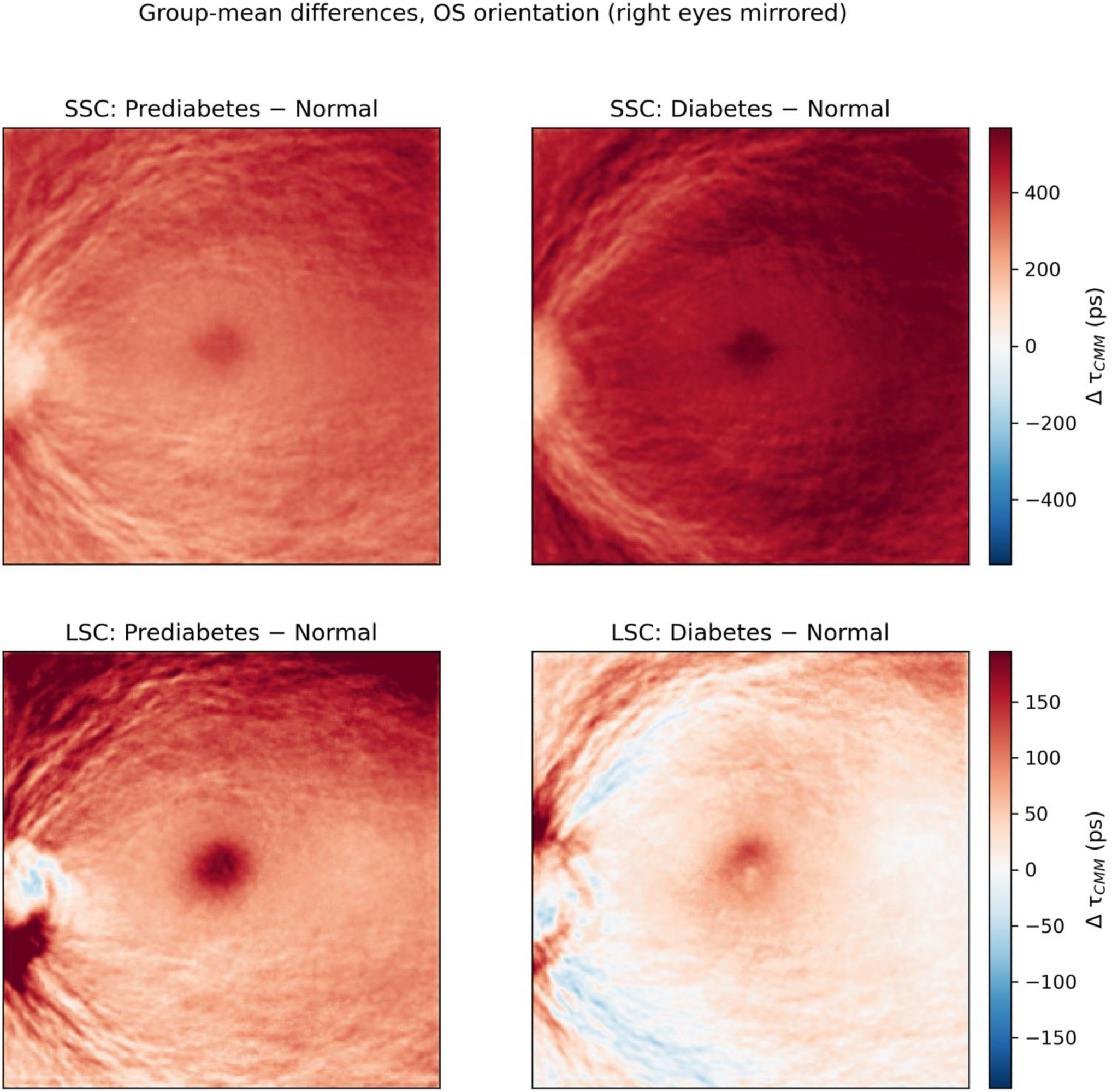
Counterpart of main-text Figure 3 (diagnosis- and treatment-aware labels; uncorrected τCMM maps). Pixel-wise group-mean differences in OS orientation (Prediabetes − Normal, left; Diabetes − Normal, right) for the SSC (top) and LSC (bottom); warm colors indicate longer τCMM in the impaired-glycemia group; color bars in picoseconds. Uncorrected values include a system timing offset, and the apparently long foveal LSC values reflect background-related intensity coupling of the uncorrected estimator (Section B).

#### F.2 HbA1c-only labels, background- and timing-offset-corrected maps

Central-window comparison: SSC ANOVA F = 6.53, p = 0.002 (age-adjusted p = 0.072); LSC ANOVA F = 7.14, p < 0.001 (age-adjusted p = 0.64) (Figures A10–A13).

**Table A7.**
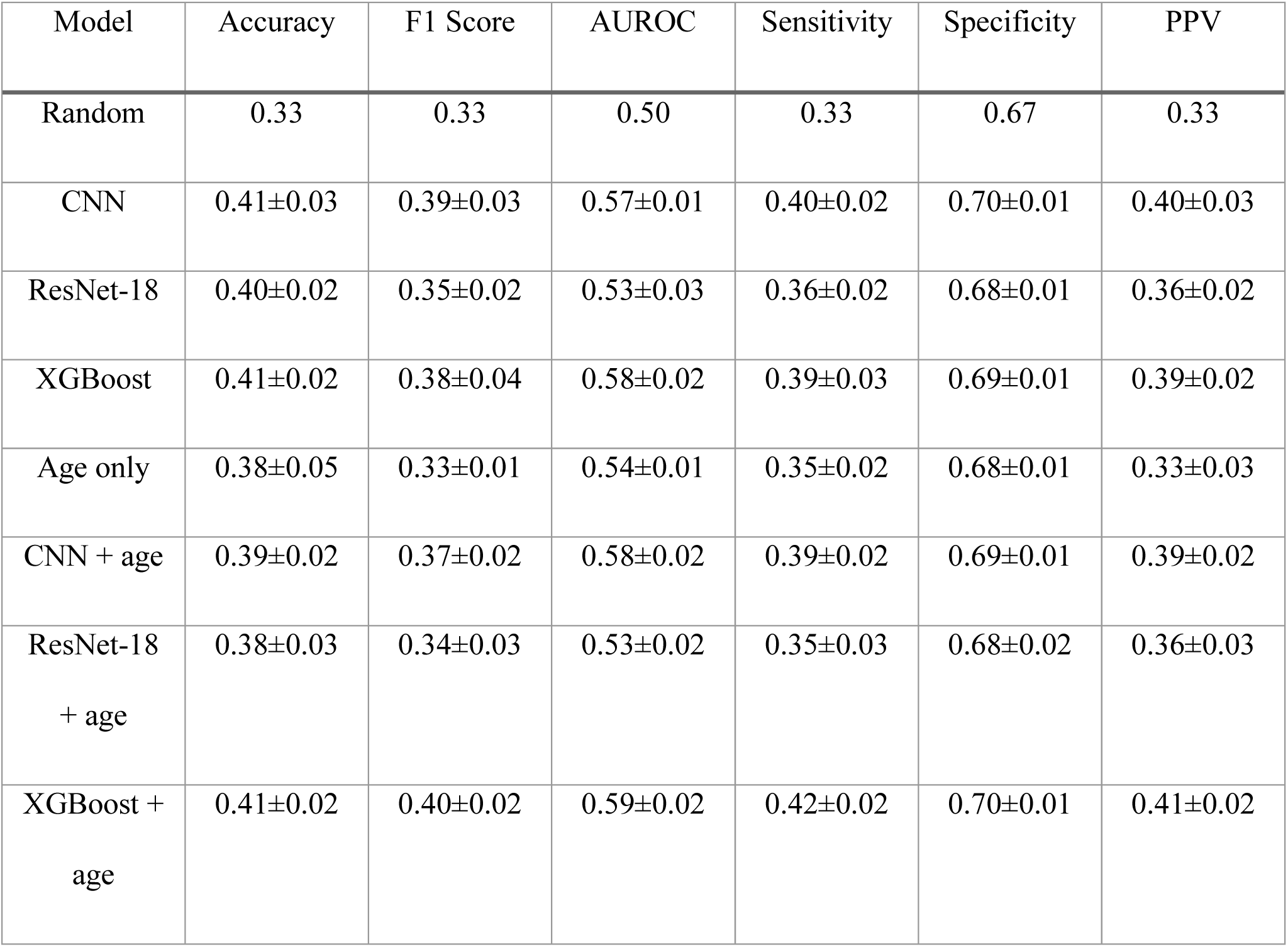
Model performance for 3-class classification, macro-averaged (HbA1c-only labels; background- and timing-offset-corrected τCMM maps). Models and cohorts as in main-text Table 2; XGBoost and Age only were evaluated on the 1,773 participants with complete ETDRS features, CNN and ResNet-18 on all 1,783 participants. Values are mean ± standard deviation across 5 outer test folds of nested cross-validation. AUROC = area under the receiver operating characteristic curve; CNN = convolutional neural network; PPV = positive predictive value; Random = uninformed random prediction.

**Table A8.**
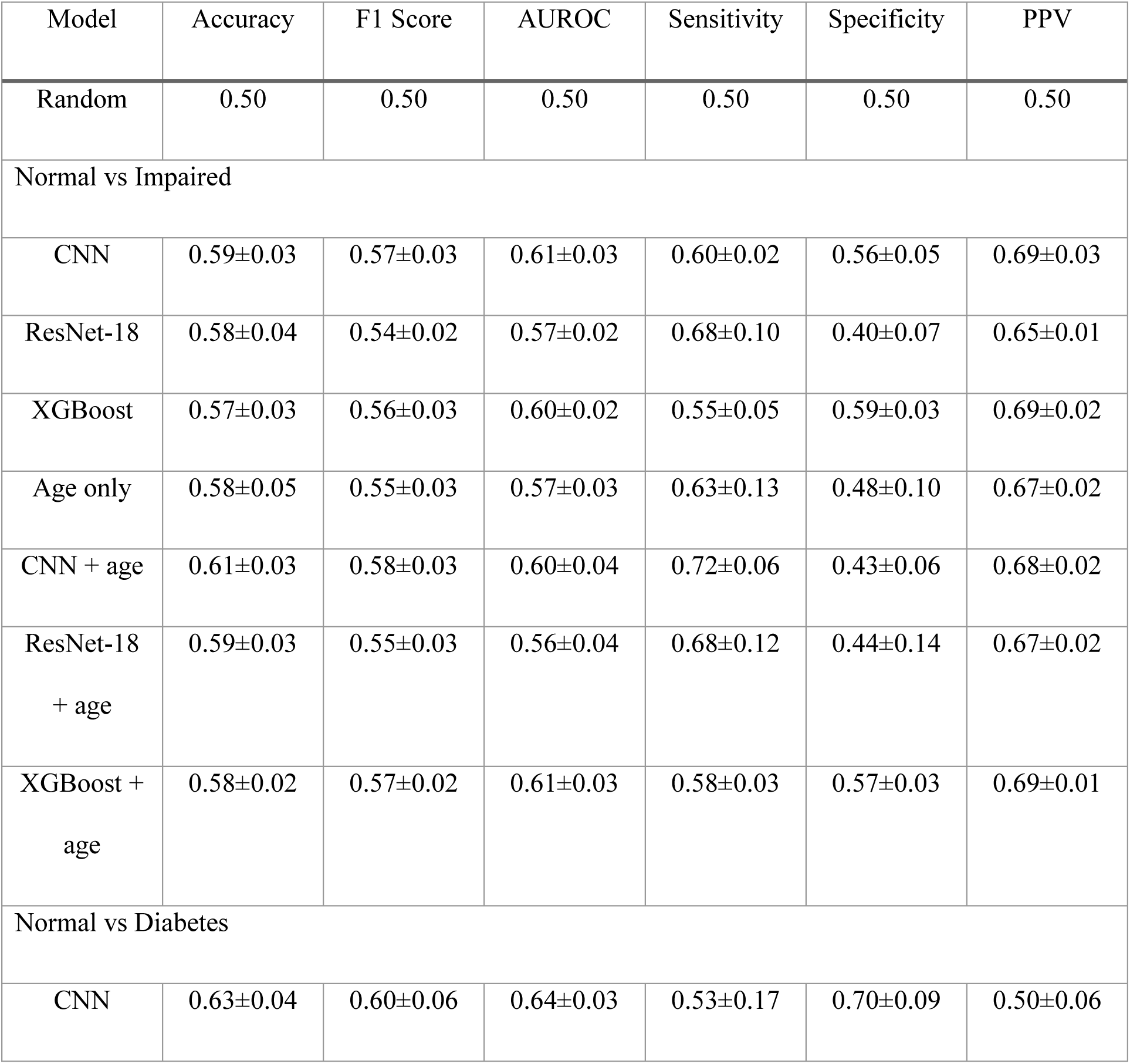
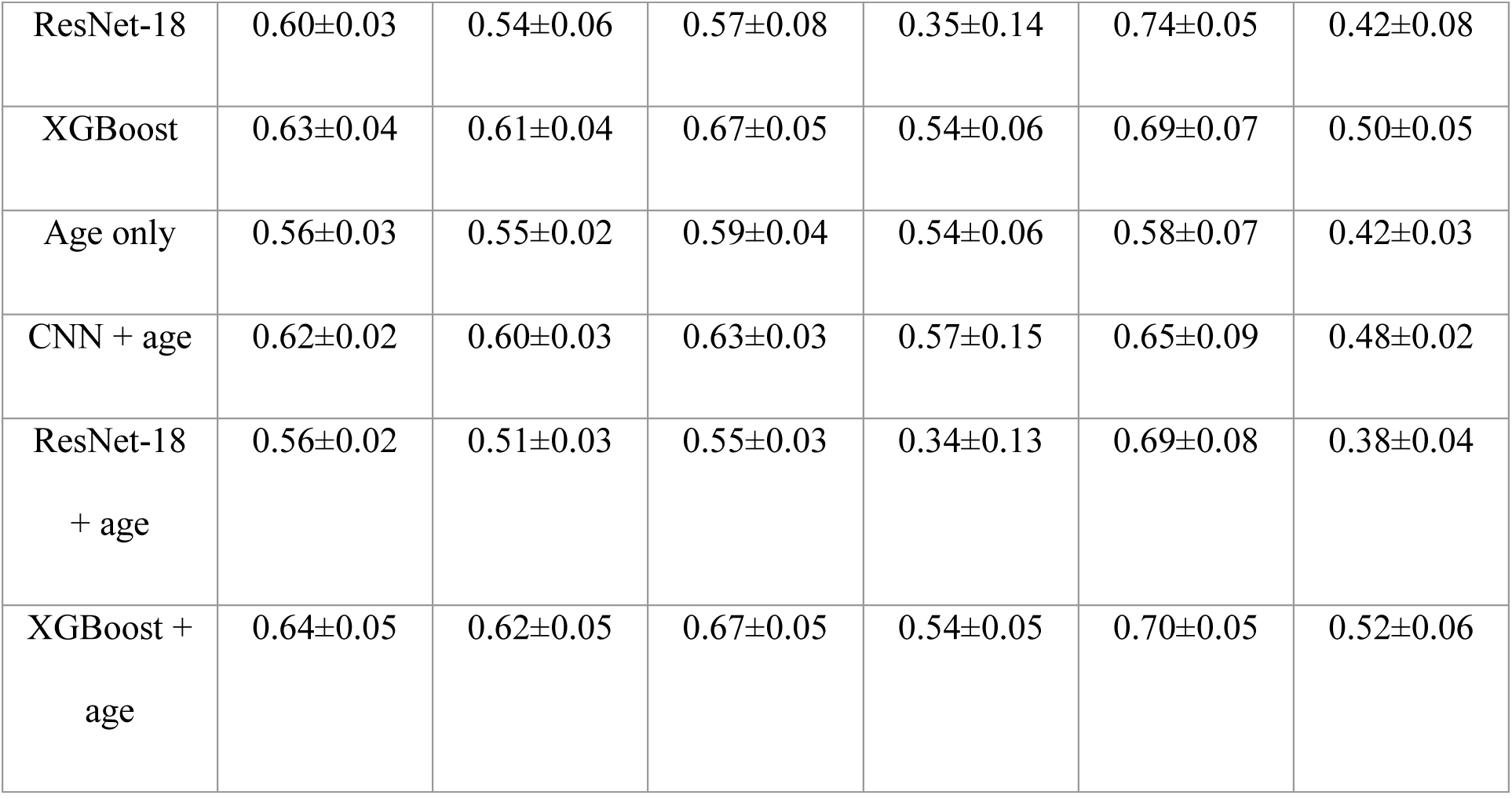
Model performance for binary classification tasks (HbA1c-only labels; background- and timing-offset-corrected τCMM maps). For Normal vs. Impaired, prediabetic and diabetic participants were merged into a single Impaired class; for Normal vs. Diabetes, prediabetic participants were excluded. Models and cohorts as in main-text Table 2; XGBoost and Age only were evaluated on the 1,773 participants with complete ETDRS features, CNN and ResNet-18 on all 1,783 participants. Values are mean ± standard deviation across 5 outer test folds of nested cross-validation. AUROC = area under the receiver operating characteristic curve; CNN = convolutional neural network; PPV = positive predictive value; Random = uninformed random prediction.

**Figure A10.**
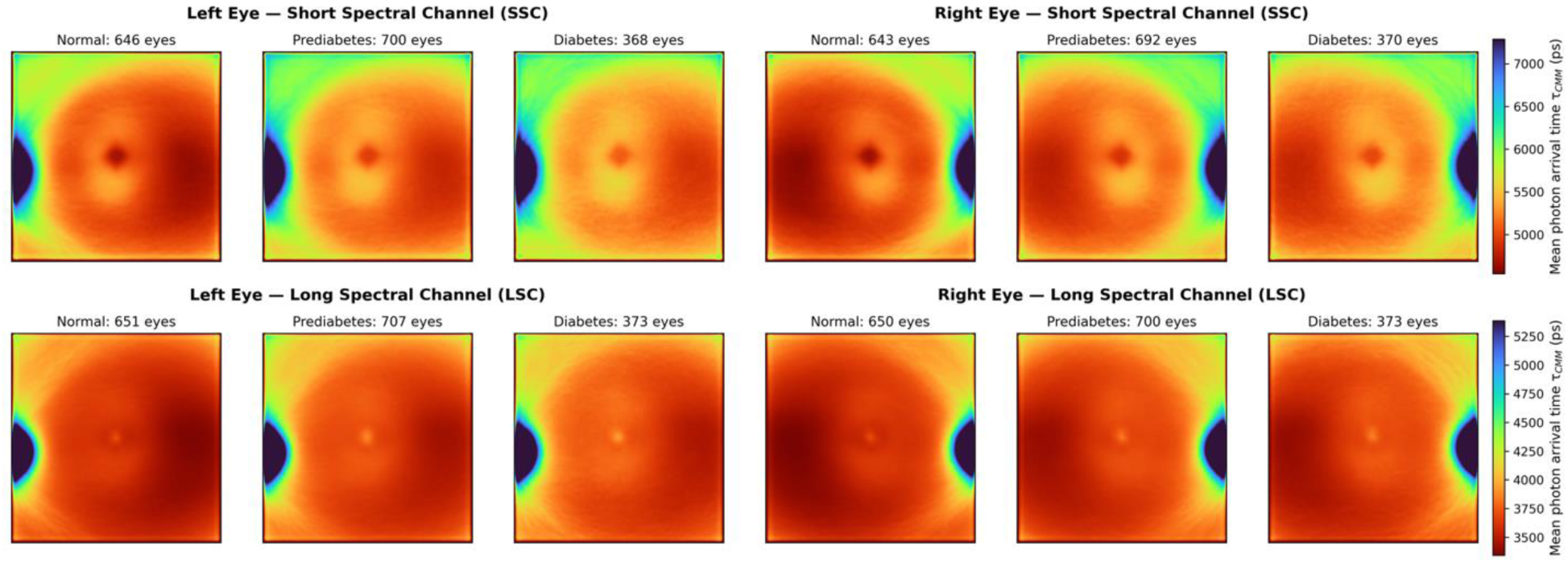
Counterpart of main-text Figure 2 (HbA1c-only labels; background- and timing-offset-corrected τCMM maps). Aggregated τCMM maps by glycemic group, spectral channel (SSC, top; LSC, bottom), and eye laterality; sample counts (eyes) above each panel; color scale from red (short) to blue (long), in picoseconds.

**Figure A11.**
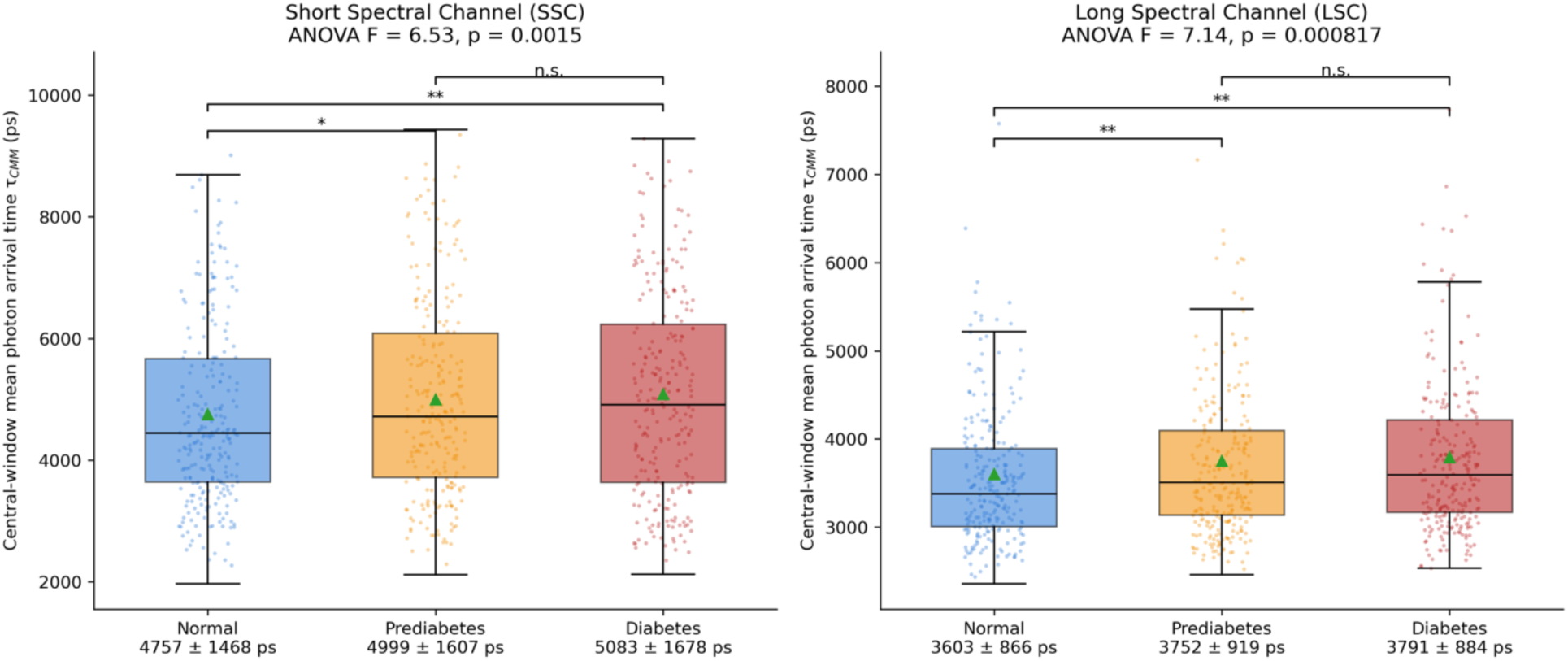
Counterpart of main-text Figure 4 (HbA1c-only labels; background- and timing-offset-corrected τCMM maps). Participant-level distributions of τCMM within the central 20 × 20-pixel window by glycemic group and spectral channel; one-way ANOVA F and p in each panel title; brackets denote Bonferroni-corrected pairwise Welch’s t-tests (n.s. = not significant; *p < 0.05; **p < 0.01); group means ± standard deviation (ps) below each x-axis label.

**Figure A12.**
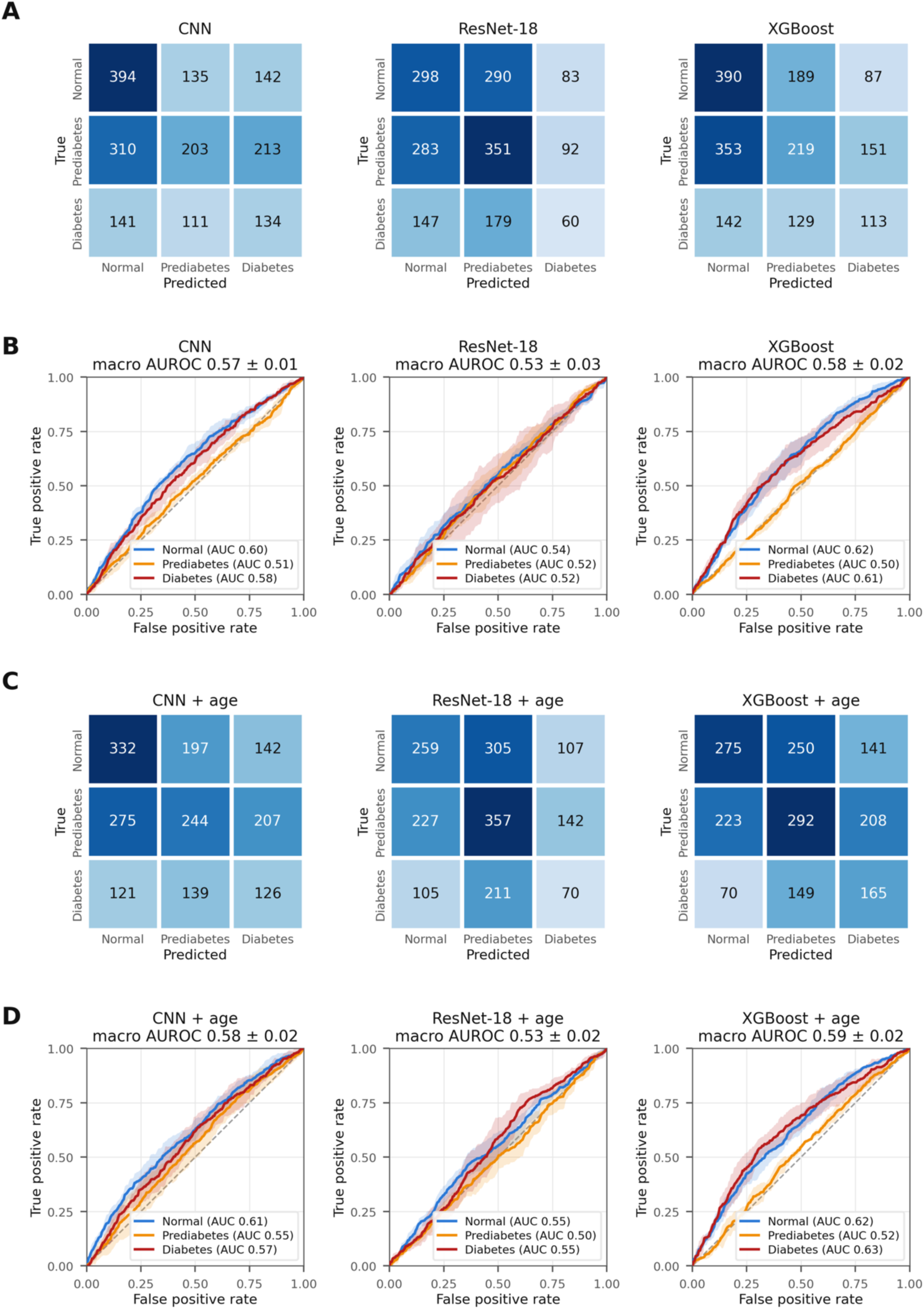
Counterpart of main-text Figure 5 (HbA1c-only labels; background- and timing-offset-corrected τCMM maps). (A) Confusion matrices (participant counts, pooled across outer test folds) and (B) per-class ROC curves (mean ± fold-to-fold standard deviation; macro AUROC in panel titles) for the CNN, ResNet-18, and XGBoost FLIO-only models; (C, D) the same for the models with age as an additional input.

**Figure A13.**
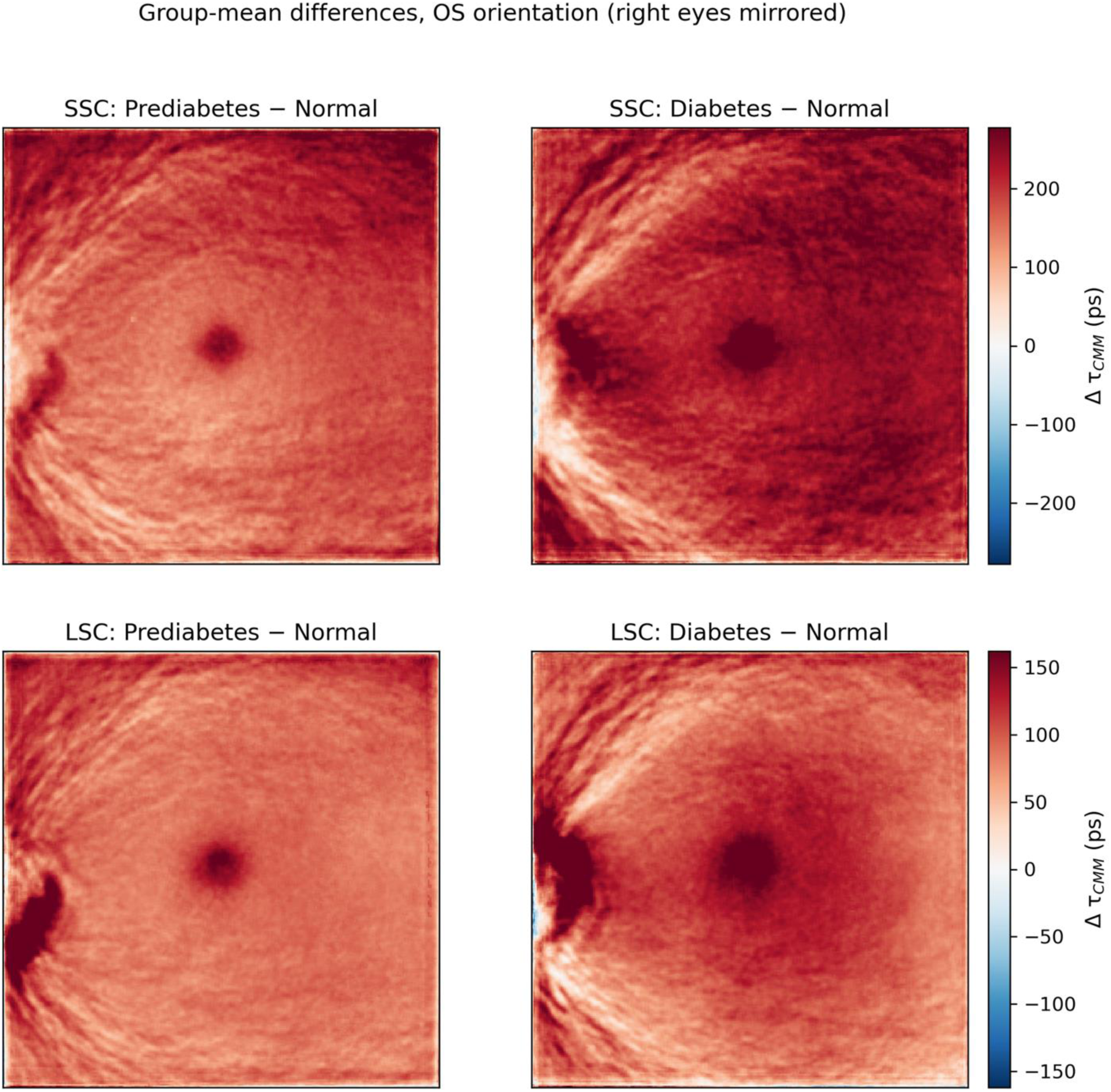
Counterpart of main-text Figure 3 (HbA1c-only labels; background- and timing-offset-corrected τCMM maps). Pixel-wise group-mean differences in OS orientation (Prediabetes − Normal, left; Diabetes − Normal, right) for the SSC (top) and LSC (bottom); warm colors indicate longer τCMM in the impaired-glycemia group; color bars in picoseconds.

#### F.3 HbA1c-only labels, uncorrected maps

Central-window comparison: SSC ANOVA F = 6.85, p = 0.001 (age-adjusted p = 0.049); LSC ANOVA F = 2.89, p = 0.056 (age-adjusted p = 0.73) (Figures A14–A17).

**Table A9.**
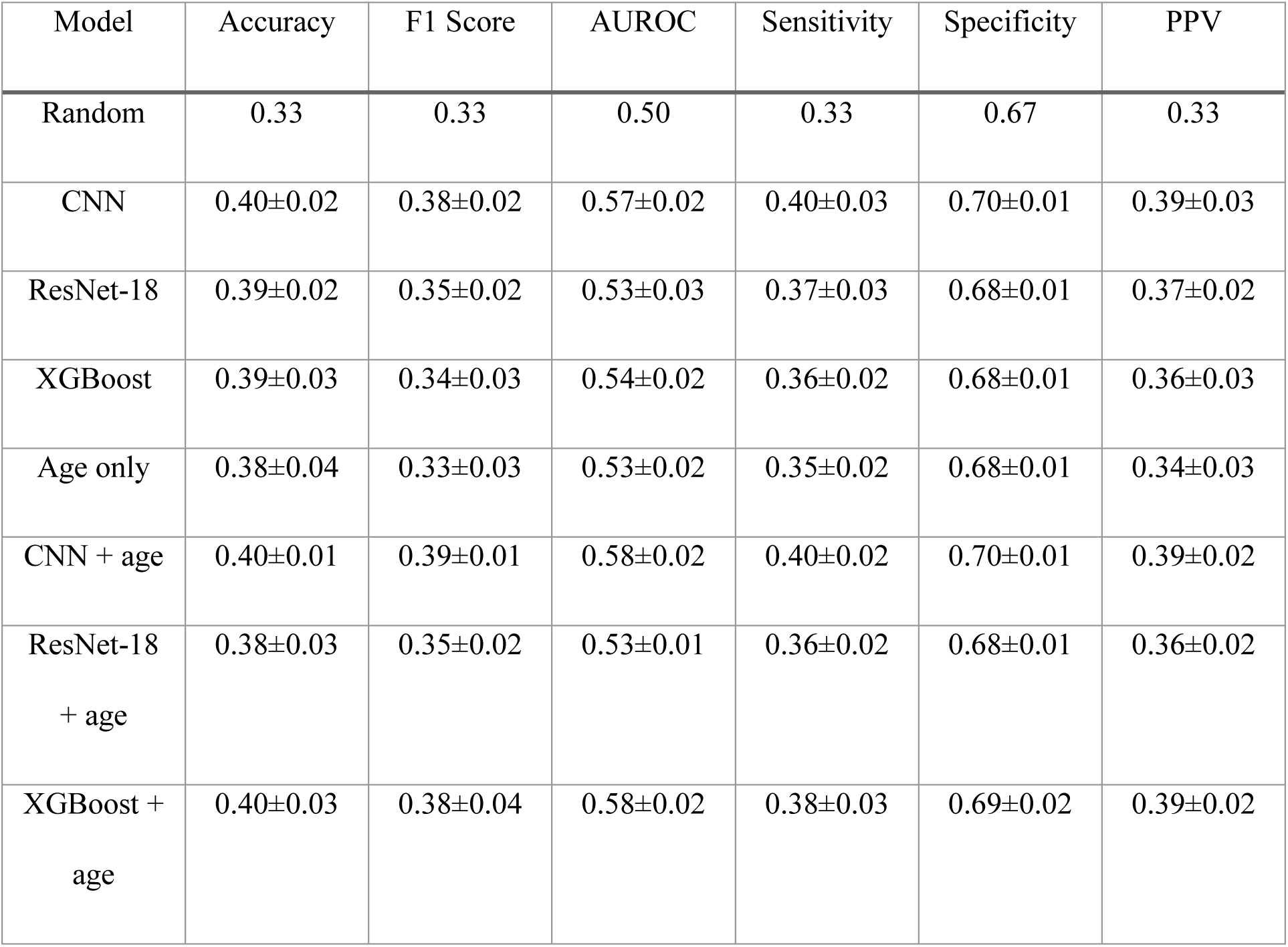
Model performance for 3-class classification, macro-averaged (HbA1c-only labels; uncorrected τCMM maps). Models and cohorts as in main-text Table 2; XGBoost and Age only were evaluated on the 1,774 participants with complete ETDRS features, CNN and ResNet-18 on all 1,783 participants. Values are mean ± standard deviation across 5 outer test folds of nested cross-validation. AUROC = area under the receiver operating characteristic curve; CNN = convolutional neural network; PPV = positive predictive value; Random = uninformed random prediction.

**Table A10.**
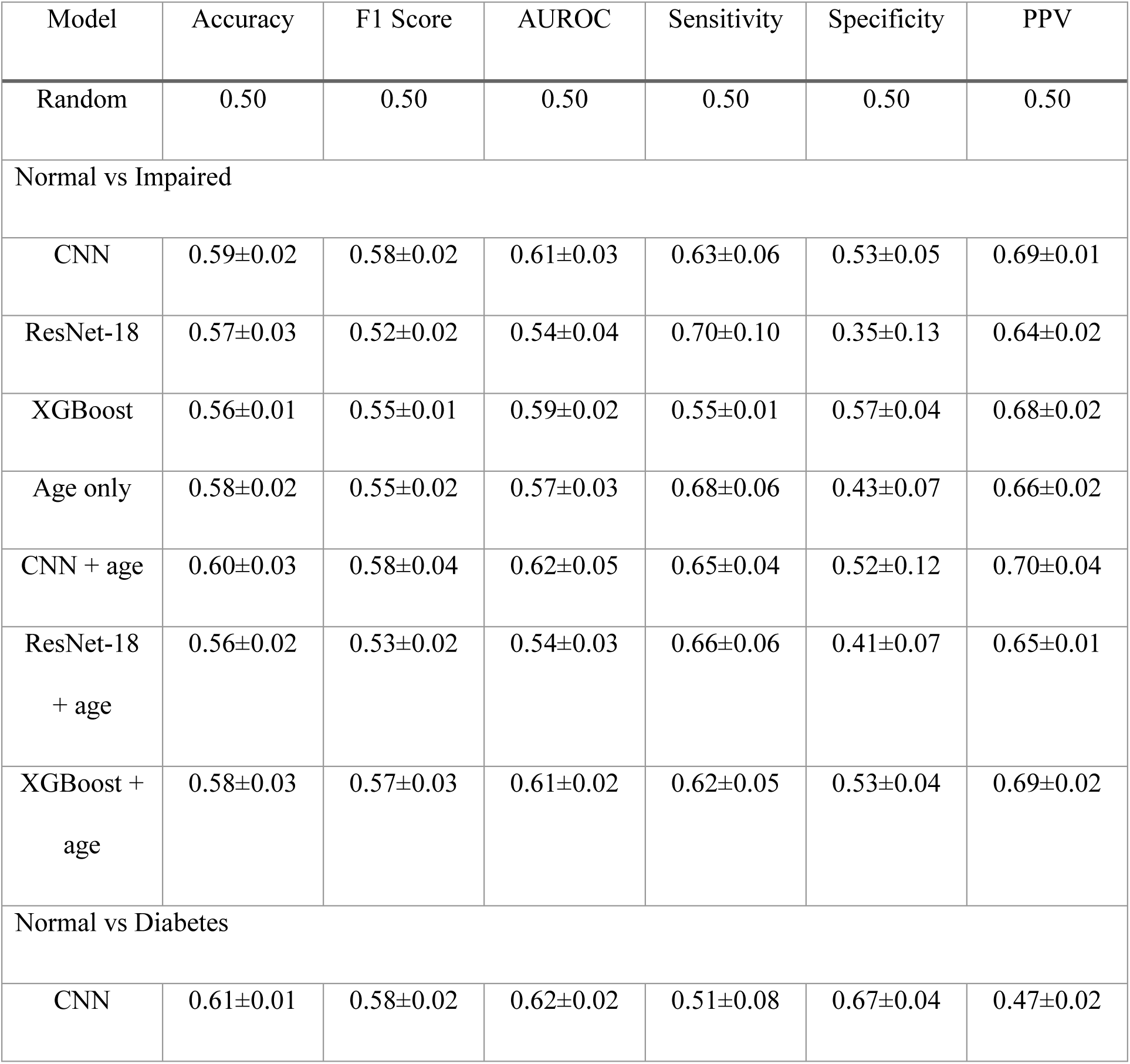
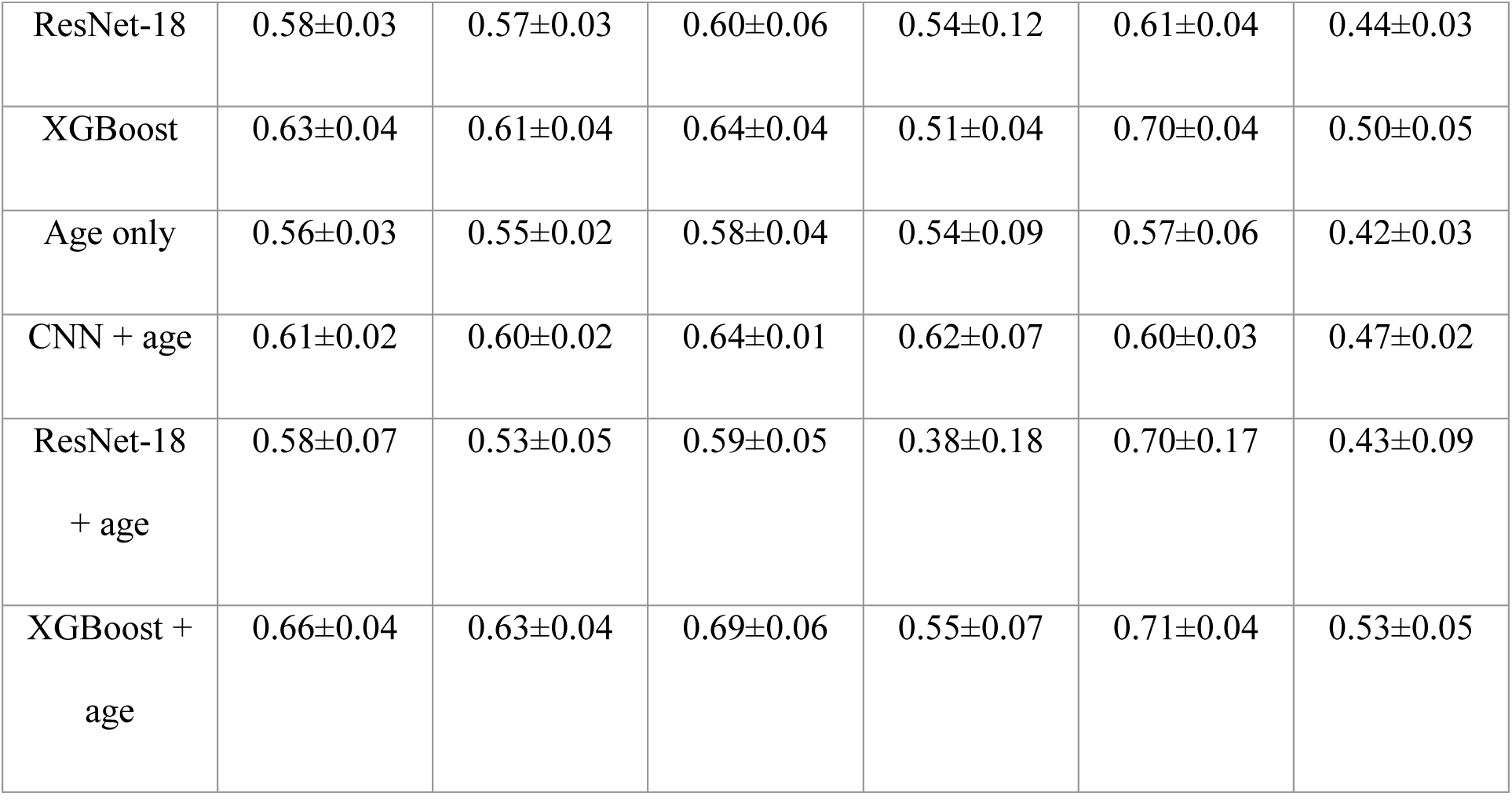
Model performance for binary classification tasks (HbA1c-only labels; uncorrected τCMM maps). For Normal vs. Impaired, prediabetic and diabetic participants were merged into a single Impaired class; for Normal vs. Diabetes, prediabetic participants were excluded. Models and cohorts as in main-text Table 2; XGBoost and Age only were evaluated on the 1,774 participants with complete ETDRS features, CNN and ResNet-18 on all 1,783 participants. Values are mean ± standard deviation across 5 outer test folds of nested cross-validation. AUROC = area under the receiver operating characteristic curve; CNN = convolutional neural network; PPV = positive predictive value; Random = uninformed random prediction.

**Figure A14.**
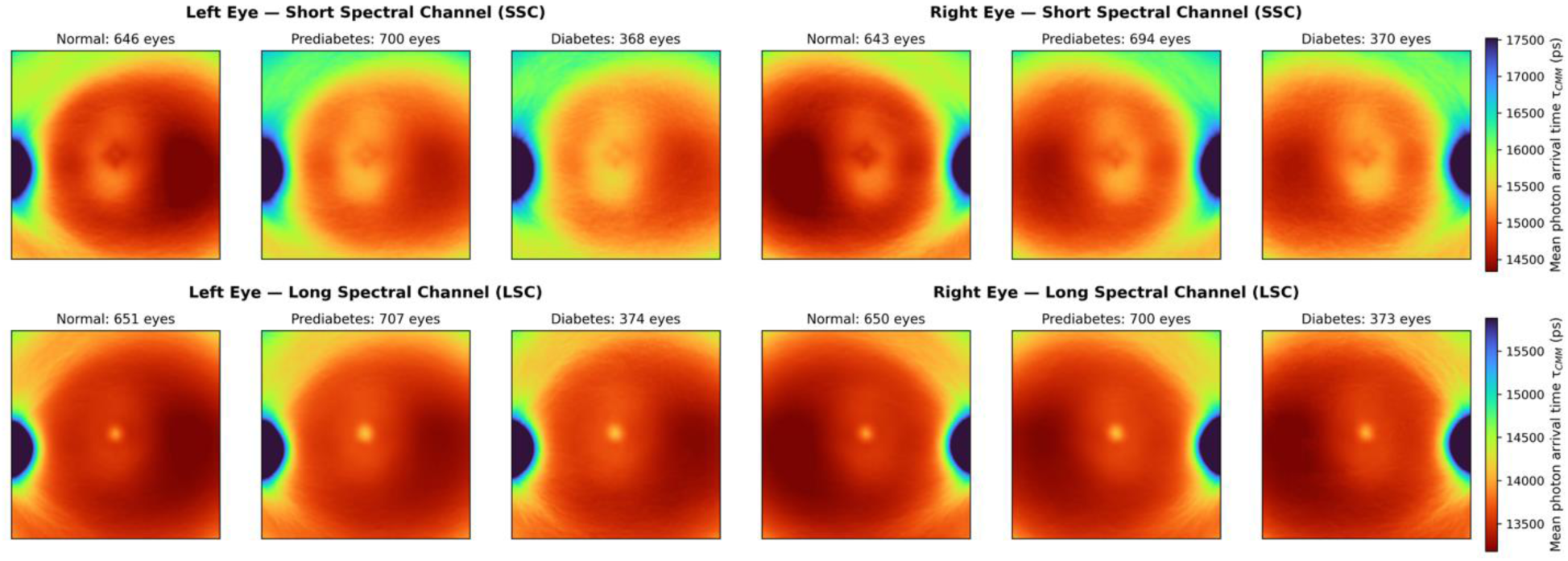
Counterpart of main-text Figure 2 (HbA1c-only labels; uncorrected τCMM maps). Aggregated τCMM maps by glycemic group, spectral channel (SSC, top; LSC, bottom), and eye laterality; sample counts (eyes) above each panel; color scale from red (short) to blue (long), in picoseconds. Uncorrected values include a system timing offset, and the apparently long foveal LSC values reflect background-related intensity coupling of the uncorrected estimator (Section B).

**Figure A15.**
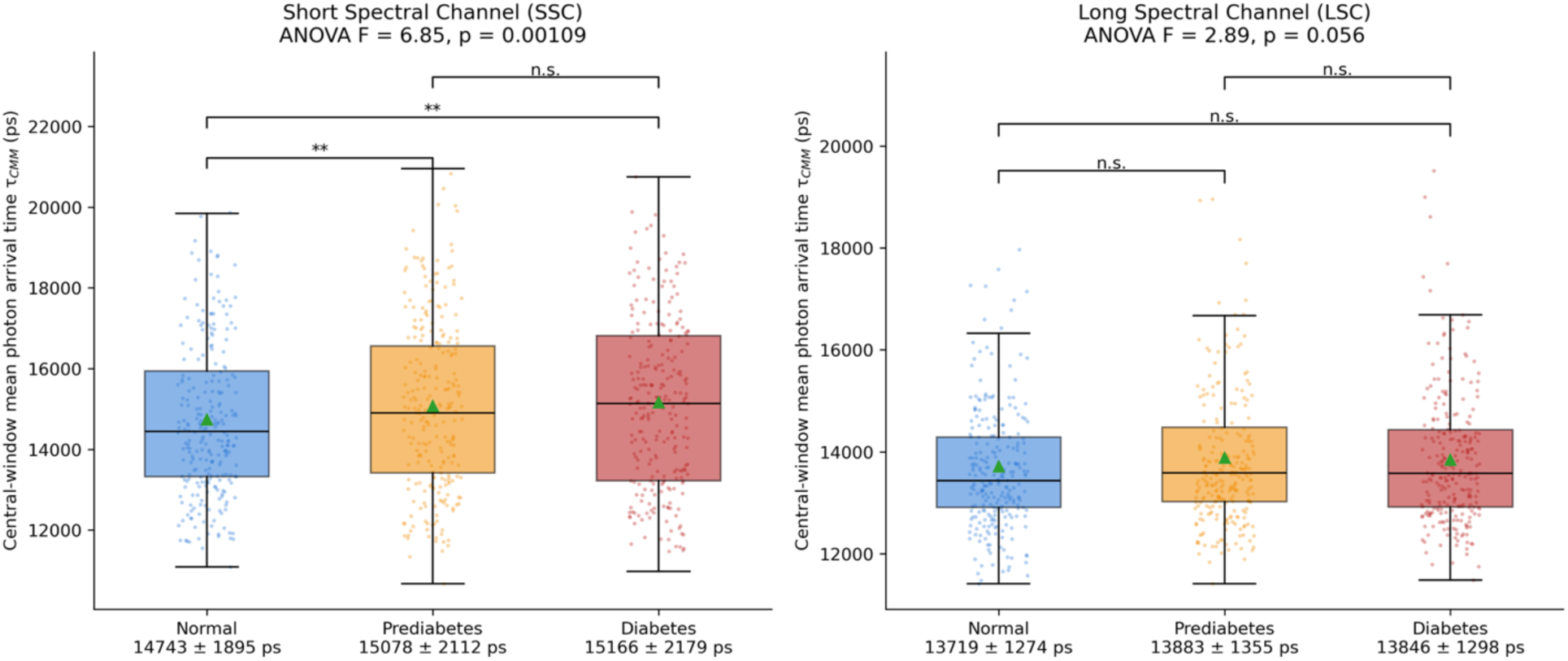
Counterpart of main-text Figure 4 (HbA1c-only labels; uncorrected τCMM maps). Participant-level distributions of τCMM within the central 20 × 20-pixel window by glycemic group and spectral channel; one-way ANOVA F and p in each panel title; brackets denote Bonferroni-corrected pairwise Welch’s t-tests (n.s. = not significant; *p < 0.05; **p < 0.01); group means ± standard deviation (ps) below each x-axis label.

**Figure A16.**
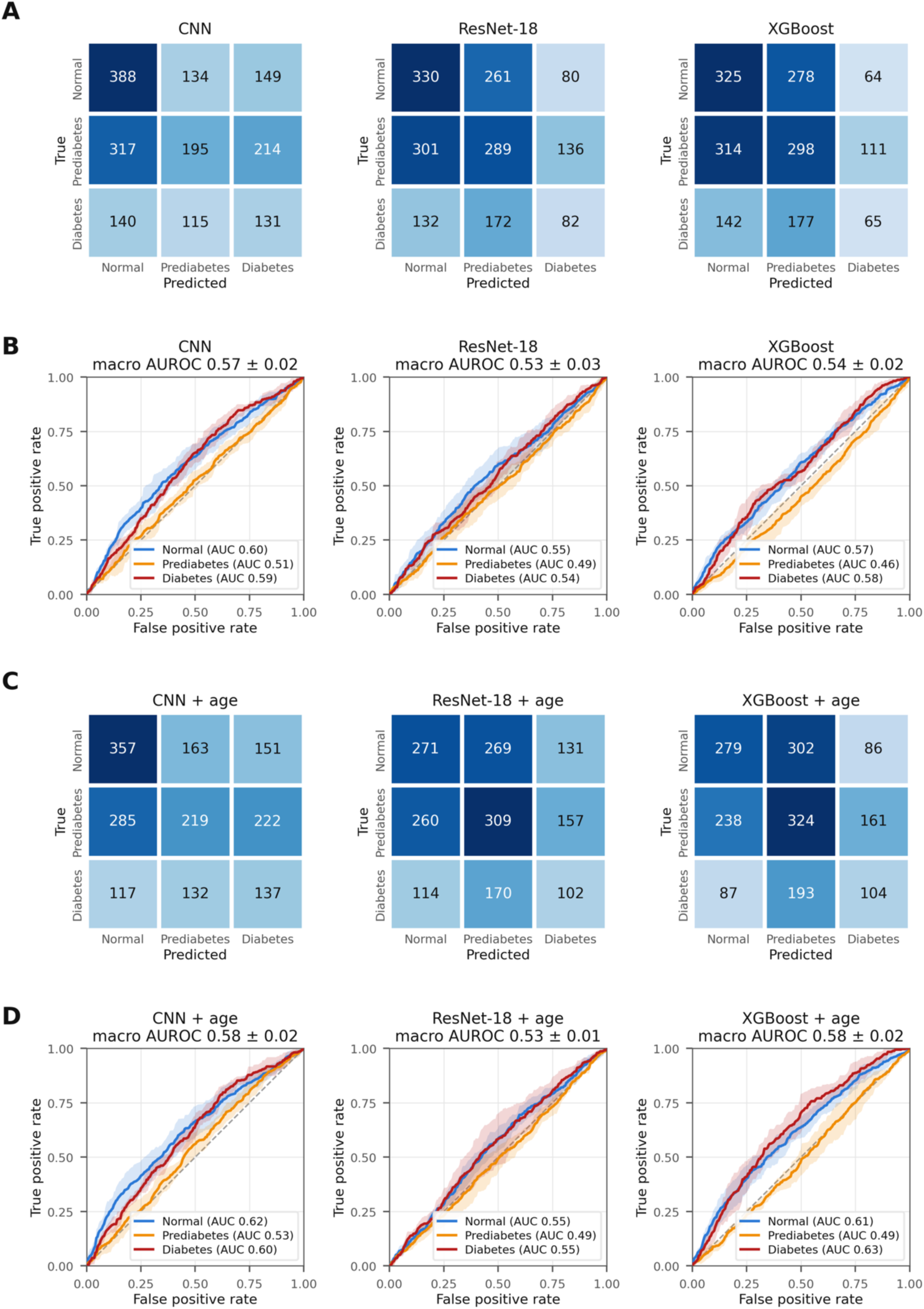
Counterpart of main-text Figure 5 (HbA1c-only labels; uncorrected τCMM maps). (A) Confusion matrices (participant counts, pooled across outer test folds) and (B) per-class ROC curves (mean ± fold-to-fold standard deviation; macro AUROC in panel titles) for the CNN, ResNet-18, and XGBoost FLIO-only models; (C, D) the same for the models with age as an additional input.

**Figure A17.**
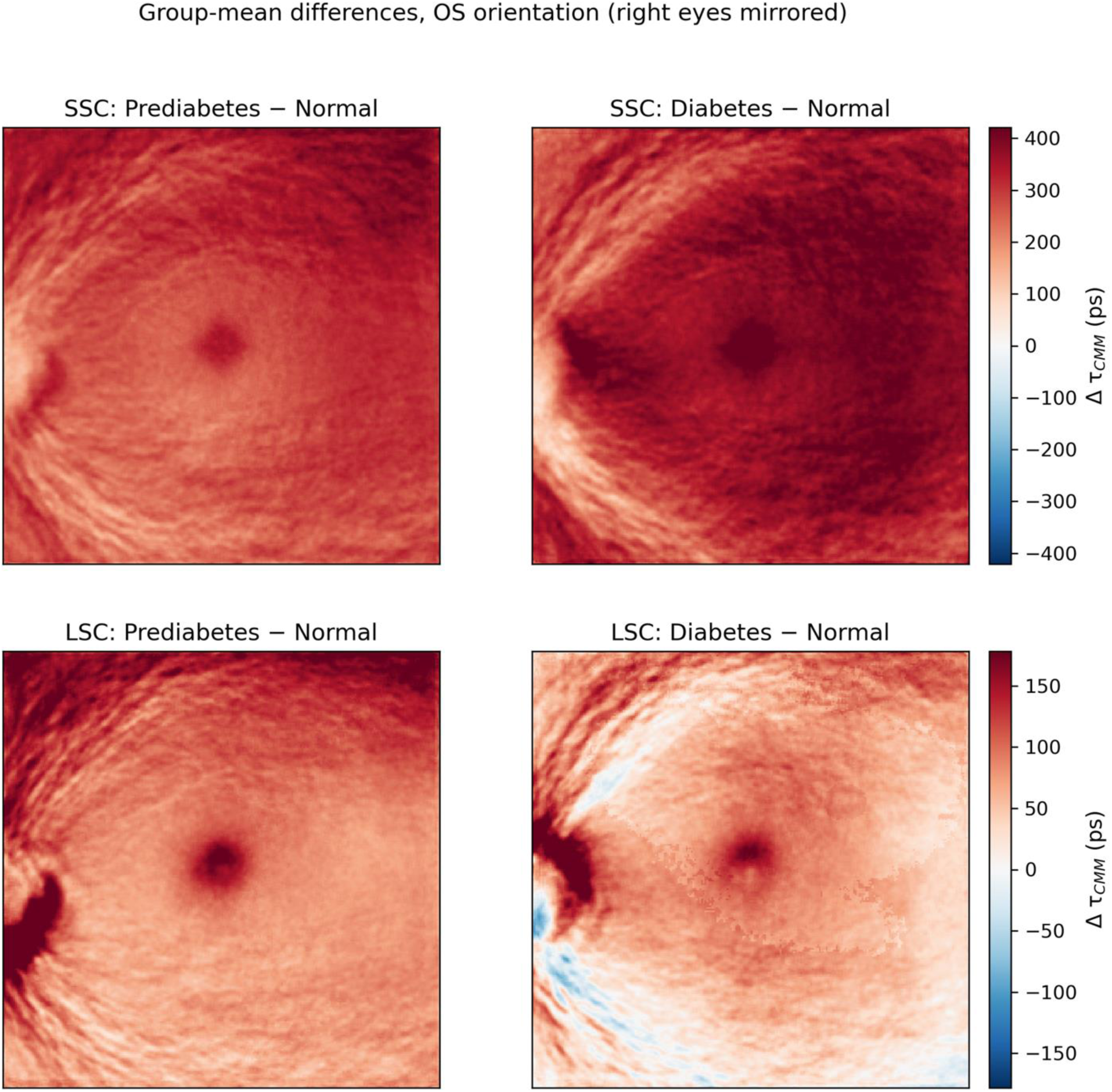
Counterpart of main-text Figure 3 (HbA1c-only labels; uncorrected τCMM maps). Pixel-wise group-mean differences in OS orientation (Prediabetes − Normal, left; Diabetes − Normal, right) for the SSC (top) and LSC (bottom); warm colors indicate longer τCMM in the impaired-glycemia group; color bars in picoseconds. Uncorrected values include a system timing offset, and the apparently long foveal LSC values reflect background-related intensity coupling of the uncorrected estimator (Section B).

