## Supplementary Materials for "Deep Learning of Fluorescence Lifetime Imaging Ophthalmoscopy for Type 2 Diabetes Classification"

**Table of Contents**

A. Model training details

A.1 CNN and ResNet-18

A.2 XGBoost: Table A1, XGBoost hyperparameter grid searched via inner cross-validation

A.3 Class-balanced loss

A.4 Random seeds and reproducibility

A.5 Participant-level aggregation

B. Details about τCMM mean lifetime estimation

B.1 Uncorrected τCMM mean lifetime estimation

B.2 Impact of Intensity Filtering on τCMM Mean-Lifetime Maps

C. Sensitivity analysis of Center Region of Interest

Figure A1. Location and scale of the analysis regions (20 × 20-pixel window and ETDRS rings)

Figure A2. Aggregated SSC τCMM map with the sweep-window outlines

Figure A3. Per-group mean τCMM vs. central-window size (SSC, LSC)

Figure A4. ANOVA and pairwise Welch p-values vs. central-window size (SSC, LSC)

D. HbA1c Prediction

D.1 Model training

D.2 Results: Figure A5, Bland–Altman plots for the CNN, ResNet-18 and XGBoost regression models

E. Sensitivity analyses: comorbidities

Table A2. Central-window τCMM group comparison after ocular and systemic comorbidity exclusions

Table A3. Classification performance (AUROC) after ocular comorbidity exclusions

F. Results under alternative label and map definitions

Table A4. Central-window τCMM group comparison across the four label–map combinations

F.1 Diagnosis- and treatment-aware labels, uncorrected maps

Tables A5 (3-class) and A6 (binary)

Figures A6–A9, the counterparts of main-text Figures 2, 4, 5 and 3

F.2 HbA1c-only labels, background- and timing-offset-corrected maps

Tables A7 (3-class) and A8 (binary)

Figures A10–A13, counterparts of Figures 2, 4, 5 and 3

F.3 HbA1c-only labels, uncorrected maps

Tables A9 (3-class) and A10 (binary)

Figures A14–A17, counterparts of Figures 2, 4, 5 and 3

### A. Model training details

#### A.1 CNN and ResNet-18

The CNN and ResNet-18 models were trained with the Adam optimizer (learning rate $1 \times 10^{-4}$, weight decay $1 \times 10^{-4}$) for up to 50 epochs per fold with a batch size of 16. A ReduceLROnPlateau scheduler (factor 0.5, patience 3 epochs) reduced the learning rate when validation macro F1 plateaued, and the model state from the epoch with the highest validation macro F1 was retained as the final fold checkpoint. No gradient clipping or weight averaging was used.

#### A.2 XGBoost

The XGBoost baseline used the hist tree method throughout, with the inner cross-validation grid summarized in Table A1.

**Table A1.** XGBoost hyperparameter grid searched via inner cross-validation.

| Hyperparameter | Search Values |
|---|---|
| **n_estimators** | 300, 500, 800, 1000 |
| **max_depth** | 3, 4, 6 |
| **learning_rate** | 0.01 – 0.05 |
| **subsample** | 0.7, 0.8 |
| **colsample_bytree** | 0.7, 0.8 |
| **reg_lambda** | 2.0, 3.0 |
| **reg_alpha** | 0.0, 0.1, 0.5 |
| **min_child_weight** | 1, 2 |
| **gamma** | 0.0, 0.1 |

| | |
|---|---|
| **early_stopping_rounds** | 50, 75, 100 |

The XGBoost hist tree method was used throughout. Hyperparameter names follow the XGBoost Python API conventions.

### A.3 Class-balanced loss

The deep models were trained with class-weighted cross-entropy. Weights followed the standard scikit-learn formulation,

$$w_i = \frac{N}{K \cdot n_i}$$

where $N$ is the total number of training participants in the fold, $K$ is the number of classes, and $n_i$ is the count of class $i$ in that fold. Weights were renormalized to unit mean before use. The same formulation was applied to the binary tasks with $K = 2$.

### A.4 Random seeds and reproducibility

A global seed of 42 was used for the outer cross-validation. The inner cross-validation seeds were offset by the outer-fold index (seed = 42 + $k$ for outer fold $k$) so that inner splits did not overlap across folds.

### A.5 Participant-level aggregation

Per-eye encoder outputs (256-dimensional for CNN, 512-dimensional for ResNet-18) were aggregated to per-participant representations by mean pooling over each participant's available eyes (1 or 2). For the participants with a single eye, the singleton embedding was used directly without zero-padding. Participant predictions were obtained from the classifier head applied to the pooled embedding. Right-eye maps were mirrored horizontally to the left-eye orientation before encoding (laterality canonicalization). In the FLIO + age models, age scaled as (age − 60)/10 was concatenated to the pooled embedding before the classifier head.

### B. Details about τCMM mean lifetime estimation

B.1 Uncorrected **τCMM mean lifetime estimation**

The mean lifetime at each pixel was estimated as:

$$T_{mean}(x,y) = \frac{\sum_{t=1}^{T} t \cdot I(x,y,t)}{\sum_{t=1}^{T} I(x,y,t)}.$$

The mean lifetimes were converted from bin units to physical time using the frame time recorded in the FLIO metadata which specifies the temporal spacing between bins. The mean fluorescence lifetime at each pixel was computed as:

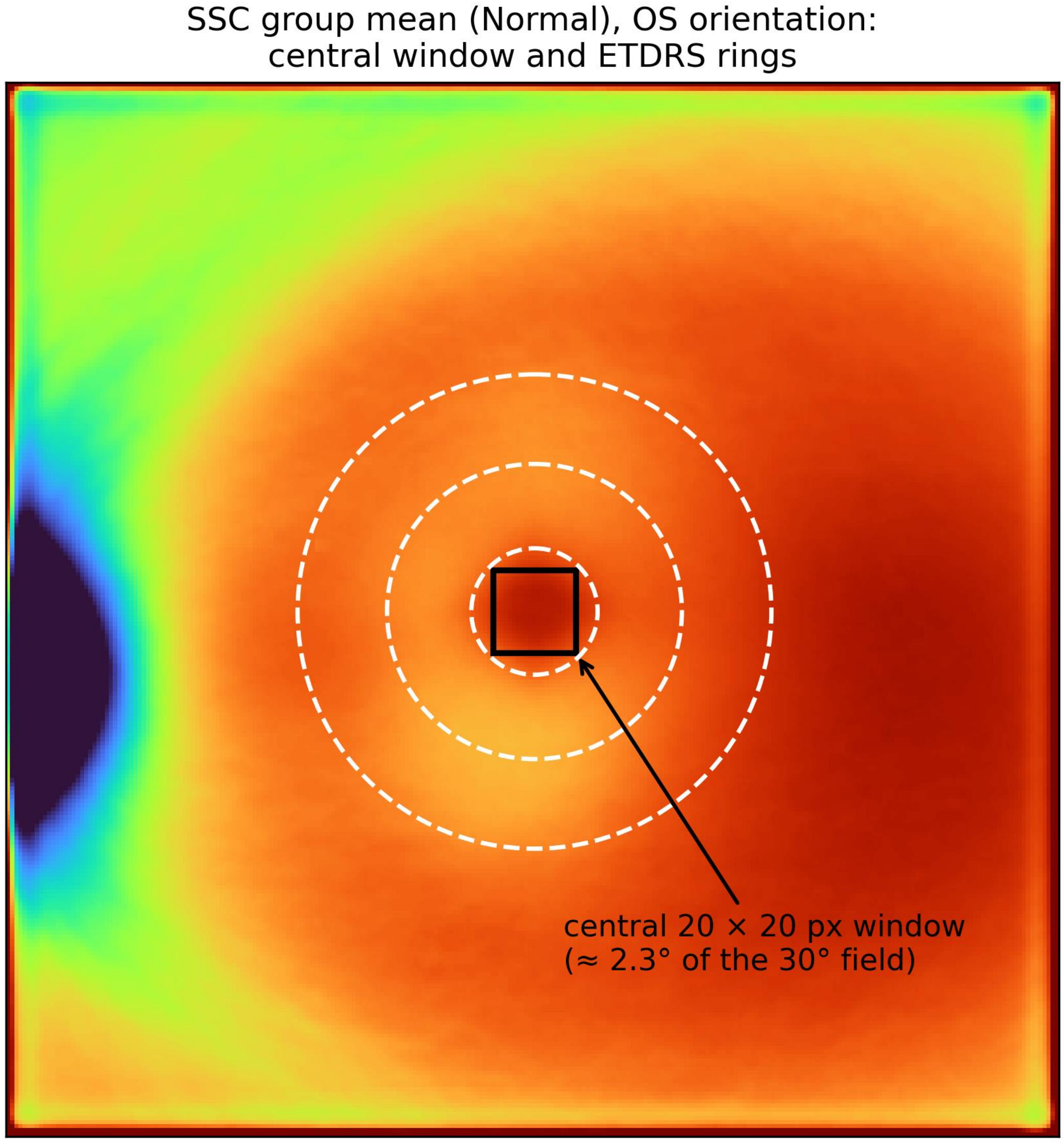


**Figure A1.** Location and scale of the analysis regions. The fixed 20 × 20-pixel central window used in the primary comparison and the ETDRS rings are overlaid on the group-mean SSC τCMM map of normal participants. The window is centered on the image, not on the fovea. ETDRS = Early Treatment Diabetic Retinopathy Study; SSC = short spectral channel; τCMM = center-of-mass mean lifetime.

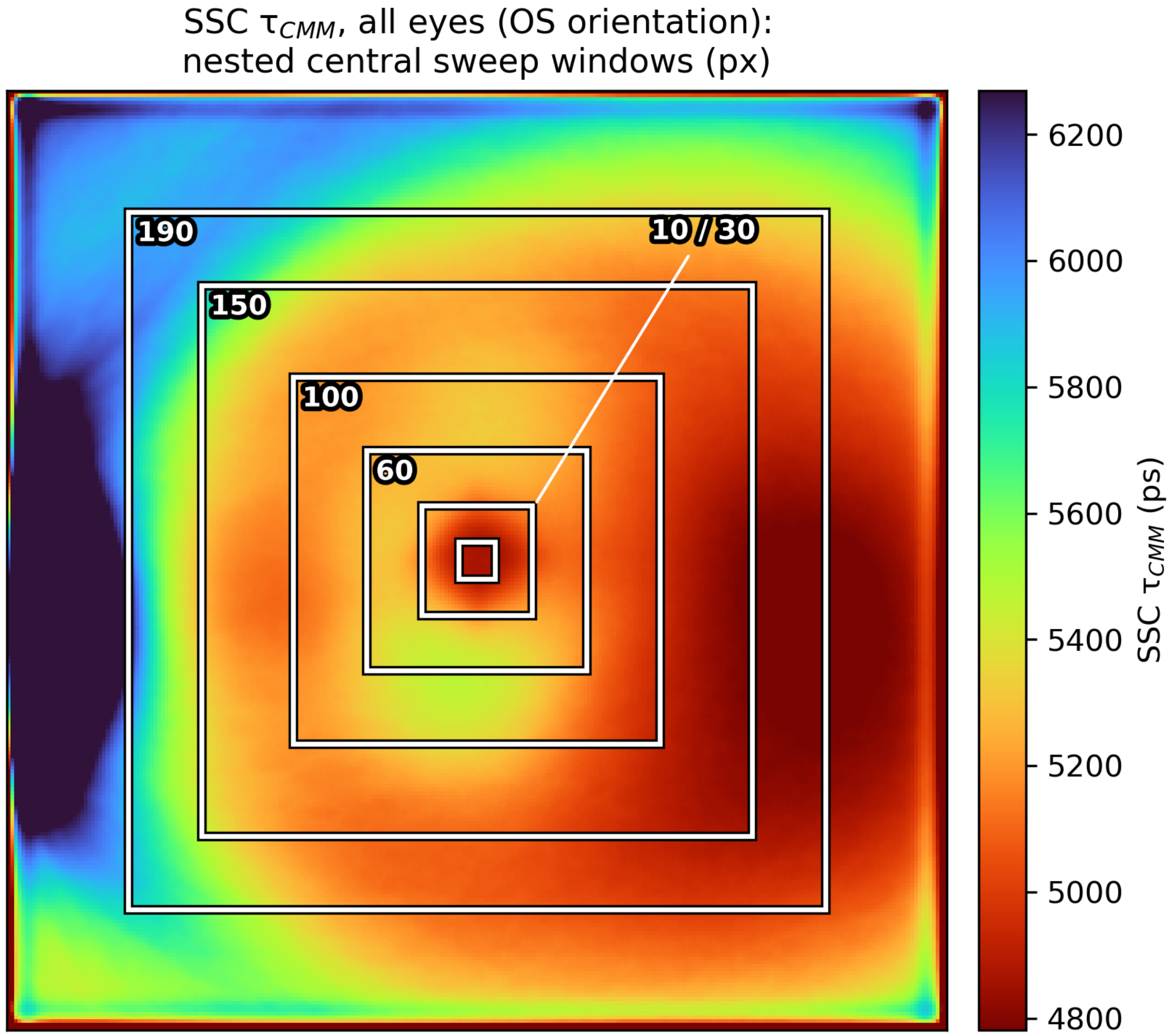


**Figure A2. Aggregated short-spectral-channel (SSC) τCMM map (corrected maps, all eyes, left-eye orientation) with the sweep-window outlines overlaid.** Nested square windows centered on the image indicate the sweep sizes (10 × 10, 30 × 30, 60 × 60, 100 × 100, 150 × 150, 190 × 190 pixels). Color bar shows SSC τCMM (ps).

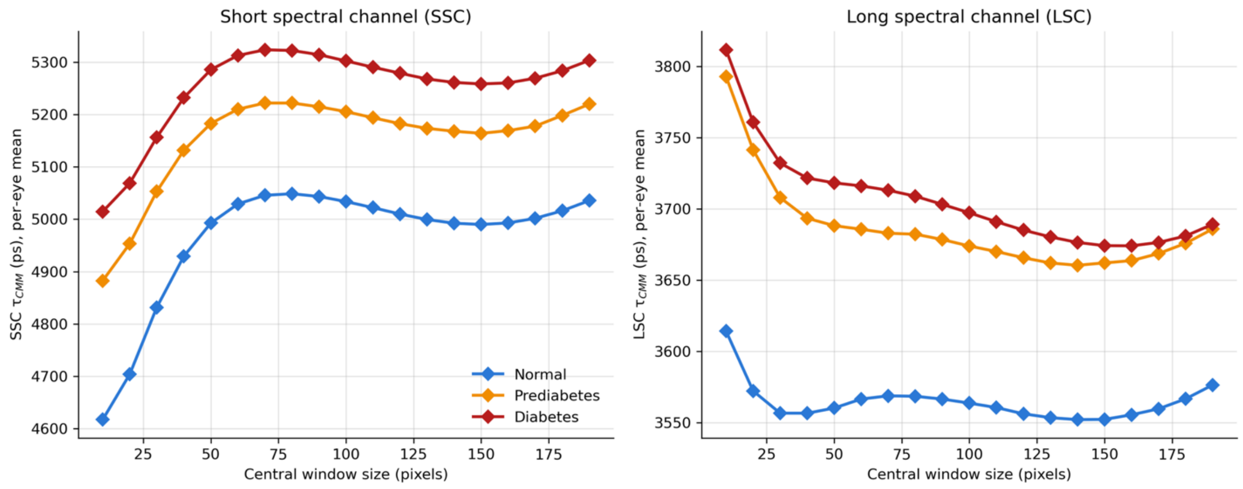


**Figure A3. Per-group mean τCMM as a function of central-window size for the short spectral channel (SSC, left) and long spectral channel (LSC, right), on corrected maps with diagnosis- and treatment-aware groups. Each point is the per-eye mean τCMM averaged across the group at that window size; window sizes were swept from 10 to 190 pixels in 10-pixel steps. Color indicates glycemic group (Normal, blue; Prediabetes, orange; Diabetes, red). τCMM = center-of-mass mean lifetime; ps = picoseconds.**

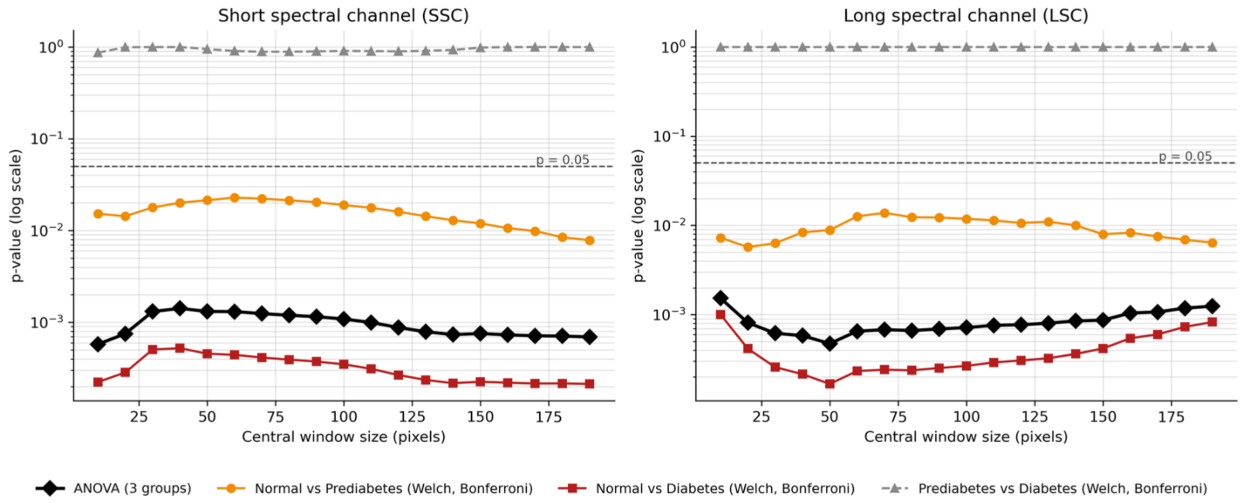


**Figure A4. One-way analysis of variance (ANOVA) and pairwise Welch's t-test p-values (log scale) as a function of central-window size for the short spectral channel (SSC, left) and long spectral channel (LSC, right), on corrected maps with diagnosis- and treatment-aware groups (participant-**

**level means).** Black diamonds: overall ANOVA p across the three groups. Orange circles, red squares, and gray triangles: pairwise Welch's t-tests (normal vs. prediabetes, normal vs. diabetes, prediabetes vs. diabetes), Bonferroni-corrected across three contrasts. The dashed gray horizontal line indicates $p = 0.05$.

### D. HbA1c Prediction

#### D.1 Model training

To assess whether FLIO-derived features can capture continuous glycemic variation, we trained models to predict HbA1c values directly from τCMM maps. The same model architectures described for classification (CNN, ResNet18, and XGBoost) were used, with the primary modification being the replacement of the final classification layer with a regression output layer.

Models were trained using the same procedures as in the classification setting. For deep learning models, Smooth L1 loss was used for optimization, while XGBoost models were trained using regression objectives.

Model performance was assessed using mean absolute error (MAE). Agreement between predicted and observed HbA1c values was evaluated using Bland–Altman analysis.

#### D.2 Results

Regression models trained to predict continuous HbA1c values achieved a test mean absolute error of approximately 0.76. However, predicted values showed strong collapse toward the mean HbA1c level, indicating that the model failed to capture meaningful variation in HbA1c across patients. These regression analyses were performed with the eye-level models of the uncorrected maps.

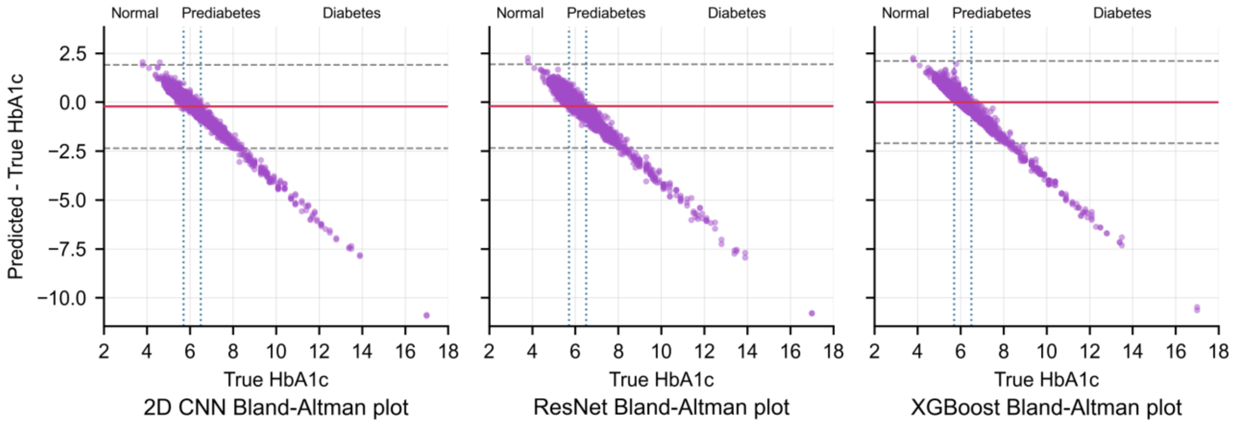


**Figure A5.** Bland-Altman plots comparing predicted versus true hemoglobin A1c (HbA1c) for the 2D CNN, ResNet-18, and XGBoost regression models. The x-axis shows true HbA1c (%); the y-axis shows the prediction error (predicted − true). The solid red line indicates the mean bias; dashed gray lines indicate the 95% limits of agreement (bias ± 1.96 × standard deviation). Vertical dotted lines mark the clinical HbA1c cutoffs (5.7% and 6.4%); Normal / Prediabetes / Diabetes labels above each panel indicate the corresponding glycemic ranges. CNN = convolutional neural network.

**E. Sensitivity analyses: comorbidities**

On the primary labels and corrected maps, the central-window group comparison (Table A2) was repeated after excluding, in turn, participants with each self-reported ocular condition (diabetic retinopathy, cataract, glaucoma, or age-related macular degeneration) and each self-reported systemic condition (hypertension, high cholesterol, obesity, kidney disease, or cardiovascular disease), and after excluding participants with any ocular condition, any systemic condition, or any ocular or systemic condition. All three classifiers were retrained, under the same nested cross-validation protocol, after excluding each of the four ocular conditions and after excluding any of the four ocular conditions (Table A3).

**Table A2.** Central-window τCMM group comparison after excluding participants with self-reported ocular or systemic comorbidities (diagnosis- and treatment-aware labels; background- and timing-offset-corrected τCMM maps). n = participants with a valid SSC central-window value. Each single-condition row excludes participants reporting that condition. Any ocular condition = diabetic retinopathy, cataract, glaucoma, AMD, or retinal vascular occlusion (RVO); any systemic condition = hypertension, high cholesterol, obesity, kidney disease, or cardiovascular disease (heart attack, stroke, other heart issues, or circulation problems); any ocular or systemic condition = any of these ten conditions. All conditions are self-reported (Table 1). ANOVA F (p): one-way analysis of variance across the three glycemic groups; age-adjusted F (p): F-test of the group effect (2 df) in an analysis of covariance with age as covariate. $p < 0.05$ was considered statistically significant and bolded.

AMD = age-related macular degeneration; LSC = long spectral channel; SSC = short spectral channel; τCMM = center-of-mass mean lifetime.

| Excluded | n | SSC ANOVA F (p) | SSC age-adjusted F (p) | LSC ANOVA F (p) | LSC age-adjusted F (p) |
|---|---|---|---|---|---|
| None (full cohort) | 1,771 | **7.22 (< 0.001)** | **4.10 (0.017)** | **7.15 (< 0.001)** | 1.86 (0.16) |
| Diabetic retinopathy | 1,699 | **7.50 (< 0.001)** | **4.32 (0.013)** | **6.65 (0.001)** | 1.75 (0.17) |
| Cataract | 1,246 | **17.4 (< 0.001)** | **13.0 (< 0.001)** | **7.39 (< 0.001)** | **3.15 (0.043)** |
| Glaucoma | 1,636 | **8.45 (< 0.001)** | **5.04 (0.007)** | **7.92 (< 0.001)** | 2.62 (0.073) |
| AMD | 1,681 | **9.51 (< 0.001)** | **5.69 (0.003)** | **9.16 (< 0.001)** | 2.41 (0.090) |
| Any ocular condition | 1,133 | **17.5 (< 0.001)** | **12.3 (< 0.001)** | **8.68 (< 0.001)** | **3.84 (0.022)** |
| Hypertension | 883 | **3.58 (0.028)** | 1.91 (0.15) | **4.01 (0.018)** | 1.52 (0.22) |
| High cholesterol | 884 | **3.06 (0.048)** | 1.67 (0.19) | 2.72 (0.067) | 0.67 (0.51) |
| Obesity | 1,129 | **4.21 (0.015)** | 1.54 (0.21) | **5.65 (0.004)** | 0.81 (0.44) |
| Kidney disease | 1,584 | **5.86 (0.003)** | **3.16 (0.043)** | **6.08 (0.002)** | 1.55 (0.21) |

| | | | | | |
|---|---|---|---|---|---|
| Cardiovascular disease | 1,324 | **5.69 (0.003)** | 2.88 (0.056) | **3.63 (0.027)** | 0.70 (0.50) |
| Any systemic condition | 354 | **0.68 (0.51)** | 0.08 (0.92) | 1.52 (0.22) | 0.60 (0.55) |
| Any ocular or systemic condition | 271 | **0.84 (0.43)** | 0.04 (0.96) | 1.40 (0.25) | 0.31 (0.73) |

**Table A3.** Classification performance (AUROC) after excluding participants with self-reported ocular comorbidities (diagnosis- and treatment-aware labels; background- and timing-offset-corrected τCMM maps). Each model was retrained on the exclusion cohort; values are mean ± standard deviation across the 5 outer test folds of nested cross-validation. n = participants evaluated by the CNN and ResNet-18; XGBoost was evaluated on the participants with complete ETDRS features (up to 10 fewer). Any of the four = diabetic retinopathy, cataract, glaucoma, or AMD; unlike the any-ocular-condition row of Table A2, participants reporting retinal vascular occlusion were not additionally excluded. AMD = age-related macular degeneration; AUROC = area under the receiver operating characteristic curve; CNN = convolutional neural network.

| Excluded | n | CNN | ResNet-18 | XGBoost |
|---|---|---|---|---|
| 3-class | | | | |
| None (full cohort) | 1,783 | 0.60±0.04 | 0.57±0.04 | 0.57±0.03 |
| Diabetic retinopathy | 1,711 | 0.57±0.03 | 0.55±0.01 | 0.56±0.02 |
| Cataract | 1,255 | 0.59±0.04 | 0.56±0.02 | 0.54±0.04 |
| Glaucoma | 1,645 | 0.61±0.02 | 0.55±0.02 | 0.55±0.02 |
| AMD | 1,693 | 0.60±0.02 | 0.55±0.01 | 0.56±0.01 |

| | | | | |
|---|---|---|---|---|
| Any of the four | 1,146 | 0.59±0.03 | 0.57±0.02 | 0.55±0.03 |
| Normal vs. diabetes | | | | |
| None (full cohort) | 1,214 | 0.67±0.04 | 0.64±0.04 | 0.62±0.04 |
| Diabetic retinopathy | 1,143 | 0.63±0.05 | 0.63±0.03 | 0.62±0.02 |
| Cataract | 840 | 0.64±0.04 | 0.65±0.06 | 0.61±0.04 |
| Glaucoma | 1,116 | 0.63±0.05 | 0.60±0.05 | 0.61±0.04 |
| AMD | 1,162 | 0.66±0.03 | 0.64±0.03 | 0.64±0.02 |
| Any of the four | 762 | 0.67±0.04 | 0.63±0.05 | 0.63±0.03 |
| Normal vs. impaired | | | | |
| None (full cohort) | 1,783 | 0.61±0.01 | 0.57±0.01 | 0.58±0.05 |
| Diabetic retinopathy | 1,711 | 0.57±0.04 | 0.58±0.02 | 0.56±0.03 |
| Cataract | 1,255 | 0.61±0.02 | 0.59±0.03 | 0.55±0.04 |
| Glaucoma | 1,645 | 0.61±0.03 | 0.57±0.04 | 0.54±0.06 |
| AMD | 1,693 | 0.59±0.05 | 0.57±0.01 | 0.59±0.01 |
| Any of the four | 1,146 | 0.61±0.05 | 0.60±0.04 | 0.56±0.06 |

**F. Results under alternative label and map definitions**

This section reports the complete results for the three alternative combinations of outcome label (diagnosis- and treatment-aware vs. HbA1c-only) and τCMM map (corrected vs. uncorrected); the primary combination (diagnosis- and treatment-aware labels, corrected maps) is reported in the main text. HbA1c-only labels assign normal (HbA1c < 5.7%), prediabetic (5.7–6.4%), and diabetic (> 6.4%) status from the measured HbA1c alone (671, 726, and 386 participants). Uncorrected maps are the center of mass of the raw photon-count histogram without background subtraction or timing-offset removal, as described in Supplementary Section B. All other procedures are identical to the primary analysis.

**Table A4. Central-window τCMM group comparison across the four label–map combinations.**

| **Labels** | **Maps** | **SSC ANOVA F (p)** | **SSC age-adjusted F (p)** | **LSC ANOVA F (p)** | **LSC age-adjusted F (p)** |
|---|---|---|---|---|---|
| Diagnosis- and treatment-aware | Corrected | 7.22 (< 0.001) | 4.10 (0.017) | 7.15 (< 0.001) | 1.86 (0.16) |
| Diagnosis- and treatment-aware | Uncorrected | 9.27 (< 0.001) | 5.88 (0.003) | 2.45 (0.086) | 1.70 (0.18) |
| HbA1c-only | Corrected | 6.53 (0.002) | 2.64 (0.072) | 7.14 (< 0.001) | 0.45 (0.64) |
| HbA1c-only | Uncorrected | 6.85 (0.001) | 3.02 (0.049) | 2.89 (0.056) | 0.31 (0.73) |

**F.1 Diagnosis- and treatment-aware labels, uncorrected maps**

Central-window comparison: SSC ANOVA F = 9.27, p < 0.001 (age-adjusted p = 0.003); LSC ANOVA F = 2.45, p = 0.086 (age-adjusted p = 0.18) (Figures A6–A9).

**Table A5.** Model performance for 3-class classification, macro-averaged (diagnosis- and treatment-aware labels; uncorrected τCMM maps). Models and cohorts as in main-text Table 2; XGBoost and Age only were evaluated on the 1,774 participants with complete ETDRS features, CNN and ResNet-18 on all 1,783 participants. Values are mean ± standard deviation across 5 outer test folds of nested cross-validation. AUROC = area under the receiver operating characteristic curve; CNN = convolutional neural network; PPV = positive predictive value; Random = uninformed random prediction.

| Model | Accuracy | F1 Score | AUROC | Sensitivity | Specificity | PPV |
|---|---|---|---|---|---|---|
| Random | 0.33 | 0.33 | 0.50 | 0.33 | 0.67 | 0.33 |
| CNN | 0.42±0.03 | 0.39±0.04 | 0.59±0.03 | 0.41±0.03 | 0.71±0.01 | 0.41±0.02 |

| | | | | | | |
|---|---|---|---|---|---|---|
| ResNet-18 | 0.42±0.03 | 0.38±0.04 | 0.55±0.03 | 0.39±0.04 | 0.70±0.02 | 0.39±0.04 |
| XGBoost | 0.40±0.04 | 0.38±0.03 | 0.56±0.04 | 0.39±0.03 | 0.70±0.02 | 0.40±0.04 |
| Age only | 0.38±0.05 | 0.36±0.04 | 0.52±0.04 | 0.36±0.04 | 0.68±0.02 | 0.37±0.05 |
| CNN + age | 0.43±0.04 | 0.41±0.05 | 0.59±0.03 | 0.42±0.04 | 0.71±0.02 | 0.42±0.04 |
| ResNet-18 + age | 0.39±0.02 | 0.36±0.03 | 0.54±0.03 | 0.37±0.03 | 0.68±0.01 | 0.37±0.03 |
| XGBoost + age | 0.40±0.04 | 0.37±0.03 | 0.57±0.04 | 0.38±0.03 | 0.69±0.02 | 0.40±0.03 |

**Table A6.** Model performance for binary classification tasks (diagnosis- and treatment-aware labels; uncorrected τCMM maps). For Normal vs. Impaired, prediabetic and diabetic participants were merged into a single Impaired class; for Normal vs. Diabetes, prediabetic participants were excluded. Models and cohorts as in main-text Table 2; XGBoost and Age only were evaluated on the 1,774 participants with complete ETDRS features, CNN and ResNet-18 on all 1,783 participants. Values are mean ± standard deviation across 5 outer test folds of nested cross-validation. AUROC = area under the receiver operating characteristic curve; CNN = convolutional neural network; PPV = positive predictive value; Random = uninformed random prediction.

| Model | Accuracy | F1 Score | AUROC | Sensitivity | Specificity | PPV |
|---|---|---|---|---|---|---|
| Random | 0.50 | 0.50 | 0.50 | 0.50 | 0.50 | 0.50 |
| Normal vs Impaired | | | | | | |
| CNN | 0.62±0.05 | 0.56±0.03 | 0.61±0.03 | 0.64±0.07 | 0.54±0.06 | 0.82±0.01 |
| ResNet-18 | 0.67±0.03 | 0.55±0.01 | 0.58±0.03 | 0.77±0.08 | 0.35±0.12 | 0.79±0.01 |
| XGBoost | 0.60±0.04 | 0.54±0.02 | 0.58±0.02 | 0.63±0.07 | 0.51±0.09 | 0.81±0.02 |

| | | | | | | |
|---|---|---|---|---|---|---|
| Age only | 0.63±0.04 | 0.54±0.03 | 0.53±0.04 | 0.70±0.05 | 0.41±0.06 | 0.79±0.02 |
| CNN + age | 0.63±0.03 | 0.55±0.02 | 0.61±0.02 | 0.69±0.05 | 0.45±0.04 | 0.80±0.01 |
| ResNet-18 + age | 0.69±0.04 | 0.53±0.02 | 0.57±0.04 | 0.83±0.08 | 0.23±0.09 | 0.78±0.01 |
| XGBoost + age | 0.63±0.02 | 0.55±0.01 | 0.59±0.02 | 0.68±0.03 | 0.45±0.05 | 0.80±0.01 |
| Normal vs Diabetes | | | | | | |
| CNN | 0.60±0.04 | 0.59±0.03 | 0.65±0.03 | 0.59±0.07 | 0.62±0.05 | 0.74±0.02 |
| ResNet-18 | 0.62±0.02 | 0.58±0.03 | 0.62±0.03 | 0.70±0.03 | 0.46±0.08 | 0.71±0.02 |
| XGBoost | 0.60±0.01 | 0.58±0.02 | 0.62±0.01 | 0.61±0.04 | 0.57±0.08 | 0.73±0.02 |
| Age only | 0.59±0.04 | 0.55±0.03 | 0.57±0.04 | 0.67±0.09 | 0.44±0.09 | 0.69±0.02 |
| CNN + age | 0.62±0.04 | 0.60±0.03 | 0.65±0.03 | 0.64±0.08 | 0.59±0.04 | 0.74±0.01 |
| ResNet-18 + age | 0.63±0.03 | 0.57±0.02 | 0.61±0.04 | 0.77±0.08 | 0.36±0.09 | 0.69±0.02 |
| XGBoost + age | 0.62±0.01 | 0.60±0.02 | 0.65±0.03 | 0.65±0.03 | 0.56±0.05 | 0.73±0.02 |

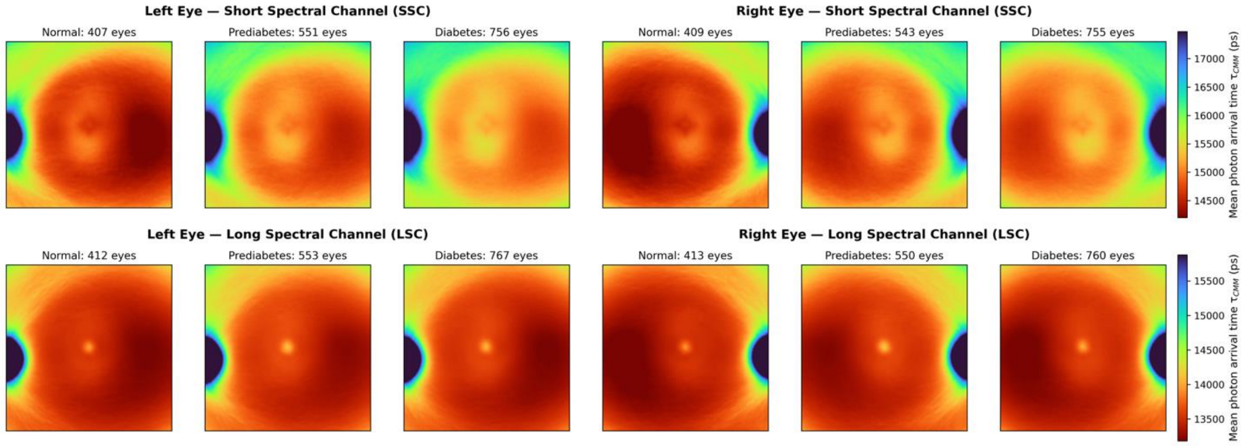


**Figure A6.** Counterpart of main-text Figure 2 (diagnosis- and treatment-aware labels; uncorrected τCMM maps). Aggregated τCMM maps by glycemic group, spectral channel (SSC, top; LSC, bottom), and eye laterality; sample counts (eyes) above each panel; color scale from red (short) to blue (long), in picoseconds. Uncorrected values include a system timing offset, and the apparently long foveal LSC values reflect background-related intensity coupling of the uncorrected estimator (Section B).

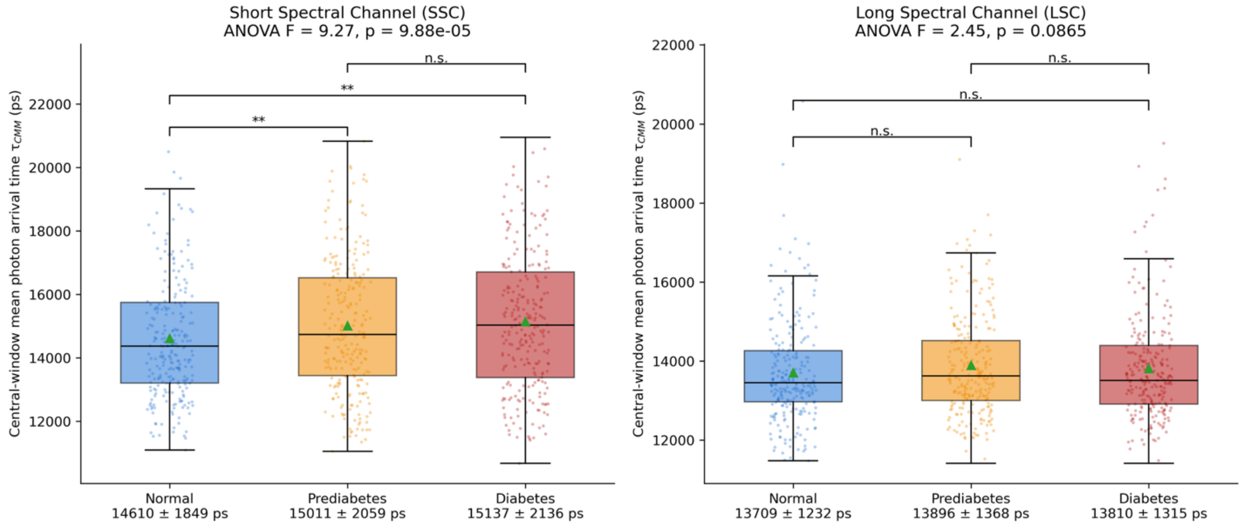


**Figure A7.** Counterpart of main-text Figure 4 (diagnosis- and treatment-aware labels; uncorrected τCMM maps). Participant-level distributions of τCMM within the central 20 × 20-pixel window by glycemic group and spectral channel; one-way ANOVA F and p in each panel title; brackets denote Bonferroni-corrected pairwise Welch's t-tests (n.s. = not significant; *p < 0.05; **p < 0.01); group means ± standard deviation (ps) below each x-axis label.

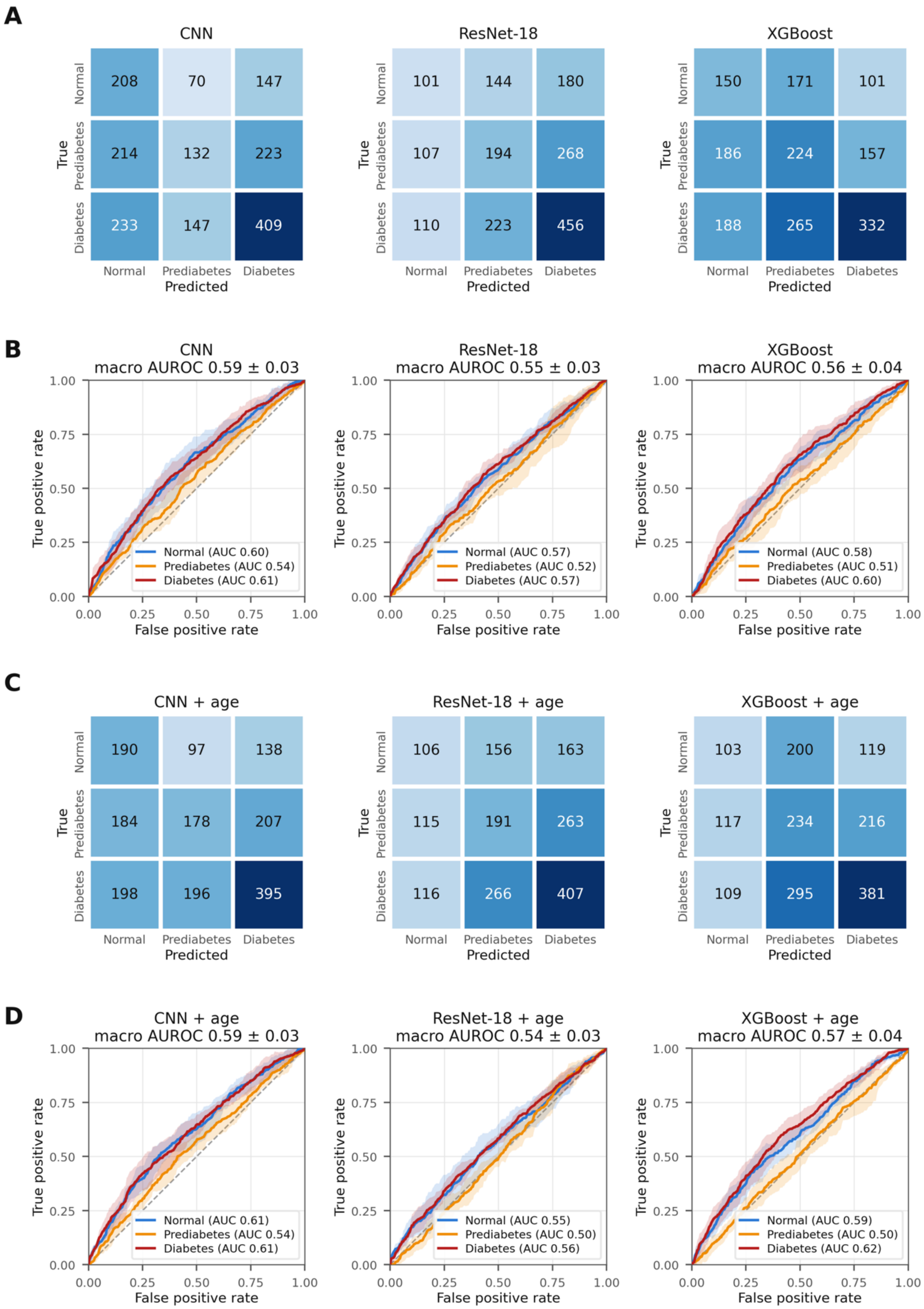
A
CNN
ResNet-18
XGBoost
Normal
Prediabetes
Diabetes
True
Predicted
208 70 147
214 132 223
233 147 409
101 144 180
107 194 268
110 223 456
150 171 101
186 224 157
188 265 332
B
CNN
macro AUROC 0.59 ± 0.03
ResNet-18
macro AUROC 0.55 ± 0.03
XGBoost
macro AUROC 0.56 ± 0.04
True positive rate
False positive rate
Normal (AUC 0.60)
Prediabetes (AUC 0.54)
Diabetes (AUC 0.61)
Normal (AUC 0.57)
Prediabetes (AUC 0.52)
Diabetes (AUC 0.57)
Normal (AUC 0.58)
Prediabetes (AUC 0.51)
Diabetes (AUC 0.60)
C
CNN + age
ResNet-18 + age
XGBoost + age
190 97 138
184 178 207
198 196 395
106 156 163
115 191 263
116 266 407
103 200 119
117 234 216
109 295 381
D
CNN + age
macro AUROC 0.59 ± 0.03
ResNet-18 + age
macro AUROC 0.54 ± 0.03
XGBoost + age
macro AUROC 0.57 ± 0.04
Normal (AUC 0.61)
Prediabetes (AUC 0.54)
Diabetes (AUC 0.61)
Normal (AUC 0.55)
Prediabetes (AUC 0.50)
Diabetes (AUC 0.56)
Normal (AUC 0.59)
Prediabetes (AUC 0.50)
Diabetes (AUC 0.62)

**Figure A8.** Counterpart of main-text Figure 5 (diagnosis- and treatment-aware labels; uncorrected τCMM maps). (A) Confusion matrices (participant counts, pooled across outer test folds) and (B) per-class ROC curves (mean ± fold-to-fold standard deviation; macro AUROC in panel titles) for the CNN, ResNet-18, and XGBoost FLIO-only models; (C, D) the same for the models with age as an additional input.

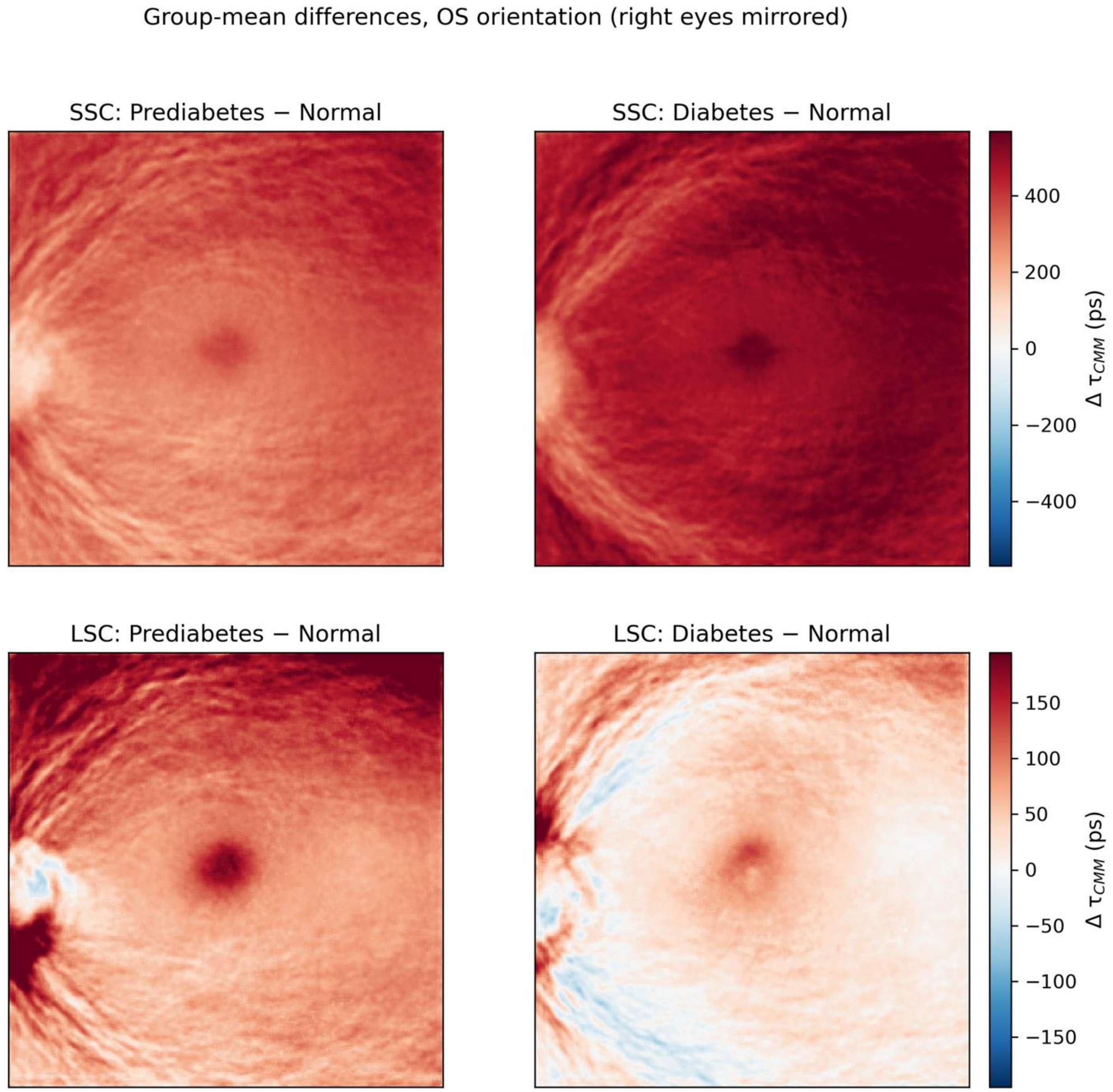


**Figure A9.** Counterpart of main-text Figure 3 (diagnosis- and treatment-aware labels; uncorrected τCMM maps). Pixel-wise group-mean differences in OS orientation (Prediabetes − Normal, left; Diabetes − Normal, right) for the SSC (top) and LSC (bottom); warm colors indicate longer τCMM in the impaired-glycemia group; color bars in picoseconds. Uncorrected values include a system timing offset, and the apparently long foveal LSC values reflect background-related intensity coupling of the uncorrected estimator (Section B).

**F.2 HbA1c-only labels, background- and timing-offset-corrected maps**

Central-window comparison: SSC ANOVA $F = 6.53$, $p = 0.002$ (age-adjusted $p = 0.072$); LSC ANOVA $F = 7.14$, $p < 0.001$ (age-adjusted $p = 0.64$) (Figures A10–A13).

**Table A7.** Model performance for 3-class classification, macro-averaged (HbA1c-only labels; background- and timing-offset-corrected τCMM maps). Models and cohorts as in main-text Table 2; XGBoost and Age only were evaluated on the 1,773 participants with complete ETDRS features, CNN and ResNet-18 on all 1,783 participants. Values are mean ± standard deviation across 5 outer test folds of nested cross-validation. AUROC = area under the receiver operating characteristic curve; CNN = convolutional neural network; PPV = positive predictive value; Random = uninformed random prediction.

| Model | Accuracy | F1 Score | AUROC | Sensitivity | Specificity | PPV |
|---|---|---|---|---|---|---|
| Random | 0.33 | 0.33 | 0.50 | 0.33 | 0.67 | 0.33 |
| CNN | 0.41±0.03 | 0.39±0.03 | 0.57±0.01 | 0.40±0.02 | 0.70±0.01 | 0.40±0.03 |
| ResNet-18 | 0.40±0.02 | 0.35±0.02 | 0.53±0.03 | 0.36±0.02 | 0.68±0.01 | 0.36±0.02 |
| XGBoost | 0.41±0.02 | 0.38±0.04 | 0.58±0.02 | 0.39±0.03 | 0.69±0.01 | 0.39±0.02 |
| Age only | 0.38±0.05 | 0.33±0.01 | 0.54±0.01 | 0.35±0.02 | 0.68±0.01 | 0.33±0.03 |
| CNN + age | 0.39±0.02 | 0.37±0.02 | 0.58±0.02 | 0.39±0.02 | 0.69±0.01 | 0.39±0.02 |
| ResNet-18 + age | 0.38±0.03 | 0.34±0.03 | 0.53±0.02 | 0.35±0.03 | 0.68±0.02 | 0.36±0.03 |
| XGBoost + age | 0.41±0.02 | 0.40±0.02 | 0.59±0.02 | 0.42±0.02 | 0.70±0.01 | 0.41±0.02 |

**Table A8.** Model performance for binary classification tasks (HbA1c-only labels; background- and timing-offset-corrected τCMM maps). For Normal vs. Impaired, prediabetic and diabetic participants were merged into a single Impaired class; for Normal vs. Diabetes, prediabetic participants were excluded. Models and cohorts as in main-text Table 2; XGBoost and Age only were evaluated on the 1,773 participants with complete ETDRS features, CNN and ResNet-18 on all 1,783 participants. Values are mean ± standard deviation across 5 outer test folds of nested cross-validation. AUROC = area under the receiver operating characteristic curve; CNN = convolutional neural network; PPV = positive predictive value; Random = uninformed random prediction.

| Model | Accuracy | F1 Score | AUROC | Sensitivity | Specificity | PPV |
|---|---|---|---|---|---|---|
| Random | 0.50 | 0.50 | 0.50 | 0.50 | 0.50 | 0.50 |
| Normal vs Impaired | | | | | | |
| CNN | 0.59±0.03 | 0.57±0.03 | 0.61±0.03 | 0.60±0.02 | 0.56±0.05 | 0.69±0.03 |
| ResNet-18 | 0.58±0.04 | 0.54±0.02 | 0.57±0.02 | 0.68±0.10 | 0.40±0.07 | 0.65±0.01 |
| XGBoost | 0.57±0.03 | 0.56±0.03 | 0.60±0.02 | 0.55±0.05 | 0.59±0.03 | 0.69±0.02 |
| Age only | 0.58±0.05 | 0.55±0.03 | 0.57±0.03 | 0.63±0.13 | 0.48±0.10 | 0.67±0.02 |
| CNN + age | 0.61±0.03 | 0.58±0.03 | 0.60±0.04 | 0.72±0.06 | 0.43±0.06 | 0.68±0.02 |
| ResNet-18 + age | 0.59±0.03 | 0.55±0.03 | 0.56±0.04 | 0.68±0.12 | 0.44±0.14 | 0.67±0.02 |
| XGBoost + age | 0.58±0.02 | 0.57±0.02 | 0.61±0.03 | 0.58±0.03 | 0.57±0.03 | 0.69±0.01 |
| Normal vs Diabetes | | | | | | |
| CNN | 0.63±0.04 | 0.60±0.06 | 0.64±0.03 | 0.53±0.17 | 0.70±0.09 | 0.50±0.06 |

| | | | | | | |
|---|---|---|---|---|---|---|
| ResNet-18 | 0.60±0.03 | 0.54±0.06 | 0.57±0.08 | 0.35±0.14 | 0.74±0.05 | 0.42±0.08 |
| XGBoost | 0.63±0.04 | 0.61±0.04 | 0.67±0.05 | 0.54±0.06 | 0.69±0.07 | 0.50±0.05 |
| Age only | 0.56±0.03 | 0.55±0.02 | 0.59±0.04 | 0.54±0.06 | 0.58±0.07 | 0.42±0.03 |
| CNN + age | 0.62±0.02 | 0.60±0.03 | 0.63±0.03 | 0.57±0.15 | 0.65±0.09 | 0.48±0.02 |
| ResNet-18 + age | 0.56±0.02 | 0.51±0.03 | 0.55±0.03 | 0.34±0.13 | 0.69±0.08 | 0.38±0.04 |
| XGBoost + age | 0.64±0.05 | 0.62±0.05 | 0.67±0.05 | 0.54±0.05 | 0.70±0.05 | 0.52±0.06 |

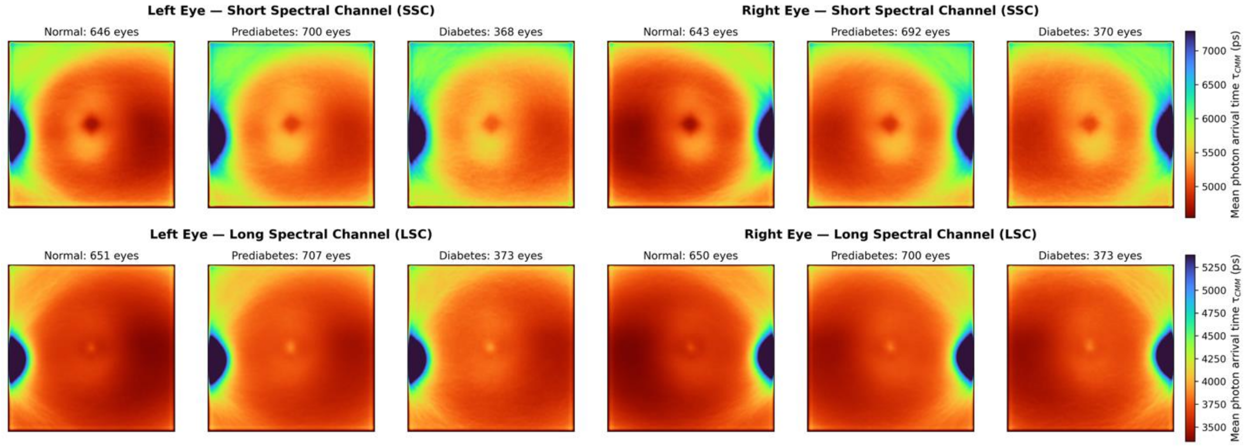


**Figure A10.** Counterpart of main-text Figure 2 (HbA1c-only labels; background- and timing-offset-corrected τCMM maps). Aggregated τCMM maps by glycemic group, spectral channel (SSC, top; LSC, bottom), and eye laterality; sample counts (eyes) above each panel; color scale from red (short) to blue (long), in picoseconds.

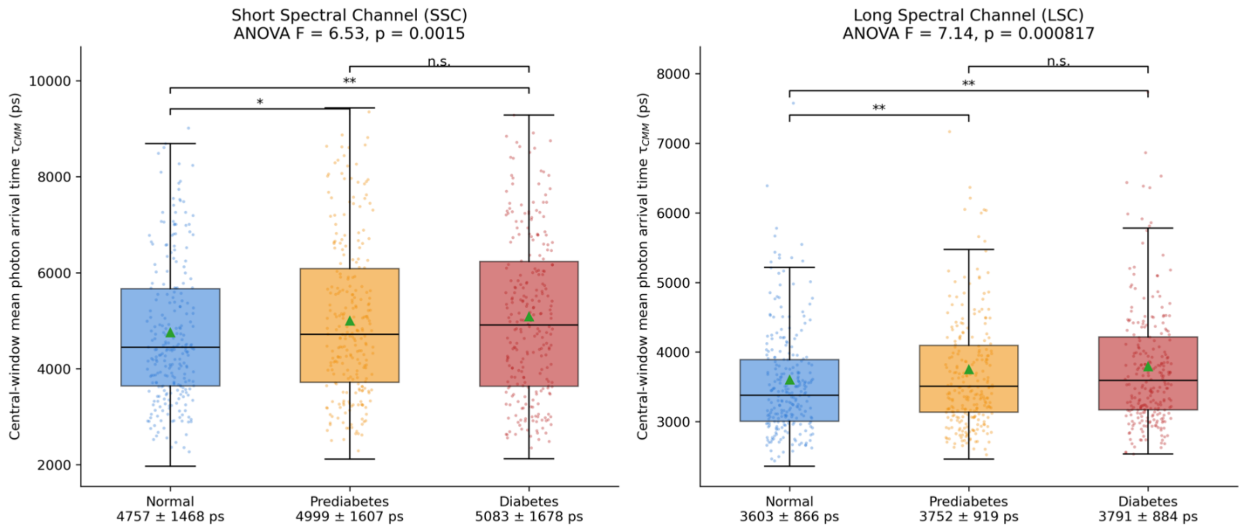


**Figure A11.** Counterpart of main-text Figure 4 (HbA1c-only labels; background- and timing-offset-corrected τCMM maps). Participant-level distributions of τCMM within the central 20 × 20-pixel window by glycemic group and spectral channel; one-way ANOVA F and p in each panel title; brackets denote Bonferroni-corrected pairwise Welch's t-tests (n.s. = not significant; *p < 0.05; **p < 0.01); group means ± standard deviation (ps) below each x-axis label.

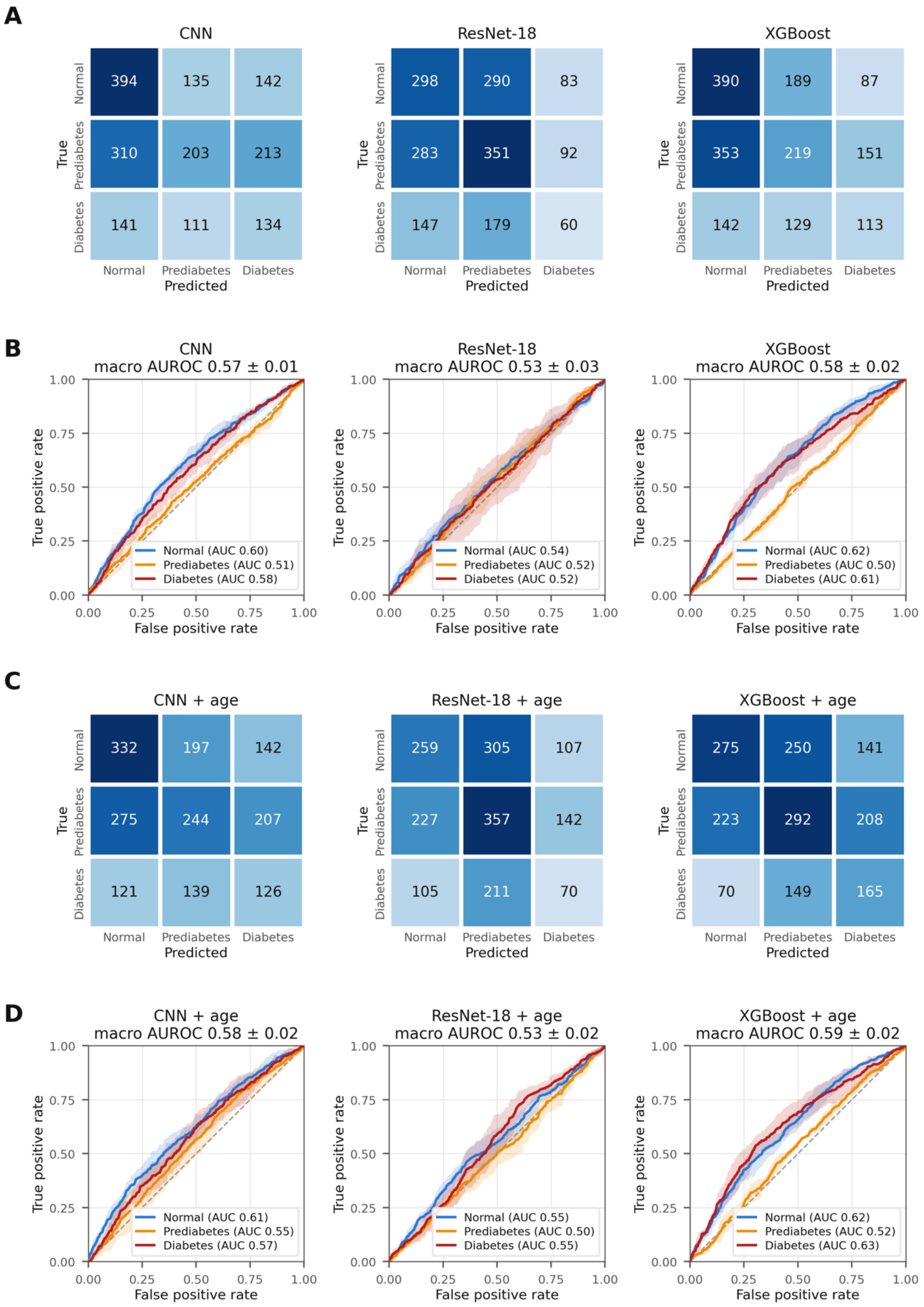
A
CNN
ResNet-18
XGBoost
True
Normal
Prediabetes
Diabetes
Predicted
394 135 142
310 203 213
141 111 134
298 290 83
283 351 92
147 179 60
390 189 87
353 219 151
142 129 113
B
CNN
macro AUROC 0.57 ± 0.01
ResNet-18
macro AUROC 0.53 ± 0.03
XGBoost
macro AUROC 0.58 ± 0.02
True positive rate
False positive rate
0.00
0.25
0.50
0.75
1.00
Normal (AUC 0.60)
Prediabetes (AUC 0.51)
Diabetes (AUC 0.58)
Normal (AUC 0.54)
Prediabetes (AUC 0.52)
Diabetes (AUC 0.52)
Normal (AUC 0.62)
Prediabetes (AUC 0.50)
Diabetes (AUC 0.61)
C
CNN + age
ResNet-18 + age
XGBoost + age
332 197 142
275 244 207
121 139 126
259 305 107
227 357 142
105 211 70
275 250 141
223 292 208
70 149 165
D
CNN + age
macro AUROC 0.58 ± 0.02
ResNet-18 + age
macro AUROC 0.53 ± 0.02
XGBoost + age
macro AUROC 0.59 ± 0.02
Normal (AUC 0.61)
Prediabetes (AUC 0.55)
Diabetes (AUC 0.57)
Normal (AUC 0.55)
Prediabetes (AUC 0.50)
Diabetes (AUC 0.55)
Normal (AUC 0.62)
Prediabetes (AUC 0.52)
Diabetes (AUC 0.63)

**Figure A12.** Counterpart of main-text Figure 5 (HbA1c-only labels; background- and timing-offset-corrected τCMM maps). (A) Confusion matrices (participant counts, pooled across outer test folds) and (B) per-class ROC curves (mean ± fold-to-fold standard deviation; macro AUROC in panel titles) for the CNN, ResNet-18, and XGBoost FLIO-only models; (C, D) the same for the models with age as an additional input.

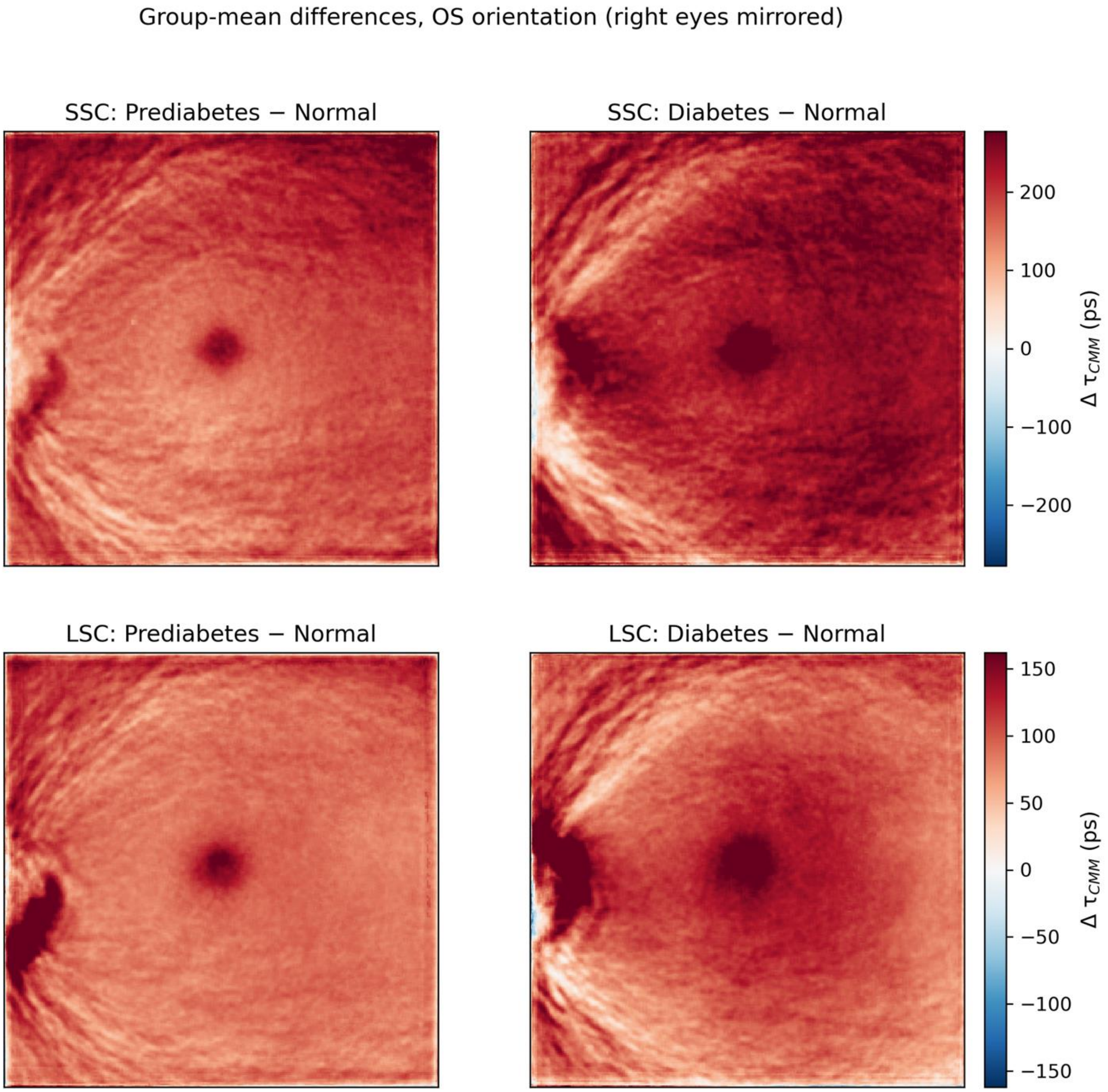


**Figure A13.** Counterpart of main-text Figure 3 (HbA1c-only labels; background- and timing-offset-corrected τCMM maps). Pixel-wise group-mean differences in OS orientation (Prediabetes − Normal, left; Diabetes − Normal, right) for the SSC (top) and LSC (bottom); warm colors indicate longer $\tau$CMM in the impaired-glycemia group; color bars in picoseconds.

**F.3 HbA1c-only labels, uncorrected maps**

Central-window comparison: SSC ANOVA $F = 6.85$, $p = 0.001$ (age-adjusted $p = 0.049$); LSC ANOVA $F = 2.89$, $p = 0.056$ (age-adjusted $p = 0.73$) (Figures A14–A17).

**Table A9.** Model performance for 3-class classification, macro-averaged (HbA1c-only labels; uncorrected $\tau$CMM maps). Models and cohorts as in main-text Table 2; XGBoost and Age only were evaluated on the 1,774 participants with complete ETDRS features, CNN and ResNet-18 on all 1,783 participants. Values are mean ± standard deviation across 5 outer test folds of nested cross-validation. AUROC = area under the receiver operating characteristic curve; CNN = convolutional neural network; PPV = positive predictive value; Random = uninformed random prediction.

| Model | Accuracy | F1 Score | AUROC | Sensitivity | Specificity | PPV |
|---|---|---|---|---|---|---|
| Random | 0.33 | 0.33 | 0.50 | 0.33 | 0.67 | 0.33 |
| CNN | 0.40±0.02 | 0.38±0.02 | 0.57±0.02 | 0.40±0.03 | 0.70±0.01 | 0.39±0.03 |
| ResNet-18 | 0.39±0.02 | 0.35±0.02 | 0.53±0.03 | 0.37±0.03 | 0.68±0.01 | 0.37±0.02 |
| XGBoost | 0.39±0.03 | 0.34±0.03 | 0.54±0.02 | 0.36±0.02 | 0.68±0.01 | 0.36±0.03 |
| Age only | 0.38±0.04 | 0.33±0.03 | 0.53±0.02 | 0.35±0.02 | 0.68±0.01 | 0.34±0.03 |
| CNN + age | 0.40±0.01 | 0.39±0.01 | 0.58±0.02 | 0.40±0.02 | 0.70±0.01 | 0.39±0.02 |
| ResNet-18 + age | 0.38±0.03 | 0.35±0.02 | 0.53±0.01 | 0.36±0.02 | 0.68±0.01 | 0.36±0.02 |
| XGBoost + age | 0.40±0.03 | 0.38±0.04 | 0.58±0.02 | 0.38±0.03 | 0.69±0.02 | 0.39±0.02 |

**Table A10.** Model performance for binary classification tasks (HbA1c-only labels; uncorrected τCMM maps). For Normal vs. Impaired, prediabetic and diabetic participants were merged into a single Impaired class; for Normal vs. Diabetes, prediabetic participants were excluded. Models and cohorts as in main-text Table 2; XGBoost and Age only were evaluated on the 1,774 participants with complete ETDRS features, CNN and ResNet-18 on all 1,783 participants. Values are mean ± standard deviation across 5 outer test folds of nested cross-validation. AUROC = area under the receiver operating characteristic curve; CNN = convolutional neural network; PPV = positive predictive value; Random = uninformed random prediction.

| Model | Accuracy | F1 Score | AUROC | Sensitivity | Specificity | PPV |
|---|---|---|---|---|---|---|
| Random | 0.50 | 0.50 | 0.50 | 0.50 | 0.50 | 0.50 |
| Normal vs Impaired | | | | | | |
| CNN | 0.59±0.02 | 0.58±0.02 | 0.61±0.03 | 0.63±0.06 | 0.53±0.05 | 0.69±0.01 |
| ResNet-18 | 0.57±0.03 | 0.52±0.02 | 0.54±0.04 | 0.70±0.10 | 0.35±0.13 | 0.64±0.02 |
| XGBoost | 0.56±0.01 | 0.55±0.01 | 0.59±0.02 | 0.55±0.01 | 0.57±0.04 | 0.68±0.02 |
| Age only | 0.58±0.02 | 0.55±0.02 | 0.57±0.03 | 0.68±0.06 | 0.43±0.07 | 0.66±0.02 |
| CNN + age | 0.60±0.03 | 0.58±0.04 | 0.62±0.05 | 0.65±0.04 | 0.52±0.12 | 0.70±0.04 |
| ResNet-18 + age | 0.56±0.02 | 0.53±0.02 | 0.54±0.03 | 0.66±0.06 | 0.41±0.07 | 0.65±0.01 |
| XGBoost + age | 0.58±0.03 | 0.57±0.03 | 0.61±0.02 | 0.62±0.05 | 0.53±0.04 | 0.69±0.02 |
| Normal vs Diabetes | | | | | | |
| CNN | 0.61±0.01 | 0.58±0.02 | 0.62±0.02 | 0.51±0.08 | 0.67±0.04 | 0.47±0.02 |

| | | | | | | |
|---|---|---|---|---|---|---|
| ResNet-18 | 0.58±0.03 | 0.57±0.03 | 0.60±0.06 | 0.54±0.12 | 0.61±0.04 | 0.44±0.03 |
| XGBoost | 0.63±0.04 | 0.61±0.04 | 0.64±0.04 | 0.51±0.04 | 0.70±0.04 | 0.50±0.05 |
| Age only | 0.56±0.03 | 0.55±0.02 | 0.58±0.04 | 0.54±0.09 | 0.57±0.06 | 0.42±0.03 |
| CNN + age | 0.61±0.02 | 0.60±0.02 | 0.64±0.01 | 0.62±0.07 | 0.60±0.03 | 0.47±0.02 |
| ResNet-18 + age | 0.58±0.07 | 0.53±0.05 | 0.59±0.05 | 0.38±0.18 | 0.70±0.17 | 0.43±0.09 |
| XGBoost + age | 0.66±0.04 | 0.63±0.04 | 0.69±0.06 | 0.55±0.07 | 0.71±0.04 | 0.53±0.05 |

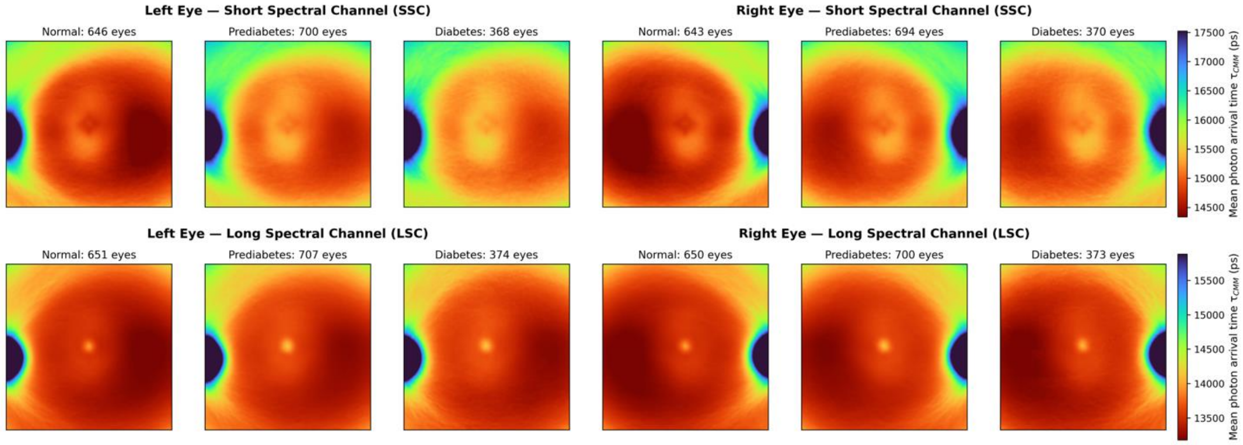


**Figure A14.** Counterpart of main-text Figure 2 (HbA1c-only labels; uncorrected τCMM maps). Aggregated τCMM maps by glycemic group, spectral channel (SSC, top; LSC, bottom), and eye laterality; sample counts (eyes) above each panel; color scale from red (short) to blue (long), in picoseconds. Uncorrected values include a system timing offset, and the apparently long foveal LSC values reflect background-related intensity coupling of the uncorrected estimator (Section B).

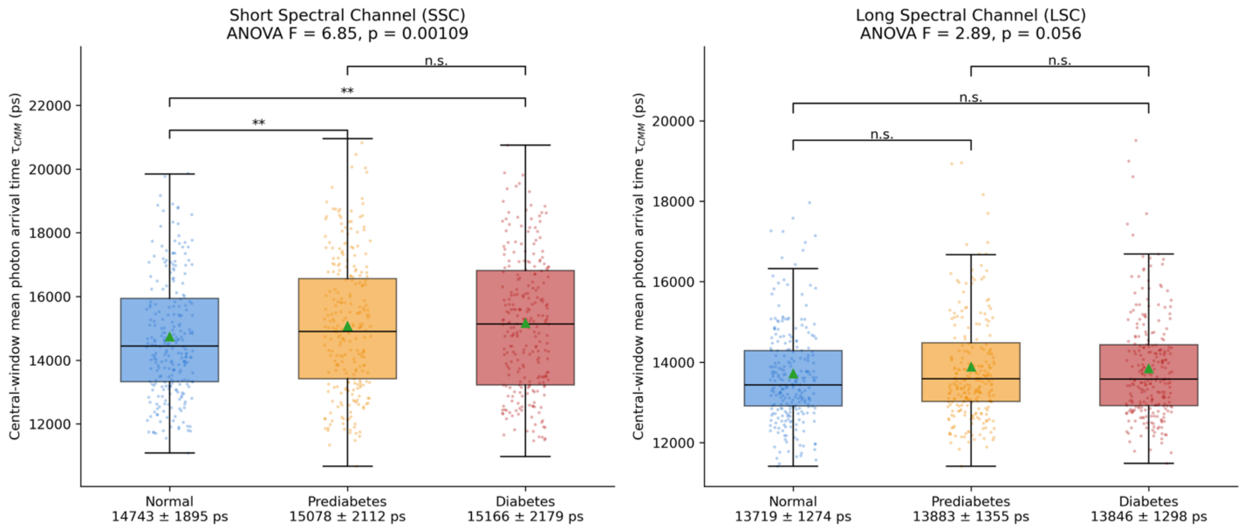


**Figure A15.** Counterpart of main-text Figure 4 (HbA1c-only labels; uncorrected τCMM maps). Participant-level distributions of τCMM within the central 20 × 20-pixel window by glycemic group and spectral channel; one-way ANOVA F and p in each panel title; brackets denote Bonferroni-corrected pairwise Welch's t-tests (n.s. = not significant; *p < 0.05; **p < 0.01); group means ± standard deviation (ps) below each x-axis label.

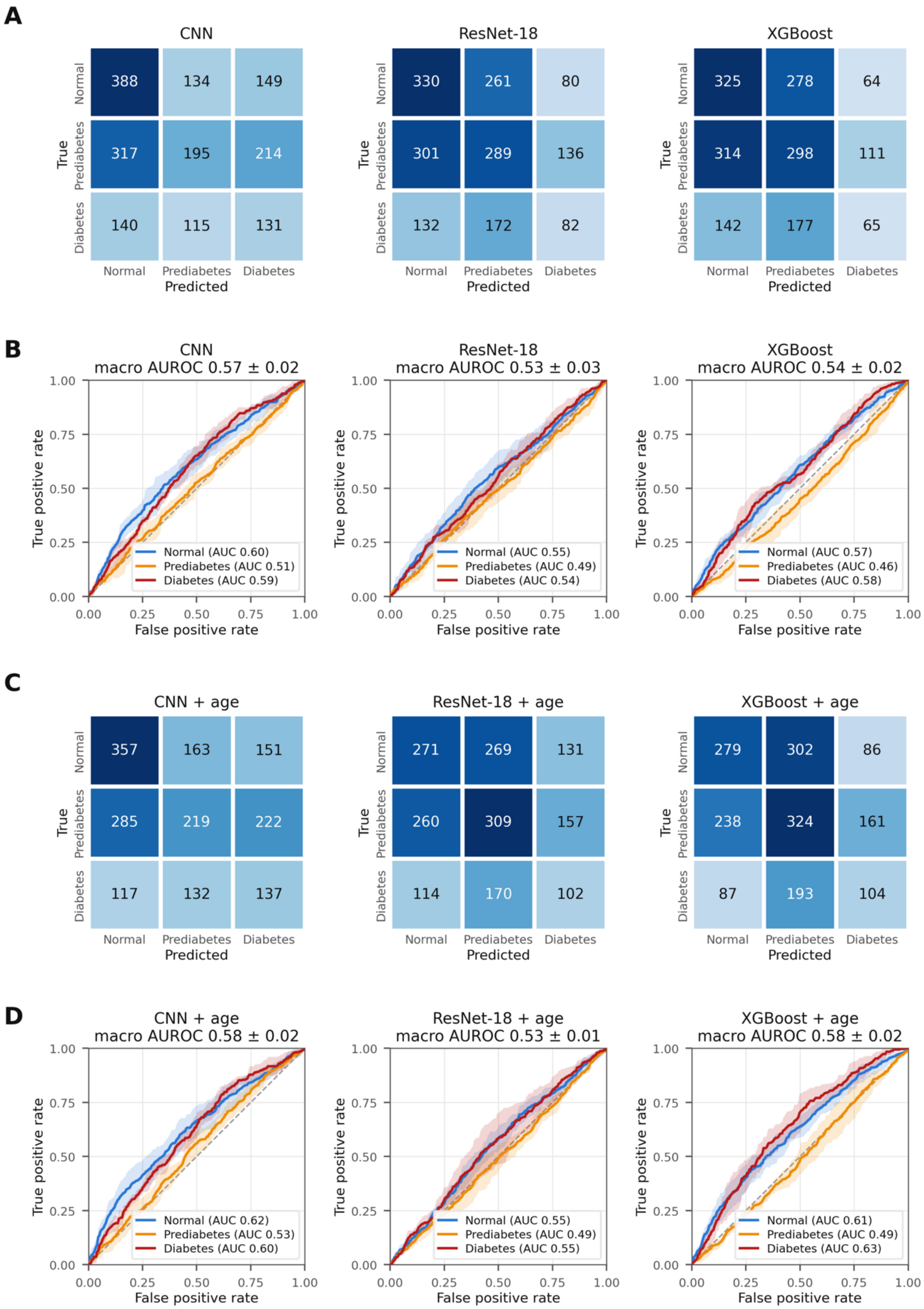
A
CNN
ResNet-18
XGBoost
388 134 149
317 195 214
140 115 131
330 261 80
301 289 136
132 172 82
325 278 64
314 298 111
142 177 65
True
Normal
Prediabetes
Diabetes
Predicted
B
CNN
macro AUROC 0.57 ± 0.02
ResNet-18
macro AUROC 0.53 ± 0.03
XGBoost
macro AUROC 0.54 ± 0.02
Normal (AUC 0.60)
Prediabetes (AUC 0.51)
Diabetes (AUC 0.59)
Normal (AUC 0.55)
Prediabetes (AUC 0.49)
Diabetes (AUC 0.54)
Normal (AUC 0.57)
Prediabetes (AUC 0.46)
Diabetes (AUC 0.58)
True positive rate
False positive rate
0.00
0.25
0.50
0.75
1.00
C
CNN + age
ResNet-18 + age
XGBoost + age
357 163 151
285 219 222
117 132 137
271 269 131
260 309 157
114 170 102
279 302 86
238 324 161
87 193 104
D
CNN + age
macro AUROC 0.58 ± 0.02
ResNet-18 + age
macro AUROC 0.53 ± 0.01
XGBoost + age
macro AUROC 0.58 ± 0.02
Normal (AUC 0.62)
Prediabetes (AUC 0.53)
Diabetes (AUC 0.60)
Normal (AUC 0.55)
Prediabetes (AUC 0.49)
Diabetes (AUC 0.55)
Normal (AUC 0.61)
Prediabetes (AUC 0.49)
Diabetes (AUC 0.63)

**Figure A16.** Counterpart of main-text Figure 5 (HbA1c-only labels; uncorrected τCMM maps). (A) Confusion matrices (participant counts, pooled across outer test folds) and (B) per-class ROC curves (mean ± fold-to-fold standard deviation; macro AUROC in panel titles) for the CNN, ResNet-18, and XGBoost FLIO-only models; (C, D) the same for the models with age as an additional input.

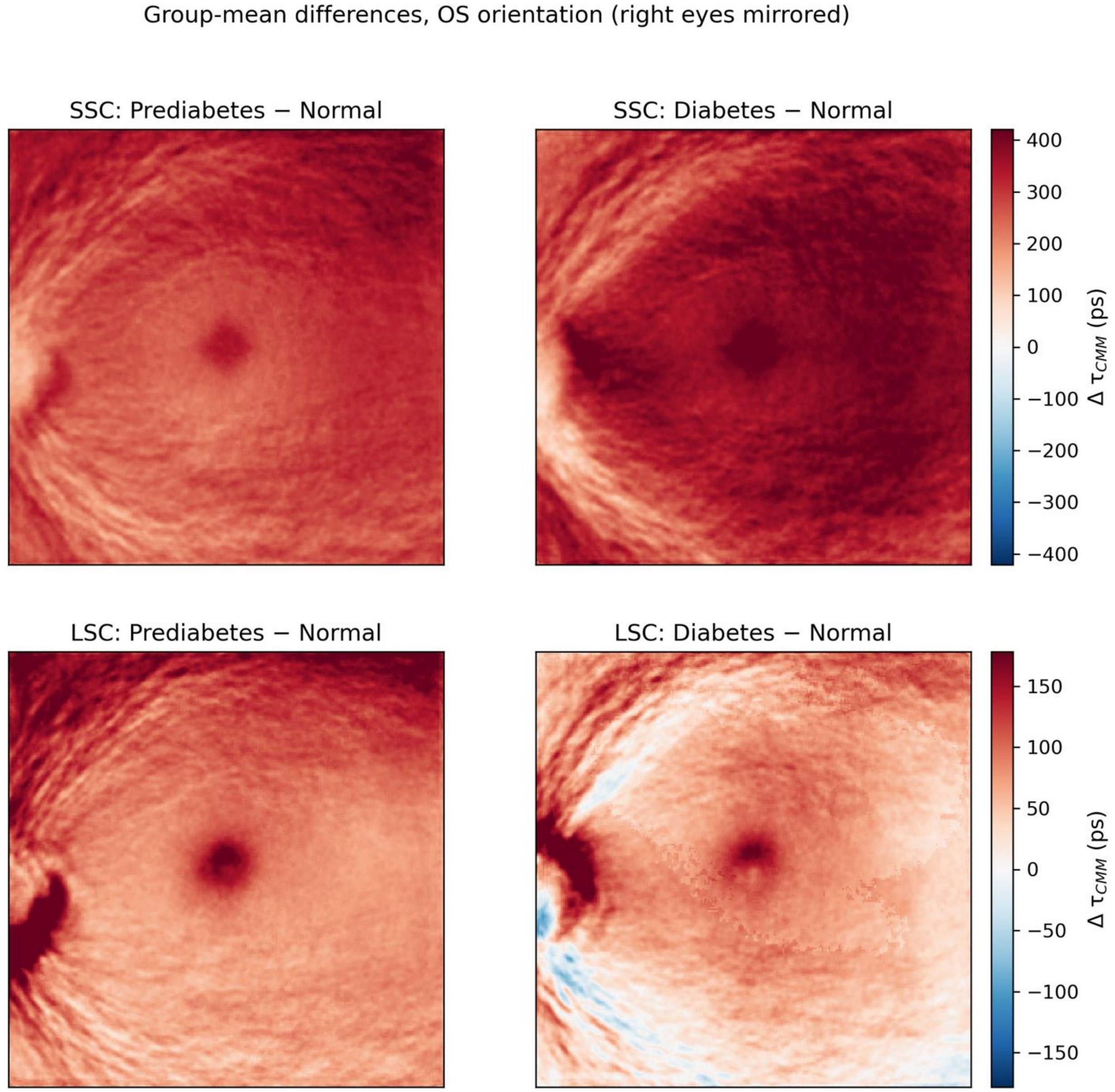


**Figure A17.** Counterpart of main-text Figure 3 (HbA1c-only labels; uncorrected τCMM maps). Pixel-wise group-mean differences in OS orientation (Prediabetes − Normal, left; Diabetes − Normal, right) for the SSC (top) and LSC (bottom); warm colors indicate longer τCMM in the impaired-glycemia group;

color bars in picoseconds. Uncorrected values include a system timing offset, and the apparently long foveal LSC values reflect background-related intensity coupling of the uncorrected estimator (Section B).
